# Mpox knowledge, attitudes, beliefs, and intended behaviour in the general population and men who are gay, bisexual, and who have sex with men

**DOI:** 10.1101/2022.12.07.22283201

**Authors:** Louise E Smith, Henry WW Potts, Julii Brainard, Tom May, Isabel Oliver, Richard Amlôt, Lucy Yardley, G James Rubin

## Abstract

**Objectives:** To investigate rates of mpox beliefs, knowledge, and intended behaviours in the general population and in gay, bisexual or other men who have sex with men (GBMSM), and factors associated with intended behaviours. To test the impact of motivational messages (vs a factual control) on intended behaviours.

**Design:** Cross-sectional online survey including a nested randomised controlled trial.

**Setting:** Data collected 5 September to 6 October 2022.

**Participants:** Participants were aged 18 years and over and lived in the UK (general population). In addition, GBMSM were male, and gay, bisexual or had sex with men. The general population sample was recruited through a market research company. GBMSM were recruited through a market research company, the dating app Grindr, and targeted adverts on Meta (Facebook and Instagram).

**Main outcome measures:** Intention to self-isolate, seek medical help, stop all sexual contact, share details of recent sexual contacts, and accept vaccination.

**Results:** Socio-demographic characteristics differed by sample. There was no effect of very brief motivational messaging on behavioural intentions. Respondents from Grindr and Meta were more likely to intend to seek help immediately, completely stop sexual behaviour and be vaccinated or intend to be vaccinated, but being less likely to intend to self-isolate (*p*s<0.001). In the general population sample, intending to carry out protective behaviours was generally associated with being female, older, having less financial hardship, greater worry, higher perceived risk to others, and higher perceived susceptibility to and severity of mpox (*p*s<0.001). There were fewer associations with behaviours in the Grindr sample, possibly due to reduced power.

**Conclusions:** GBMSM were more likely to intend to enact protective behaviours, except for self-isolation. This may reflect targeted public health efforts and engagement with this group. Associations with socio-economic factors suggests that providing financial support may encourage people to engage with protective behaviours.

**STRENGTHS AND LIMITATIONS:**

- Anonymous cross-sectional survey in large samples of the general population and men who are gay, bisexual, or have sex with men (recruited from a market research company, the dating app Grindr, and targeted adverts on Meta [Facebook and Instagram]).
- Data collection occurred over a short period (5 September to 6 October 2022) during the mpox outbreak.
- Responses may have been affected by social desirability or recall bias, although the anonymous nature of the survey should mitigate this somewhat.
- Socio-demographic characteristics differed by sample. Participants recruited from Grindr and Meta were more likely to be working, highly educated, of higher socio-economic grade, and have less financial hardship.
- We measured behavioural intentions. Rates of engagement with behaviours may be lower. Factors associated with intentions should still be valid.

## INTRODUCTION

Mpox (also known as monkeypox) is an orthopox virus that causes fever, headache, exhaustion, swollen glands and aches (joint, muscle, back), followed by a rash with blisters.(1) It spreads from person to person through touching clothing, bedding or towels used by someone with mpox rash, touching mpox skin blisters or scabs, and through the coughs or sneezes of someone with mpox rash (droplet transmission).(1) Since May 2022, there has been a multi-country outbreak of mpox in non-endemic countries.(2) Estimates from the World Health Organization (WHO; data up to 1 December 2022), indicate that there have been over 81,000 cases in 110 countries, resulting in 59 deaths.(3) Most cases have been in men who are gay, bisexual, or have sex with men, with close human skin-to skin contact (including sexual) being the primary driver of transmission.(4) The UK is the sixth country most affected by this mpox outbreak with 3725 cases.(3) Within the UK, most cases have been identified in England (with 69% of English cases in London). Almost all (99%) cases were men; English cases had a median age of 37 years.(5) The peak of the epidemic was seen in June and July 2022, with case numbers falling since the end of July.(6)

In the UK, people who thought they might have mpox were asked to call a sexual health clinic, to stay at home (self-isolate) and avoid close contact with other people.(1) Suspected cases were tested for mpox, and confirmed cases were asked to self-isolate for up to 21 days and engage with contact tracing.(7) Cases, their close contacts, and those most likely to be exposed to mpox were advised to be vaccinated with modified vaccinia Ankara vaccine – which offers cross-protection – to reduce transmission and prevent severe illness.(8, 9) While similar public health actions are now familiar to the public as a result of the COVID-19 pandemic, research conducted during the pandemic indicates that engagement with uptake of testing and self-isolation was sub-optimal in some groups.(10–12)

Various theories have guided research into the psychological factors associated with the uptake of health behaviours. One such theory is the Protection Motivation Theory (PMT), which states that people’s intention to carry out a protective behaviour is influenced by their appraisal of the threat (perceived susceptibility to and severity of, e.g. mpox) and the coping response (perceived effectiveness of and self-efficacy for, e.g. testing, self-isolation, contact tracing, vaccination).(13) During the COVID-19 pandemic, psychological, and socio-demographic factors associated with testing and self-isolating included: higher perceived risk of COVID-19, knowledge of transmission modes, higher perceived effectiveness of protective behaviours, believing that your behaviour had an impact on transmission, and belief that others in the same position would also self-isolate, being female, and having less financial hardship.(10–12, 14) Generally speaking, engagement with protective behaviours was associated with being older, apart from uptake of lateral flow testing, which was higher in younger people.(10, 11) Factors associated with COVID-19 vaccine uptake (completed and intended) included perceiving vaccination to be safe and necessary, perceiving COVID-19 to be more severe, and thinking that others would also be vaccinated.(15–18) Perception of side effects is one of the most common reasons given for refusing vaccination.(19) Historically, smallpox vaccines have been associated with severe adverse effects.(8) While vaccines currently licensed are associated with fewer severe adverse effects, this may affect people’s intention to be vaccinated.

At the time of writing, few scientific studies have investigated behaviour during the 2022 mpox epidemic. While most have investigated vaccination acceptability, few have investigated engagement with a contact tracing system. Those studies that have been done suggest that knowledge of transmission modes is incomplete.(20–22) Greater agreement with vaccination for mpox was associated with perceiving the virus as more dangerous and virulent and higher worry about mpox in a survey of members of the Saudi Arabian general public.(23) In Dutch GBMSM, willingness to accept a vaccine was associated with being more worried about getting mpox, perceiving a higher risk of mpox, perceiving mpox to be more severe, thinking that vaccination is important, thinking that the vaccine was effective, and greater social norms for vaccination.(24) Another study also conducted in the Netherlands found that vaccination intention in GBMSM was associated with higher worry about mpox, knowing someone who had mpox, and being single but dating or in an open/polyamorous relationship.(25) This study also investigated self-isolation intention for 21 days, finding that higher intentions were associated with thinking that mpox had more problematic consequences, and lower education.(25) A study conducted in the UK found that agreement with self-isolation was associated with not having completed a higher education degree, not being employed, and identifying as having a disability.(26) There was no difference in agreement between GBMSM and those who were not GBMSM. Further research is needed to investigate how psychological factors may affect engagement with public health measures (isolating, testing, contact tracing, vaccination) put in place to control the spread of the mpox outbreak in the UK.

Official communications are vital during new and emerging outbreaks, and serve to inform the public about the threat, the public health response, and behaviours that people should engage with in order to protect themselves and others.(27) Messages based on theories of health behaviours, such as the PMT,(13) may therefore increase engagement with protective behaviours. For example, research suggests that COVID-19 vaccination intention increased when communications emphasised the safety and effectiveness of the vaccine and using social norms interventions (e.g. asking people to “join the millions” being vaccinated).(28) While findings relating to messages emphasising the benefits of vaccination to oneself and others were mixed, there is some evidence that the influence of these messages may be most evident in strongly hesitant groups.

In this study, we recruited a general population sample and three GBMSM samples from: a market research company, Grindr, and Meta (Facebook and Instagram). We investigated knowledge, attitudes and beliefs about mpox, and intentions for key behaviours that could prevent the spread of mpox (self-isolation, help seeking [as the advised route into testing], sexual contact behaviour when symptomatic, contact sharing, vaccination). We used an experimental approach to investigate the impact of different brief communication approaches (promoting perceived susceptibility to and severity of illness / necessity and efficacy of the response / benefits of the response / low perceived costs of response) on intended behaviour. Psychological and socio-demographic factors associated with intended uptake of behaviours were also investigated.

## METHOD

### Design

Online cross-sectional survey conducted by Savanta, a Market Research Society (MRS) Company Partner. Data were collected between 5 September and 6 October 2022.

### Participants

Eligibility criteria for the general population sample were living in the UK and being aged 18 years or over. For the GBMSM samples, additional criteria were being a man and identifying as being gay, bisexual, or having sex with men.

Recruitment for the general population sample used quota sampling, a standard opinion polling method that allows for rapid data collection. Members of Savanta’s specialist research panel (n=150,000 across proprietary panels; people who have signed up to complete online surveys) were sent the survey link. Quota sampling uses pre-determined targets, based on pre-selected socio-demographic characteristics (quotas) that match the national population.

Participants who belong to a quota that has already been filled are prevented from completing the survey. Therefore, response rate is not an accurate measure of response bias in quota samples. For this study, quotas were based on age, gender, socio-economic grade and Government Office Region, and reflected targets based on 2020 mid-year estimates.(29)

We recruited three GBMSM samples (Savanta, Grindr, Meta [Facebook and Instagram]). A “boost” sample of 250 GBMSM was recruited by Savanta, using the same quota sampling (excluding gender as all participants were male). We also recruited for the GBMSM sample through a push-notification inbox advert on Grindr (1.1 million adverts delivered, 24,933 opened, and 4288 clicks) and using targeted adverts on Meta (308,472 adverts shown, 108,865 adverts seen, and 3675 clicks). No quotas were placed on these samples.

### Study materials

Full study materials are in supplementary materials 1. Items were based on previously validated measures,(30, 31) and items used in previous surveys by our group.(11, 14, 32, 33)

#### Outcome measures

Self-isolation intention was measured using two items, asking participants to imagine that they were contacted by public health officials and told that they needed to self-isolate for 21 days because they had mpox, and because they had come into high-risk contact with a case. Responses were given on a five-point scale from “definitely would not” to “definitely would”. The order of items was randomised between participants to mitigate potential order effects. As answers on both items were highly correlated across all respondents (*r* = 0.77, *p* = 0.001), we combined responses to give a nine-point scale (2 to 10; summing responses “definitely would not” = 1 to “definitely would” = 5; higher number indicates greater intention).

To measure help-seeking intention, participants were asked to imagine that they developed an unexplained rash with blisters and learned that they had come into contact with a mpox case. They were then asked what they would do, from a list of ten behaviours including waiting to see if they got better, contacting healthcare services, letting people you had been in recent close contact know, and searching for information. Responses for each item were given on the same five-point “definitely would not” to “definitely would” scale. We created a binary variable, coding participants as intending to seek help immediately if they answered “probably” or “definitely would” to any help seeking where they would encounter a health professional (trying to book an appointment with a general practitioner, visiting a pharmacist/chemist, going to Accident & Emergency or another NHS service, calling NHS111 or 999, visiting a walk-in sexual health clinic, or calling a sexual health clinic), and did not select “wait a day or two to see if they get better or clear up on their own” vs did not select any help-seeking behaviour where they would encounter a health professional, or selected a help-seeking behaviour but also stated that they would “wait a day or two to see…”.

Participants were then asked about their intended contact behaviour in the same scenario, being asked “in the following 21 days, realistically how much” they would come into skin-to skin contact with others, have sexual contact, have sex without using a condom, go to a crowded place, help or provide care for a vulnerable person, and go to a public place where they may come into physical contact with someone else. For each item, participants responded “I would completely stop doing this”, “less than normal”, “same as normal”, “more than normal”, “not applicable, I wouldn’t do this anyway”, “don’t know” or “prefer not to say”. We focused our analyses on the item asking about sexual contact (from kissing to intercourse) with other people, recoding it into a binary item (“I would completely stop doing this” vs would do this less than, same as, or more than usual). Answers of “not applicable, I wouldn’t do this anyway”, “don’t know”, and “prefer not to say” were coded as missing.

We measured intention to share details of close contacts by asking participants to indicate how likely they were, if asked by public health officials, to share contact details of every person who had been in their home, they had had sexual contact with, they had skin-to-skin contact with, and that they had shared bedding, towels or clothes with in the last seven days, and every place they had had sexual contact with someone. Responses were given on a five-point “definitely would not” (1) to “definitely would” (5) scale. We used the most recent official guidance on contact tracing available at the start of data analysis to select the item most relevant to contact tracing efforts (identifying every person you had sexual contact with in the last seven days).(7)

Participants were asked if they had received a smallpox vaccine in 2022. Those who indicated they had not had a vaccine were asked about their vaccination intention. We asked participants how likely they would be to have a smallpox vaccine if they were offered one. Responses were given on a five-point “definitely would not” (1) to “definitely would” (5) scale.

#### Motivational messaging

Very brief motivational messages were constructed based on components of the PMT.(13) The general population sample were randomised to one of four groups, and were shown messages about: 1) perceived risk of illness and necessity and efficacy of the response, 2) perceived risk of illness plus benefits of the response, 3) perceived risk of illness plus low perceived costs of response, and 4) a control message of similar length giving factual details about the mpox outbreak. Messages are shown in supplementary materials 1. Messages to promote perceived risk of illness (i.e. susceptibility to and severity of mpox) were included in all motivational messages, as we hypothesised that perceiving a risk is necessary before choosing to adopt a behaviour to mitigate that risk. Due to anticipated smaller sample sizes, the GBMSM samples were randomised to one of two messages. The first included all motivational components, whereas the second, a control message, gave factual information about the mpox outbreak.

#### Psychological factors

We asked participants how much they had seen or heard about mpox, how worried they were about mpox, and how much risk they thought mpox posed to people in the UK and themselves personally. For these items, answers of “don’t know” were coded as missing.

Participants were also asked about their perceived susceptibility to mpox (two items: would be likely to come into contact with a case, would be likely to catch mpox if in contact with a case) and perceived severity of mpox. We asked participants how much they agreed that: their personal behaviour had an impact on the spread of mpox; their life had been negatively affected by changes made in response to the mpox outbreak; the risks of mpox were being exaggerated; people who catch mpox usually make a full recovery without treatment; and that mpox is only a risk to men who are gay, bisexual, or have sex with men. For perceptions such as these, with no right or wrong answers, we recoded answers of “don’t know” as the mid-point on the scale.

To measure perceived knowledge, we asked participants three items about whether they had a good idea how people catch mpox, they knew the main symptoms of mpox, and they thought it would be easy to tell if someone had mpox. Knowledge about mpox symptoms was measured using a question asking participants to identify the main symptoms of mpox from a list of fifteen taken from the NHS mpox website (1) and common, non-specific symptoms. Participants could select up to four symptoms. Understanding of transmission was measured using seven items, asking about contact and droplet transmission (adapted from (30), and other modes of transmission as specified by the World Health Organization (WHO) website (34)). For factual questions such as these, we coded “don’t know” as missing.

Behaviour-specific perceptions were also investigated. We used a series of ten items to measure factors that may be associated with self-isolation, including perceived effectiveness, social norms, having the necessary support, and impact on social connectedness, family wellbeing, and finances. Factors that may affect intention to seek help were measured by six items asking participants to what extent they agreed that they would not want to know the results of a mpox test, they would be worried what their friends, family, or employers thought of them if they had mpox, not wanting to have a mpox test on their medical record, testing is an effective way to prevent the spread of mpox, and being willing to contact a sexual health clinic if they thought they had mpox symptoms or had come into contact with a case. Vaccination attitudes were measured by eight items, asking about general vaccine attitudes, perceived social norms, perceived effectiveness of vaccination, worry about vaccine side-effects, that the vaccine could make you infectious to others, and thinking that those who come into high-risk contact with mpox should be vaccinated. Responses of “don’t know” were recoded to the mid-point of the scale.

#### Socio-demographic characteristics

Participants were asked to report their age, gender,(35) sexual orientation,(36) socio-economic grade,(37) financial hardship (adapted from (38)), employment status, highest level of education, ethnicity, marital status, how many people lived in their household, whether they were the parent or guardian of any dependent children, and if they had any pets. Those who were employed were asked if they were a frontline health or social care worker and if they needed to leave home for work. For these items, we coded participants who were not employed as not being a frontline health or social care worker (prefer not to say coded as missing) and not needing to leave home for work, respectively. Participants were asked for their full postcode, from which region and indices of multiple deprivation were determined.(39)

We asked participants if they or a household member had a chronic illness, whether they were pregnant, had ever taken pre-exposure prophylaxis (PrEP) for HIV, and for their vaccination status for smallpox (in 2022, and before 2022), hepatitis A, and COVID-19 (two doses or more).

Participants were also asked how many male and female sexual partners they had had in the last 3 weeks and last 3 months.

### Ethics

Ethical approval for this study was given by the King’s College London Psychiatry, Nursing and Midwifery Research Ethics Panel (reference number: LRS/DP-21/22-32287).

Participants gave informed consent before beginning survey materials.

### Patient and public involvement

To ensure the research aims and study information were appropriate, members of the public were involved in the development of the funding application and survey materials. For the funding application, six people gave feedback on the initial proposal resulting in changes to aims of the study and terminology used. For the survey materials, four lay people (two GBMSM) gave feedback on the questionnaire and motivational messages, resulting in changes to survey items and messages to improve clarity, validity, and readability of statements.

### Power

A sample size of 3000 allows a 95% confidence interval of plus or minus 1.8% for the prevalence estimate for a survey item with a prevalence of around 50% (sample size of 250 gives a confidence interval of plus or minus 6.2%; sample size of 1000 gives a confidence interval of plus or minus 3.1%).

For multiple linear regression analyses, a sample of 830 allowed over 99% power to detect small effect sizes (*f*=0.10) at *p*=0.001 (43 predictors). For logistic regression analyses, a sample of 830 allowed over 99% power to detect small effect sizes (OR=1.68 (40)) at *p*=0.001 (42.5% self-isolation, 18.0% requesting a test, 79.1% sharing details of close contacts (11)).

### Analysis

Information about data preparation is reported in supplementary materials 2.

We tested whether socio-demographic characteristics of GBMSM samples were different depending on the recruitment method (Savanta, Grindr, Meta). Due to significant differences, further analyses were conducted in each sample separately.

First, we tested the influence of motivational messages on outcomes using χ^2^ tests (binary outcomes), one-way ANOVAs (general population sample, continuous outcomes), or t-tests (GBMSM, continuous outcomes). For vaccination, we investigated the influence of motivational message on intention to be vaccinated if advised (excluding people already vaccinated).

Next, we investigated psychological and contextual factors associated with intended behaviours (self-isolation, help seeking, sexual contact, sharing details of contacts, vaccination). To minimise analyses conducted, we investigated one variable per outcome, except for vaccination (GBMSM sample investigated two outcomes). For vaccination, as smallpox vaccine uptake in GBMSM was high, we used two binary outcomes: vaccination uptake in 2022 (vaccinated vs not vaccinated), and a computed variable indicating vaccine intention and uptake (vaccinated or intend to be vaccinated [“definitely” or “probably would”] vs not vaccinated and do not intend to be vaccinated [“not sure”, “probably would not”, “definitely would not”]). For the general population sample, we used only the computed variable. We conducted regressions (binary logistic for binary outcomes, linear for continuous outcomes) in the general population sample and Grindr sample (the target GBMSM sample who were actively seeking new partners; Savanta sample excluded due to small numbers, Meta sample excluded as they differed significantly from the general population).

We entered variables into regressions in blocks. In the first block, we entered socio-demographic characteristics: gender (general population sample only, male / female), sexual orientation (general population sample only, straight or heterosexual / gay, lesbian, bisexual, or queer), age, region, having a dependent child in household (no / yes), employment status (working / not working), education (GCSE, vocational, A-level, no formal qualifications / degree or higher), ethnicity (White British / White other / Black, Asian, other minoritized ethnicity), marital status, living alone, having a chronic illness oneself (none / present), index of multiple deprivation (deciles), socio-economic grade (ABC1 / C2DE), financial hardship, and motivational message. In the Grindr sample, we also included smallpox vaccination status in 2022 (except for vaccine uptake outcome) and ever having taken PrEP for HIV.

In the second block, we entered psychological and contextual factors: self-reported knowledge, knowledge of mpox symptoms, knowledge of mpox transmission, amount heard about mpox, worry about mpox, perceived risk of mpox (to oneself and to people in the UK), perceived susceptibility and severity of mpox, thinking that you are immune to mpox, that your personal behaviour has an impact on how mpox spreads, that your life has been negatively affected by changes made in response to the mpox outbreak, that the risks of mpox are being exaggerated, that mpox is only a risk to men who are gay, bisexual or have sex with men, and that people who catch mpox usually make a full recovery even without treatment.

For self-isolation, help seeking and vaccination, a third block was also added, which included specific factors potentially associated with individual outcomes. Items were chosen through principal component analyses (see supplementary materials 3).

All analyses were carried out in SPSS 28. Data are unweighted.

Many comparisons were investigated in regression models (n=40 to n=43, based on outcome). Therefore, we applied a conservative Bonferroni correction and only report as significant results with *p*≤0.001 to reduce the risk of Type I errors.

## RESULTS

Top-line results for all survey materials, by sample, are shown in supplementary materials 1.

For regression analyses, we report imputed values. Results using imputed values were compared with non-imputed data. There were no substantial differences in results with and without imputed values.

### Participant characteristics

There were significant differences in participant characteristics by sampling method. Most notably, participants recruited from Grindr and Meta were more likely to be working, need to leave home for work, more highly educated, higher socio-economic grade, and have less financial hardship (Table 1). In the general population sample, participants were mostly female (57%), white British (87%), with a mean age of 49 years.

**Table 1.** Participant characteristics, by recruitment method.

|  |  | General population, n=3050 | GBMSM samples |  |  | p-value |
| --- | --- | --- | --- | --- | --- | --- |
|  |  |  | Savanta GBMSM, n=247 | Grindr, n=831 | Meta, n=1036 |  |
| Gender | Male (including trans man) | 1289 (42.4) | 247 (100.0) | 831 (100.0) | 1036 (100.0) | - |
|  | Female (including trans woman) | 1736 (57.1) | - | - | - | - |
|  | Non-binary | 17 (0.6) | - | - | - | - |
| Sexual orientation | Straight or heterosexual | 2795 (92.7) | - | - | - | - |
|  | Gay, lesbian, bisexual, or queer | 221 (7.3) | 247 (100.0) | 831 (100.0) | 1036 (100.0) | - |
| Age | Range 18 to 98 years | M=48.6,<br>SD=17.4 | M=47.1,<br>SD=16.5 | M=44.2,<br>SD=12.5 | M=47.6,<br>SD=11.9 | <0.001* |
| Region† | Midlands (East and West) | 522 (17.1) | 39 (15.8) | 73 (8.8) | 85 (8.2) | <0.001* |
|  | South (East, West, East of England) | 992 (32.5) | 70 (28.3) | 207 (24.9) | 279 (26.9) |  |
|  | North (East, West, Yorkshire and the Humber) | 774 (25.4) | 58 (23.5) | 119 (14.3) | 155 (15.0) |  |
|  | London | 308 (10.1) | 46 (18.6) | 195 (23.5) | 379 (36.6) |  |
|  | Devolved nations (Scotland, Wales, and Northern Ireland) | 454 (14.9) | 34 (13.8) | 95 (11.4) | 84 (8.1) |  |
|  | Not specified | 0 (0.0) | 0 (0.0) | 142 (17.1) | 54 (5.2) |  |
| Dependent child in household | No | 2075 (68.0) | 210 (85.0) | 796 (95.8) | 1014 (97.9) | <0.001* |
|  | Yes | 975 (32.0) | 37 (15.0) | 35 (4.2) | 22 (2.1) |  |
| Employment status | Not working | 1286 (42.6) | 93 (38.3) | 157 (19.0) | 201 (19.5) | <0.001* |
|  | Working | 1736 (57.4) | 150 (61.7) | 669 (81.0) | 831 (80.5) |  |
| Frontline health or social care worker | No | 2668 (88.3) | 224 (90.7) | 734 (88.6) | 909 (87.9) | 0.66 |
|  | Yes | 355 (11.7) | 23 (9.3) | 94 (11.4) | 125 (12.1) |  |
| Need to leave home for work | Do not need to leave home for work | 1946 (63.8) | 149 (60.3) | 371 (44.6) | 499 (48.2) | <0.001* |
|  | Need to leave home for work | 1104 (36.2) | 98 (39.7) | 460 (55.4) | 537 (51.8) |  |
| Education | GCSE/vocational/A-level/No formal qualifications | 2065 (67.7) | 159 (64.4) | 295 (35.5) | 253 (24.4) | <0.001* |
|  | Degree or higher (Bachelors, Masters, PhD) | 985 (32.3) | 88 (35.6) | 536 (64.5) | 783 (75.6) |  |
| Ethnicity | White British | 2649 (87.1) | 214 (86.6) | 629 (76.2) | 799 (78.0) | <0.001* |
|  | White other | 116 (3.8) | 18 (7.3) | 117 (14.2) | 163 (15.9) |  |
|  | Black, Asian, other minoritized ethnicity | 275 (9.0) | 15 (6.1) | 80 (9.7) | 63 (6.1) |  |
| Marital status | Not partnered | 1265 (41.6) | 141 (57.3) | 561 (68.3) | 486 (47.3) | <0.001* |
|  | Partnered | 1775 (58.4) | 105 (42.7) | 260 (31.7) | 541 (52.7) |  |
| Live alone | Live with someone else | 2387 (78.3) | 146 (59.1) | 473 (56.9) | 656 (63.3) | 0.02 |
|  | Live alone | 663 (21.7) | 101 (40.9) | 358 (43.1) | 380 (36.7) |  |
| Own chronic illness | None | 2207 (72.4) | 167 (67.6) | 619 (74.5) | 731 (70.6) | 0.05 |
|  | Present | 843 (27.6) | 80 (32.4) | 212 (25.5) | 305 (29.4) |  |
| Household member chronic illness | None | 2659 (87.2) | 226 (91.5) | 743 (89.4) | 921 (88.9) | 0.49 |
|  | Present | 391 (12.8) | 21 (8.5) | 88 (10.6) | 115 (11.1) |  |
| Ever taken PrEP for HIV | No | 2993 (98.6) | 216 (88.5) | 427 (51.5) | 547 (52.9) | <0.001* |
|  | Yes | 42 (1.4) | 28 (11.5) | 402 (48.5) | 487 (47.1) |  |
| Vaccinated for smallpox in 2022 | Not vaccinated | 2888 (94.7) | 221 (89.5) | 566 (68.1) | 614 (59.3) | <0.001* |
|  | Vaccinated | 162 (5.3) | 26 (10.5) | 265 (31.9) | 422 (40.7) |  |
| Index of multiple deprivation† | Deciles (1<sup>st</sup> = most deprived, 10<sup>th</sup> = least deprived) | M=4.5,<br>SD=2.9 | M=4.4,<br>SD=3.0 | N=689,<br>M=4.7,<br>SD=2.8 | N=982,<br>M=4.9,<br>SD=2.7 | <0.001* |
| Socio-economic grade | ABC1 | 1796 (58.9) | 169 (68.4) | 694 (83.5) | 944 (91.1) | <0.001* |
|  | C2DE | 1254 (41.1) | 78 (31.6) | 137 (16.5) | 92 (8.9) |  |
| Financial hardship† | 4 (lowest hardship) to 13 (most hardship) | N=2743,<br>M=6.1,<br>SD=2.3 | N=223,<br>M=5.9,<br>SD=2.4 | N=784,<br>M=5.2,<br>SD=1.8 | N=1004,<br>M=4.7,<br>SD=1.3 | <0.001* |

\* $p \leq 0.001$
†Using original (not imputed) data.

### Motivational messaging

There was no effect of motivational messaging on outcomes, except for in the sample recruited from Meta (see supplementary materials 4). In this sample, those receiving the motivational message were less likely to intend to self-isolate for 21 days (*t*(1034)= −2.81, *p*=0.005; motivational message, n=529, M=7.1, SD=2.3; control message, n=507, M=7.5, SD=2.2), share details of all recent sexual contacts (*t*(1029)= −2.05, *p*=0.04; motivational message, n=526, M=4.1, SD=1.2; control message, n=505, M=4.2, SD=1.1), and be vaccinated for smallpox if advised (*t*(612)= −2.21, *p*=0.03; motivational message, n=304, M=4.7, SD=0.9; control message, n=310, M=4.8, SD=0.6).

### Self-isolation

Rates of intended self-isolation were higher when imagining you were a case than a high-risk contact. Three-quarters of the general population sample intended to self-isolate for 21 days if they were to develop mpox (75.2%, 95% CI 73.7% to 76.7%, n=2294; Table 2). However, only 68.9% (95% CI 67.2% to 70.5%, n=2100) intended to self-isolate if they were to come into contact with a case. Intention to self-isolate was lower in Grindr and Meta samples.

**Table 2.** Main behavioural outcomes, by sample.

|  |  |  | N (% , 95% CI) |  |  |  | p-value |
| --- | --- | --- | --- | --- | --- | --- | --- |
| | | | General population, total $n=3050$ | Savanta GBMSM, total $n=247$ | Grindr, total $n=831$ | Meta, total $n=1036$ | |
| **Self-isolation for 21 days** | If “you have” mpox | Probably or definitely would not, or not sure | 756 (24.8, 23.3-26.3) | 53 (21.5, 16.3-26.6) | 246 (29.6, 26.5-32.7) | 275 (26.5, 23.9-29.2) | - |
|  |  | Probably or definitely would | 2294 (75.2, 73.7-76.7) | 194 (78.5, 73.4-83.7) | 585 (70.4, 67.3-73.5) | 761 (73.5, 70.8-76.1) |  |
|  | If “you have come into high-risk contact with someone who has” mpox | Probably or definitely would not, or not sure | 950 (31.1, 29.5-32.8) | 77 (31.2, 25.4-37.0) | 371 (44.6, 41.3-48.0) | 491 (47.4, 44.3-50.4) | - |
|  |  | Probably or definitely would | 2100 (68.9, 67.2-70.5) | 170 (68.8, 63.0-74.6) | 460 (55.4, 52-58.7) | 545 (52.6, 49.6-55.7) |  |
| | Sum of intention if case and high-risk contact | 2 (lowest intention) to 10 (highest intention) | $M=8.0$, $SD=2.2$ | $M=7.9$, $SD=2.3$ | $M=7.2$, $SD=2.4$ | $M=7.3$, $SD=2.3$ | <0.001* |
| **Help seeking** | Any action that involves contacting a healthcare professional (by phone or in person) | Would not seek help or would “wait and see” for a day or two to see if symptoms resolved | 1548 (50.8, 49.0-52.5) | 125 (50.6, 44.3-56.9) | 388 (46.7, 43.3-50.1) | 391 (37.7, 34.8-40.7) | <0.001* |
|  |  | Would seek help immediately | 1502 (49.2, 47.5-51.0) | 122 (49.4, 43.1-55.7) | 443 (53.3, 49.9-56.7) | 645 (62.3, 59.3-65.2) |  |
| **Sexual contact behaviour †** | Have sexual contact with others | Would not completely stop | 567 (22.8, 21.1-24.4) | 45 (20.8, 15.4-26.3) | 69 (8.7, 6.8-10.7) | 69 (7.0, 5.4-8.6) | <0.001* |
|  |  | Completely stop | 1923 (77.2, 75.6-78.9) | 171 (79.2, 73.7-84.6) | 721 (91.3, 89.3-93.2) | 922 (93.0, 91.4-94.6) |  |
| **Contact sharing** | Sexual contacts | Probably or definitely would not, or not sure | 697 (23.3, 21.7-24.8) | 60 (24.7, 19.2-30.2) | 167 (20.3, 17.5-23.0) | 204 (19.8, 17.4-22.2) | - |
|  |  | Probably or definitely would | 2300 (76.7, 75.2-78.3) | 183 (75.3, 69.8-80.8) | 656 (79.7, 77-82.5) | 827 (80.2, 77.8-82.6) |  |

|  | Scale | 1 (definitely would not) to 5 (definitely would) | N=2997, M=4.2, SD=1.2 | N=243, M=4.1, SD=1.2 | N=823, M=4.1, SD=1.2 | N=1031, M=4.2, SD=1.1 | 0.57 |
| --- | --- | --- | --- | --- | --- | --- | --- |
| **Vaccination intention** | Vaccinated in 2022 | No or not sure | 2888 (94.7, 93.9-95.5) | 221 (89.5, 85.6-93.3) | 566 (68.1, 64.9-71.3) | 614 (59.3, 56.3-62.3) | <0.001* |
|  |  | Yes | 162 (5.3, 4.5-6.1) | 26 (10.5, 6.7-14.4) | 265 (31.9, 28.7-35.1) | 422 (40.7, 37.7-43.7) |  |
|  | Vaccinated in 2022 or would be vaccinated if advised | Not vaccinated and probably or definitely would not be vaccinated or not sure | 786 (25.8, 24.2-27.3) | 37 (15.0, 10.5-19.5) | 54 (6.5, 4.8-8.2) | 38 (3.7, 2.5-4.8) | <0.001* |
|  |  | Vaccinated or probably or definitely would be vaccinated | 2264 (74.2, 72.7-75.8) | 210 (85.0, 80.5-89.5) | 777 (93.5, 91.8-95.2) | 998 (96.3, 95.2-97.5) |  |

\* $p \leq 0.001$
†Answers of “don’t know”, “prefer not to say” and “not applicable, I wouldn’t do this anyway” were coded as missing, therefore total *ns* are substantially lower (general population, *n*=2490; Savanta GBMSM, *n*=216; Grindr, *n*=790; Meta, *n*=991).

In the general population, intention to self-isolate was associated with: less financial hardship, being more worried about mpox, perceiving a bigger threat of mpox to people in the UK, greater perceived susceptibility and severity of mpox, and greater perceived social norms for self-isolation (Tables 3 and 4). Not intending to self-isolate was associated with agreeing that if you had mpox symptoms, you would not want to tell anyone as you did not want to self-isolate, believing the risks of mpox were being exaggerated, and that if you had to self-isolate due to mpox it would have a negative impact on your work (Table 4). In the Grindr sample, self-isolation intention was associated with greater perceived social norms (Table 4). Not intending to self-isolate was associated with agreeing that if you had mpox symptoms, you would not want to tell anyone as you did not want to self-isolate, and that if you had to self-isolate due to mpox it would have a negative impact on your work (Table 4).

**Table 3.** Associations between intending to self-isolate and socio-demographic characteristics and motivational message, by sample. A higher score indicates greater intention to self-isolate. Variables were entered into the linear regression model in blocks (block 1: socio-demographic variables and motivational message, block 2: psychological factors, block 3: isolation-specific beliefs). Results for block 3, using pooled estimates are reported.

| Participant characteristics | Level | General population |  | Grindr |  |
| --- | --- | --- | --- | --- | --- |
|  |  | *B* (95% CI) | *p*-value | *B* (95% CI) | *p*-value |
| Gender | Male (including trans man) | Ref | - | - | - |
|  | Female (including trans woman) | 0.15 (0.01 to 0.30) | 0.03 | - | - |
| Sexual orientation | Straight or heterosexual | Ref | - | - | - |
|  | Gay, lesbian, bisexual, or queer | 0.06 (-0.21 to 0.33) | 0.67 | - | - |
| Age | Range 18 to 98 years | 0.01 (0.00 to 0.01) | 0.06 | 0.00 (-0.01 to 0.02) | 0.66 |
|  | Quadratic term, (age – mean)<sup>2</sup> | 0.0002 (-0.0001 to 0.0005) | 0.17 | 0.001 (0.000 to 0.002) | 0.02 |
| Region | Midlands (East and West) | Ref | - | Ref | - |
|  | South (East, West, East of England) | -0.19 (-0.39 to 0.01) | 0.07 | 0.02 (-0.51 to 0.55) | 0.95 |
|  | North (East, West, Yorkshire and the Humber) | -0.07 (-0.28 to 0.13) | 0.49 | 0.06 (-0.53 to 0.64) | 0.85 |
|  | London | -0.11 (-0.39 to 0.16) | 0.42 | -0.01 (-0.59 to 0.57) | 0.98 |
|  | Devolved nations (Scotland, Wales, and Northern Ireland) | -0.04 (-0.30 to 0.22) | 0.78 | 0.48 (-0.20 to 1.16) | 0.17 |
| Dependent child in household | No | Ref | - | Ref | - |
|  | Yes | -0.15 (-0.32 to 0.03) | 0.10 | -0.42 (-1.13 to 0.29) | 0.25 |
| Employment status | Not working | Ref | - | Ref | - |
|  | Working | 0.09 (-0.12 to 0.31) | 0.39 | -0.13 (-0.61 to 0.34) | 0.58 |
| Frontline health or social care worker | No | Ref | - | Ref | - |
|  | Yes | 0.08 (-0.14 to 0.30) | 0.47 | 0.68 (0.24 to 1.13) | 0.003 |
| Need to leave home for work | Do not need to leave home for work | Ref | - | Ref | - |
|  | Need to leave home for work | -0.26 (-0.45 to -0.07) | 0.007 | 0.14 (-0.22 to 0.51) | 0.44 |
| Education | GCSE/vocational/A-level/No formal qualifications | Ref | - | Ref | - |
|  | Degree or higher (Bachelors, Masters, PhD) | 0.02 (-0.13 to 0.18) | 0.77 | -0.16 (-0.48 to 0.16) | 0.31 |
| Ethnicity | White British | Ref | - | Ref | - |
|  | White other | -0.23 (-0.59 to 0.13) | 0.21 | -0.33 (-0.75 to 0.09) | 0.12 |
|  | Black, Asian, other minoritized ethnicity | 0.11 (-0.15 to 0.37) | 0.40 | -0.03 (-0.52 to 0.46) | 0.91 |
| Marital status | Not partnered | Ref | - | Ref | - |
|  | Partnered | 0.15 (-0.04 to 0.34) | 0.12 | 0.13 (-0.24 to 0.49) | 0.50 |
| Live alone | Live with someone else | Ref | - | Ref | - |
|  | Live alone | 0.06 (-0.15 to 0.28) | 0.56 | -0.07 (-0.40 to 0.26) | 0.69 |
| Own chronic illness | None | Ref | - | Ref | - |
|  | Present | 0.21 (0.05 to 0.38) | 0.009 | 0.10 (-0.24 to 0.44) | 0.56 |
| Ever taken PrEP for HIV | No | - | - | Ref | - |
|  | Yes | - | - | -0.11 (-0.43 to 0.22) | 0.51 |
| Vaccinated for smallpox in 2022 | Not vaccinated | - | - | Ref | - |
|  | Vaccinated | - | - | 0.03 (-0.32 to 0.39) | 0.86 |
| Index of multiple deprivation | Deciles (1<sup>st</sup> = most deprived, 10<sup>th</sup> = least deprived) | -0.01 (-0.04 to 0.02) | 0.48 | 0.02 (-0.05 to 0.08) | 0.66 |
| Socio-economic grade | ABC1 | Ref | - | Ref | - |
|  | C2DE | -0.09 (-0.24 to 0.06) | 0.24 | -0.03 (-0.46 to 0.39) | 0.88 |
| Financial hardship | 4 (lowest hardship) to 13 (most hardship) | -0.08 (-0.12 to -0.05) | <0.001* | -0.02 (-0.11 to 0.07) | 0.70 |
| Total number of sexual partners (male and female) in last three weeks | 0 | Ref | - | - | - |
|  | 1 | -0.04 (-0.23 to 0.14) | 0.65 | - | - |
|  | 2 to 4 | -0.25 (-0.59 to 0.10) | 0.16 | - | - |
|  | 5 or more | -0.09 (-0.69 to 0.52) | 0.77 | - | - |
|  | Prefer not to say | -0.22 (-0.42 to -0.02) | 0.03 | - | - |
| Number of male sexual partners in last three weeks | 0 | - | - | Ref | - |
|  | 1 | - | - | -0.50 (-0.92 to -0.08) | 0.02 |
|  | 2 to 4 | - | - | -0.09 (-0.47 to 0.30) | 0.65 |
|  | 5 to 9 | - | - | -0.48 (-1.00 to 0.03) | 0.07 |
|  | 10 or more | - | - | -0.32 (-0.96 to 0.33) | 0.34 |
|  | Prefer not to say | - | - | 0.83 (0.12 to 1.53) | 0.02 |
| Motivational message | Perceived risk of illness and necessity and efficacy of the response | -0.06 (-0.25 to 0.12) | 0.50 | - | - |
|  | Perceived risk of illness and benefits of the response | -0.09 (-0.28 to 0.09) | 0.33 | - | - |
|  | Perceived risk of illness and low perceived costs of response | -0.19 (-0.38 to 0.00) | 0.05 | - | - |
|  | Control | Ref | - | - | - |
| Motivational message | All motivational components | - | - | -0.10 (-0.37 to 0.18) | 0.48 |
|  | Control | - | - | Ref | - |

\* $p \leq 0.001$

**Table 4.** Associations between intending to self-isolate and psychological factors and isolation-specific beliefs, by sample. A higher score indicates greater intention to self-isolate. Variables were entered into the linear regression model in blocks (block 1: socio-demographic variables and motivational message, block 2: psychological factors, block 3: isolation-specific beliefs). Results for block 3, using pooled estimates are reported.

| Factor | Level | General population<br>B (95% CI) | p-value | Grindr<br>B (95% CI) | p-value |
| --- | --- | --- | --- | --- | --- |
| Amount heard about mpox | I have not seen or heard anything (1) to I have seen or heard a lot (3) | 0.09 (-0.07 to 0.24) | 0.26 | -0.09 (-0.41 to 0.23) | 0.57 |
| Worry about mpox | Not at all worried (1) to extremely worried (4) | 0.26 (0.13 to 0.39) | <0.001* | 0.41 (0.12 to 0.71) | 0.006 |
| Perceived risk of mpox to oneself | No risk at all (1) to very high risk (5) | -0.05 (-0.17 to 0.06) | 0.39 | -0.25 (-0.46 to -0.04) | 0.02 |
| Perceived risk of mpox to people in UK | No risk at all (1) to very high risk (5) | 0.19 (0.07 to 0.31) | 0.001* | 0.030 (-0.20 to 0.25) | 0.82 |
| Perceived susceptibility and severity | Lowest (1) to highest (5) | 0.03 (0.19 to 0.42) | <0.001* | 0.12 (-0.13 to 0.37) | 0.36 |
| I am already immune to mpox | Strongly disagree, disagree, neither agree nor disagree, don't know | Ref | - | Ref | - |
|  | Strongly agree and agree | 0.27 (0.03 to 0.52) | 0.03 | -0.12 (-0.61 to 0.36) | 0.62 |
| People who catch mpox usually make a full recovery, even if they do not receive any treatment | Strongly disagree (1) to strongly agree (5) | -0.05 (-0.14 to 0.03) | 0.23 | -0.26 (-0.43 to -0.08) | 0.004 |
| My personal behaviour has an impact on how mpox spreads | Strongly disagree (1) to strongly agree (5) | 0.09 (0.03 to 0.15) | 0.004 | 0.08 (-0.06 to 0.22) | 0.26 |
| My life has been negatively affected by changes made in response to the mpox outbreak | Strongly disagree (1) to strongly agree (5) | -0.01 (-0.09 to 0.07) | 0.89 | -0.03 (-0.17 to 0.11) | 0.70 |
| The risks of mpox are being exaggerated | Strongly disagree (1) to strongly agree (5) | -0.19 (-0.27 to -0.11) | <0.001* | -0.07 (-0.24 to 0.09) | 0.39 |
| Mpox is only a risk to men who are gay, bisexual or have sex with men | Strongly disagree (1) to strongly agree (5) | -0.02 (-0.08 to 0.05) | 0.58 | -0.17 (-0.32 to -0.02) | 0.03 |
| Perceived knowledge | Lowest (0) to highest (3) | -0.01 (-0.08 to 0.07) | 0.88 | -0.18 (-0.37 to 0.02) | 0.08 |
| Knowledge of mpox symptoms | Identified no symptoms (0) to identified four symptoms (4) | 0.08 (0.03 to 0.14) | 0.003 | 0.06 (-0.06 to 0.19) | 0.31 |
| Knowledge of mpox transmission | Lowest (0) to highest (6) | 0.02 (-0.03 to 0.07) | 0.42 | 0.04 (-0.10 to 0.17) | 0.59 |
| If I had mpox symptoms, I wouldn't want to tell anyone as I don't want to self-isolate | Strongly disagree (1) to strongly agree (5) | -0.66 (-0.73 to -0.60) | <0.001* | -1.01 (-1.15 to -0.87) | <0.001* |
| Most people would self-isolate if they were told to | Strongly disagree (1) to strongly agree (5) | 0.34 (0.28 to 0.41) | <0.001* | 0.41 (0.28 to 0.54) | <0.001* |
| If I had to self-isolate because I had tested positive for mpox...it would have a negative impact on my work | Strongly disagree (1) to strongly agree (5) | -0.15 (-0.21 to -0.09) | <0.001* | -0.34 (-0.46 to -0.21) | <0.001* |

\* $p \leq 0.001$

### Help seeking

Approximately half of participants in the general population, Savanta GBMSM, and Grindr samples indicated that they would seek help immediately (49.2% to 53.3%; Table 2). Intention to seek help immediately was higher in the Meta sample (62.3%, 95% CI 59.3 to 65.2, n=645).

In the general population, intention to seek help immediately was associated with being older (aOR 1.012, 95% CI 1.006 to 1.019, *p*<0.001), disagreeing that the risks of mpox are being exaggerated (aOR 0.83, 95% CI 0.76 to 0.91, *p*<0.001), and being willing to contact a sexual health clinic if you thought you had mpox symptoms or had been in contact with a case (aOR 1.25, 95% CI 1.16 to 1.34, *p*<0.001; supplementary materials 5). In the Grindr sample, intention to seek help immediately was associated with being willing to contact a sexual health clinic if you thought you had mpox symptoms or had been in contact with a case (aOR 1.57, 95% CI 1.29 to 1.91, *p*<0.001; supplementary materials 5).

### Sexual contact behaviour when symptomatic

In the general population and Savanta GBMSM, 77.2% (95% CI 75.6% to 78.9%, n=1923) and 79.2% (73.7% to 84.6%, n=171) intended to completely stop sexual contact if they were to develop an unexplained rash with blisters and learn that they had come into contact with a mpox case (Table 2). Rates of intending to completely stop sexual contact were significantly higher in Grindr (91.3%, 95% CI 89.3% to 93.2%, n=721) and Meta samples (93.0%, 95% CI 91.4% to 94.6%, n=922). The number of days that participants would wait before resuming sexual contact from the start of their symptoms was also higher in Grindr and Meta samples than in general population and Savanta GBMSM samples (*F*(3, 3900)=29.0, *p*<0.001; general population: n=2021, M=15.7, SD=15.3; Savanta GBMSM: n=190, M=16.2, SD=14.4; Grindr: n=746, M=20.4, SD=15.8; Meta: n=947, M=20.0, SD=12.3).

In the general population, intending to completely stop any sexual contact if symptomatic was associated with being female, older, less financial hardship, and being more knowledgeable about mpox transmission (Tables 5 and 6). Not intending to completely stop sexual behaviour was associated with preferring not to say how many recent sexual partners you had had, thinking that you were already immune to mpox, that your life had been negatively affected by changes made in response to the mpox outbreak, and that the risks of mpox were being exaggerated (Tables 5 and 6). No associations reached significance (Bonferroni corrected) in the Grindr sample.

**Table 5.** Associations between intending to completely stop any sexual contact and socio-demographic characteristics and motivational message, by sample. Variables were entered into the logistic regression model in blocks (block 1: socio-demographic variables and motivational message, block 2: psychological factors). Results for block 2, using pooled estimates are reported.

| Participant characteristics | Level | General population |  | Grindr |  |
| --- | --- | --- | --- | --- | --- |
|  |  | aOR (95% CI) | p-value | aOR (95% CI) | p-value |
| Gender | Male (including trans man) | Ref | - | - | - |
|  | Female (including trans woman) | 1.90 (1.48 to 2.45) | <0.001* | - | - |
| Sexual orientation | Straight or heterosexual | Ref | - | - | - |
|  | Gay, lesbian, bisexual, or queer | 0.91 (0.58 to 1.41) | 0.67 | - | - |
| Age | Range 18 to 98 years | 1.04 (1.03 to 1.05) | <0.001* | 1.01 (0.98 to 1.04) | 0.53 |
|  | Quadratic term, (age – mean)<sup>2</sup> | 1.000 (0.999 to 1.001) | 0.87 | 1.000 (0.998 to 1.001) | 0.71 |
| Region | Midlands (East and West) | Ref | - | Ref | - |
|  | South (East, West, East of England) | 1.08 (0.76 to 1.54) | 0.67 | 1.18 (0.38 to 3.66) | 0.77 |
|  | North (East, West, Yorkshire and the Humber) | 1.14 (0.79 to 1.66) | 0.49 | 1.48 (0.35 to 6.21) | 0.58 |
|  | London | 1.17 (0.73 to 1.86) | 0.52 | 0.94 (0.27 to 3.25) | 0.92 |
|  | Devolved nations (Scotland, Wales, and Northern Ireland) | 1.26 (0.79 to 2.01) | 0.33 | 1.26 (0.31 to 5.15) | 0.74 |
| Dependent child in household | No | Ref | - | Ref | - |
|  | Yes | 0.97 (0.72 to 1.29) | 0.82 | 0.37 (0.11 to 1.28) | 0.12 |
| Employment status | Not working | Ref | - | Ref | - |
|  | Working | 1.22 (0.90 to 1.66) | 0.20 | 0.92 (0.39 to 2.18) | 0.85 |
| Frontline health or social care worker | No | Ref | - | Ref | - |
|  | Yes | 0.97 (0.67 to 1.39) | 0.86 | 1.77 (0.67 to 4.65) | 0.25 |
| Education | GCSE/vocational/A-level/No formal qualifications | Ref | - | Ref | - |
|  | Degree or higher (Bachelors, Masters, PhD) | 0.96 (0.73 to 1.27) | 0.79 | 1.87 (0.97 to 3.59) | 0.06 |
| Ethnicity | White British | Ref | - | Ref | - |
|  | White other | 0.47 (0.26 to 0.84) | 0.01 | 0.84 (0.35 to 2.06) | 0.71 |
|  | Black, Asian, other minoritized ethnicity | 0.85 (0.56 to 1.27) | 0.42 | 0.36 (0.15 to 0.85) | 0.02 |
| Marital status | Not partnered | Ref | - | Ref | - |
|  | Partnered | 0.90 (0.65 to 1.24) | 0.50 | 1.11 (0.51 to 2.41) | 0.79 |
| Live alone | Live with someone else | Ref | - | Ref | - |
|  | Live alone | 1.16 (0.77 to 1.73) | 0.48 | 1.12 (0.56 to 2.23) | 0.75 |
| Own chronic illness | None | Ref | - | Ref | - |
|  | Present | 0.97 (0.72 to 1.31) | 0.85 | 1.10 (0.53 to 2.27) | 0.80 |
| Ever taken PrEP for HIV | No | - | - | Ref | - |
|  | Yes | - | - | 1.36 (0.69 to 2.71) | 0.37 |
| Vaccinated for smallpox in 2022 | Not vaccinated | - | - | Ref | - |
|  | Vaccinated | - | - | 0.71 (0.33 to 1.50) | 0.37 |
| Index of multiple deprivation | Deciles (1<sup>st</sup> = most deprived, 10<sup>th</sup> = least deprived) | 1.00 (0.96 to 1.06) | 0.85 | 1.01 (0.86 to 1.19) | 0.88 |
| Socio-economic grade | ABC1 | Ref | - | Ref | - |
|  | C2DE | 1.13 (0.86 to 1.49) | 0.39 | 4.15 (1.45 to 11.85) | 0.008 |
| Financial hardship | 4 (lowest hardship) to 13 (most hardship) | 0.85 (0.80 to 0.90) | <0.001* | 0.89 (0.74 to 1.07) | 0.21 |
| Total number of sexual partners (male and female) in last three weeks | 0 | Ref | - | - | - |
|  | 1 | 0.83 (0.59 to 1.18) | 0.30 | - | - |
|  | 2 to 4 | 0.68 (0.39 to 1.18) | 0.17 | - | - |
|  | 5 or more | 0.37 (0.14 to 0.96) | 0.04 | - | - |
|  | Prefer not to say | 0.45 (0.32 to 0.64) | <0.001* | - | - |
| Number of male sexual partners in last three weeks | 0 | - | - | Ref | - |
|  | 1 | - | - | 0.52 (0.18 to 1.46) | 0.21 |
|  | 2 to 4 | - | - | 0.38 (0.14 to 0.98) | 0.05 |
|  | 5 to 9 | - | - | 0.21 (0.07 to 0.65) | 0.007 |
|  | 10 or more | - | - | 0.20 (0.06 to 0.72) | 0.01 |
|  | Prefer not to say | - | - | 0.30 (0.08 to 1.11) | 0.07 |
| Motivational message | Perceived risk of illness and necessity and efficacy of the response | 1.37 (0.97 to 1.93) | 0.08 | - | - |
|  | Perceived risk of illness and benefits of the response | 1.07 (0.76 to 1.49) | 0.70 | - | - |
|  | Perceived risk of illness and low perceived costs of response | 1.10 (0.79 to 1.55) | 0.57 | - | - |
|  | Control | Ref | - | - | - |
| Motivational message | All motivational components | - | - | 1.43 (0.80 to 2.54) | 0.23 |
|  | Control | - | - | Ref | - |

\* $p \leq 0.001$

**Table 6.** Associations between intending to completely stop any sexual contact and psychological and contextual factors, by sample. Variables were entered into the logistic regression model in blocks (block 1: socio-demographic variables and motivational message, block 2: psychological factors). Results for block 2, using pooled estimates are reported.

| Factor | Level | General population<br>aOR (95% CI) | p-value | Grindr<br>aOR (95% CI) | p-value |
| --- | --- | --- | --- | --- | --- |
| Amount heard about mpox | I have not seen or heard anything (1) to I have seen or heard a lot (3) | 1.42 (1.08 to 1.86) | 0.01 | 1.38 (0.70 to 2.72) | 0.35 |
| Worry about mpox | Not at all worried (1) to extremely worried (4) | 1.11 (0.88 to 1.40) | 0.36 | 1.51 (0.82 to 2.77) | 0.19 |
| Perceived risk of mpox to oneself | No risk at all (1) to very high risk (5) | 0.83 (0.68 to 1.02) | 0.08 | 0.93 (0.60 to 1.44) | 0.75 |
| Perceived risk of mpox to people in UK | No risk at all (1) to very high risk (5) | 1.10 (0.90 to 1.34) | 0.35 | 0.99 (0.64 to 1.53) | 0.95 |
| Perceived susceptibility and severity | Lowest (1) to highest (5) | 1.27 (1.03 to 1.56) | 0.03 | 1.18 (0.72 to 1.94) | 0.51 |
| I am already immune to mpox | Strongly disagree, disagree, neither agree nor disagree, don't know | Ref | - | Ref | - |
|  | Strongly agree and agree | 0.37 (0.26 to 0.53) | <0.001* | 0.78 (0.32 to 1.90) | 0.59 |
| People who catch mpox usually make a full recovery, even if they do not receive any treatment | Strongly disagree (1) to strongly agree (5) | 0.87 (0.74 to 1.01) | 0.08 | 0.66 (0.45 to 0.97) | 0.04 |
| My personal behaviour has an impact on how mpox spreads | Strongly disagree (1) to strongly agree (5) | 1.03 (0.92 to 1.16) | 0.57 | 1.42 (1.08 to 1.87) | 0.01 |
| My life has been negatively affected by changes made in response to the mpox outbreak | Strongly disagree (1) to strongly agree (5) | 0.70 (0.62 to 0.80) | <0.001* | 0.73 (0.55 to 0.97) | 0.03 |
| The risks of mpox are being exaggerated | Strongly disagree (1) to strongly agree (5) | 0.77 (0.67 to 0.88) | <0.001* | 0.81 (0.58 to 1.11) | 0.18 |
| mpox is only a risk to men who are gay, bisexual or have sex with men | Strongly disagree (1) to strongly agree (5) | 0.87 (0.77 to 0.97) | 0.02 | 0.77 (0.58 to 1.03) | 0.08 |
| Perceived knowledge | Lowest (0) to highest (3) | 1.00 (0.88 to 1.15) | 0.94 | 0.86 (0.58 to 1.28) | 0.46 |
| Knowledge of mpox symptoms | Identified no symptoms (0) to identified four symptoms (4) | 1.14 (1.03 to 1.26) | 0.01 | 1.29 (1.00 to 1.66) | 0.05 |
| Knowledge of mpox transmission | Lowest (0) to highest (6) | 1.18 (1.08 to 1.29) | <0.001* | 1.31 (1.00 to 1.73) | 0.05 |

\* $p \leq 0.001$

### Sharing details of contacts

There was no difference in intention to share details of all sexual partners in the last seven days between samples (Table 2), with 75.3% to 80.2% saying that they probably or definitely would.

In the general population sample, intention to share details of every sexual contact in the last seven days was associated with being female, older, less financial hardship, higher perceived susceptibility to and severity of mpox, and higher knowledge about modes of mpox transmission (Tables 7 and 8). Not intending to share details of every recent sexual contact was associated with preferring not to say how many recent sexual partners you had had, thinking that your life had been negatively affected by changes made in response to the mpox outbreak, and thinking that the risks of mpox had been exaggerated (Tables 7 and 8). No associations reached our threshold for significance in the Grindr sample.

**Table 7.**
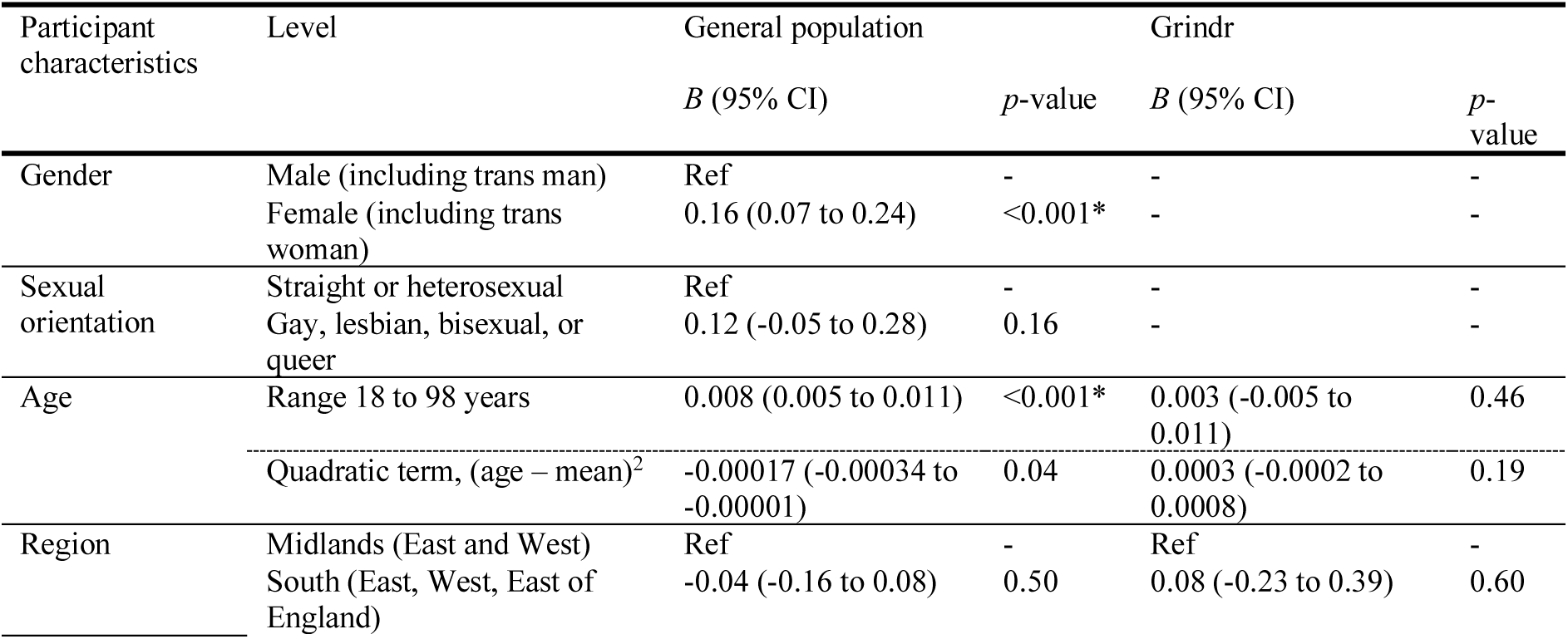
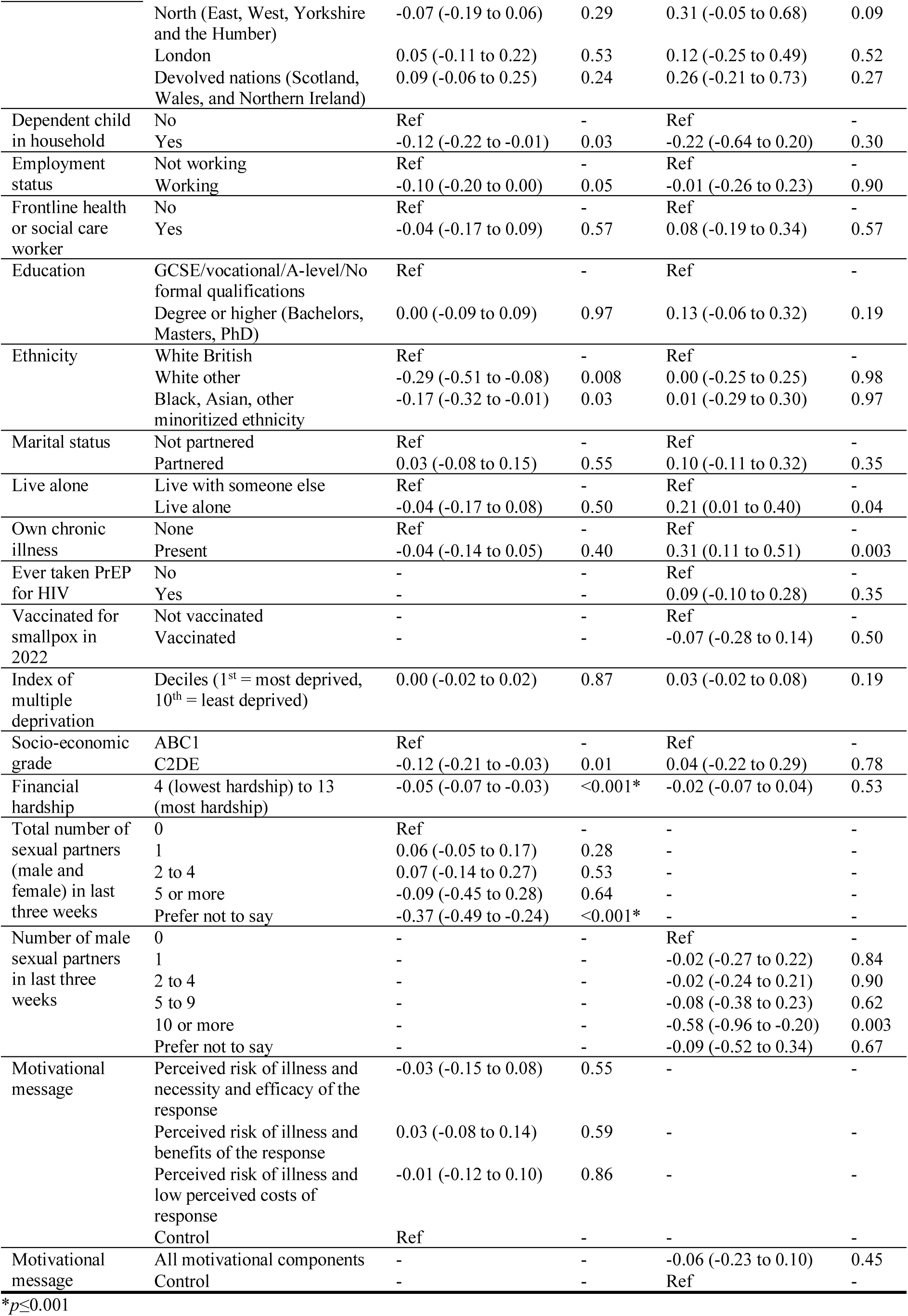
Associations between intending to share details of every sexual contact in the last seven days and socio-demographic characteristics and motivational message, by sample. A higher score indicates greater intention to share details. Variables were entered into the linear regression model in blocks (block 1: socio-demographic variables and motivational message, block 2: psychological factors). Results for block 2, using pooled estimates are reported.

| Participant characteristics | Level | General population |  | Grindr |  |
| --- | --- | --- | --- | --- | --- |
|  |  | *B* (95% CI) | *p*-value | *B* (95% CI) | *p*-value |
| Gender | Male (including trans man) | Ref | - | - | - |
|  | Female (including trans woman) | 0.16 (0.07 to 0.24) | <0.001* | - | - |
| Sexual orientation | Straight or heterosexual | Ref | - | - | - |
|  | Gay, lesbian, bisexual, or queer | 0.12 (-0.05 to 0.28) | 0.16 | - | - |
| Age | Range 18 to 98 years | 0.008 (0.005 to 0.011) | <0.001* | 0.003 (-0.005 to 0.011) | 0.46 |
|  | Quadratic term, (age – mean)<sup>2</sup> | -0.00017 (-0.00034 to -0.00001) | 0.04 | 0.0003 (-0.0002 to 0.0008) | 0.19 |
| Region | Midlands (East and West) | Ref | - | Ref | - |
|  | South (East, West, East of England) | -0.04 (-0.16 to 0.08) | 0.50 | 0.08 (-0.23 to 0.39) | 0.60 |
|  | North (East, West, Yorkshire and the Humber) | -0.07 (-0.19 to 0.06) | 0.29 | 0.31 (-0.05 to 0.68) | 0.09 |
|  | London | 0.05 (-0.11 to 0.22) | 0.53 | 0.12 (-0.25 to 0.49) | 0.52 |
|  | Devolved nations (Scotland, Wales, and Northern Ireland) | 0.09 (-0.06 to 0.25) | 0.24 | 0.26 (-0.21 to 0.73) | 0.27 |
| Dependent child in household | No | Ref | - | Ref | - |
|  | Yes | -0.12 (-0.22 to -0.01) | 0.03 | -0.22 (-0.64 to 0.20) | 0.30 |
| Employment status | Not working | Ref | - | Ref | - |
|  | Working | -0.10 (-0.20 to 0.00) | 0.05 | -0.01 (-0.26 to 0.23) | 0.90 |
| Frontline health or social care worker | No | Ref | - | Ref | - |
|  | Yes | -0.04 (-0.17 to 0.09) | 0.57 | 0.08 (-0.19 to 0.34) | 0.57 |
| Education | GCSE/vocational/A-level/No formal qualifications | Ref | - | Ref | - |
|  | Degree or higher (Bachelors, Masters, PhD) | 0.00 (-0.09 to 0.09) | 0.97 | 0.13 (-0.06 to 0.32) | 0.19 |
| Ethnicity | White British | Ref | - | Ref | - |
|  | White other | -0.29 (-0.51 to -0.08) | 0.008 | 0.00 (-0.25 to 0.25) | 0.98 |
|  | Black, Asian, other minoritized ethnicity | -0.17 (-0.32 to -0.01) | 0.03 | 0.01 (-0.29 to 0.30) | 0.97 |
| Marital status | Not partnered | Ref | - | Ref | - |
|  | Partnered | 0.03 (-0.08 to 0.15) | 0.55 | 0.10 (-0.11 to 0.32) | 0.35 |
| Live alone | Live with someone else | Ref | - | Ref | - |
|  | Live alone | -0.04 (-0.17 to 0.08) | 0.50 | 0.21 (0.01 to 0.40) | 0.04 |
| Own chronic illness | None | Ref | - | Ref | - |
|  | Present | -0.04 (-0.14 to 0.05) | 0.40 | 0.31 (0.11 to 0.51) | 0.003 |
| Ever taken PrEP for HIV | No | - | - | Ref | - |
|  | Yes | - | - | 0.09 (-0.10 to 0.28) | 0.35 |
| Vaccinated for smallpox in 2022 | Not vaccinated | - | - | Ref | - |
|  | Vaccinated | - | - | -0.07 (-0.28 to 0.14) | 0.50 |
| Index of multiple deprivation | Deciles (1<sup>st</sup> = most deprived, 10<sup>th</sup> = least deprived) | 0.00 (-0.02 to 0.02) | 0.87 | 0.03 (-0.02 to 0.08) | 0.19 |
| Socio-economic grade | ABC1 | Ref | - | Ref | - |
|  | C2DE | -0.12 (-0.21 to -0.03) | 0.01 | 0.04 (-0.22 to 0.29) | 0.78 |
| Financial hardship | 4 (lowest hardship) to 13 (most hardship) | -0.05 (-0.07 to -0.03) | <0.001* | -0.02 (-0.07 to 0.04) | 0.53 |
| Total number of sexual partners (male and female) in last three weeks | 0 | Ref | - | - | - |
|  | 1 | 0.06 (-0.05 to 0.17) | 0.28 | - | - |
|  | 2 to 4 | 0.07 (-0.14 to 0.27) | 0.53 | - | - |
|  | 5 or more | -0.09 (-0.45 to 0.28) | 0.64 | - | - |
|  | Prefer not to say | -0.37 (-0.49 to -0.24) | <0.001* | - | - |
| Number of male sexual partners in last three weeks | 0 | - | - | Ref | - |
|  | 1 | - | - | -0.02 (-0.27 to 0.22) | 0.84 |
|  | 2 to 4 | - | - | -0.02 (-0.24 to 0.21) | 0.90 |
|  | 5 to 9 | - | - | -0.08 (-0.38 to 0.23) | 0.62 |
|  | 10 or more | - | - | -0.58 (-0.96 to -0.20) | 0.003 |
|  | Prefer not to say | - | - | -0.09 (-0.52 to 0.34) | 0.67 |
| Motivational message | Perceived risk of illness and necessity and efficacy of the response | -0.03 (-0.15 to 0.08) | 0.55 | - | - |
|  | Perceived risk of illness and benefits of the response | 0.03 (-0.08 to 0.14) | 0.59 | - | - |
|  | Perceived risk of illness and low perceived costs of response | -0.01 (-0.12 to 0.10) | 0.86 | - | - |
|  | Control | Ref | - | - | - |
| Motivational message | All motivational components | - | - | -0.06 (-0.23 to 0.10) | 0.45 |
|  | Control | - | - | Ref | - |

\* $p \leq 0.001$

**Table 8.** Associations between intending to share details of every sexual contact in the last seven days and psychological and contextual factors, by sample. A higher score indicates greater intention to share details. Variables were entered into the linear regression model in blocks (block 1: socio-demographic variables and motivational message, block 2: psychological factors). Results for block 2, using pooled estimates are reported.

| Factor | Level | General population<br>B (95% CI) | p-value | Grindr<br>B (95% CI) | p-value |
| --- | --- | --- | --- | --- | --- |
| Amount heard about mpox | I have not seen or heard anything (1) to I have seen or heard a lot (3) | -0.04 (-0.13 to 0.05) | 0.43 | 0.01 (-0.18 to 0.20) | 0.92 |
| Worry about mpox | Not at all worried (1) to extremely worried (4) | 0.08 (0.00 to 0.16) | 0.06 | 0.19 (0.01 to 0.36) | 0.03 |
| Perceived risk of mpox to oneself | No risk at all (1) to very high risk (5) | -0.04 (-0.11 to 0.03) | 0.27 | -0.06 (-0.18 to 0.06) | 0.34 |
| Perceived risk of mpox to people in UK | No risk at all (1) to very high risk (5) | 0.07 (0.00 to 0.14) | 0.06 | -0.01 (-0.14 to 0.12) | 0.85 |
| Perceived susceptibility and severity | Lowest (1) to highest (5) | 0.12 (0.05 to 0.19) | <0.001* | -0.09 (-0.24 to 0.06) | 0.24 |
| I am already immune to mpox | Strongly disagree, disagree, neither agree nor disagree, don't know | Ref | - | Ref | - |
|  | Strongly agree and agree | -0.18 (-0.32 to -0.04) | 0.01 | -0.09 (-0.37 to 0.20) | 0.55 |
| People who catch mpox usually make a full recovery, even if they do not receive any treatment | Strongly disagree (1) to strongly agree (5) | -0.01 (-0.06 to 0.04) | 0.71 | -0.06 (-0.17 to 0.04) | 0.23 |
| My personal behaviour has an impact on how mpox spreads | Strongly disagree (1) to strongly agree (5) | 0.04 (0.01 to 0.08) | 0.03 | 0.02 (-0.06 to 0.10) | 0.64 |
| My life has been negatively affected by changes made in response to the mpox outbreak | Strongly disagree (1) to strongly agree (5) | -0.10 (-0.15 to -0.06) | <0.001* | -0.03 (-0.11 to 0.06) | 0.56 |
| The risks of mpox are being exaggerated | Strongly disagree (1) to strongly agree (5) | -0.11 (-0.15 to -0.06) | <0.001* | -0.15 (-0.25 to -0.05) | 0.004 |
| Mpox is only a risk to men who are gay, bisexual or have sex with men | Strongly disagree (1) to strongly agree (5) | -0.02 (-0.06 to 0.02) | 0.29 | -0.07 (-0.16 to 0.01) | 0.10 |
| Perceived knowledge | Lowest (0) to highest (3) | 0.05 (0.00 to 0.09) | 0.04 | 0.02 (-0.10 to 0.13) | 0.78 |
| Knowledge of mpox symptoms | Identified no symptoms (0) to identified four symptoms (4) | 0.05 (0.02 to 0.08) | 0.004 | 0.05 (-0.02 to 0.13) | 0.14 |
| Knowledge of mpox transmission | Lowest (0) to highest (6) | 0.05 (0.02 to 0.08) | <0.001* | 0.05 (-0.03 to 0.13) | 0.22 |

\* $p \leq 0.001$

### Vaccination

Few people had been vaccinated for smallpox in 2022 in the general population sample (5.3%, 95% CI 4.5% to 6.1%, n=162; Table 2). This was significantly higher in GBMSM samples (10.5% to 40.7%). In a measure of actual and intended vaccination, 96.3% (95% CI 95.2% to 97.5%, n=998) of the Meta sample and 93.5% (95% CI 91.8% to 95.2%, n=777) of the Grindr sample were vaccinated or intended to be vaccinated for smallpox if advised. Rates were significantly lower in the general population and Savanta GBMSM samples.

In the general population, being vaccinated for smallpox in 2022 or intending to be vaccinated if offered a vaccine was associated with being older (aOR 1.015, 95% CI 1.007 to 1.023, *p*<0.001), more worried about mpox (1.49, 95% CI 1.24 to 1.80, *p*<0.001), and perceiving a higher susceptibility to and severity of mpox (1.31, 95% CI 1.11 to 1.55, *p*<0.001; supplementary materials 6). Not intending to be vaccinated was associated with needing to leave home for work (aOR 0.56, 95% CI 0.43 to 0.73, *p*<0.001) and agreeing that the risks of mpox were being exaggerated (aOR 0.76, 95% CI 0.68 to 0.85, *p*<0.001; supplementary materials 6).

In the Grindr sample, vaccination uptake was associated with ever having taken PrEP for HIV (aOR 8.95, 95% CI 5.61 to 14.28, *p*<0.001) and agreeing that you are already immune to mpox (aOR 8.83, 95% CI 4.52 to 17.22, *p*<0.001; supplementary materials 6). When including vaccination intention, being vaccinated for smallpox in 2022 or intending to be vaccinated if offered a vaccine was associated with agreeing that if you got a smallpox vaccine, you would be protected against mpox (aOR 3.25, 95% CI 1.97 to 5.35, *p*<0.001; supplementary materials 6).

## DISCUSSION

We investigated mpox attitudes, beliefs, and intended behaviours in a general population sample and three GBMSM samples. Samples differed by socio-demographic characteristics. This was reflected in intended behaviours, with GBMSM recruited from Grindr or Meta being more likely to intend to seek help immediately for mpox symptoms, completely stop sexual contact when symptomatic, and be vaccinated for smallpox if advised. High rates of smallpox vaccination in these samples perhaps reflects that they were mostly educated and working, and thus more likely to be health literate and engaged with services. This was also reflected in knowledge about mpox, with Grindr and Meta samples being more likely to correctly identify mpox transmission modes and the symptoms of mpox. Within the general population, rates of “don’t know” answers were high, with 18.4% selecting “don’t know” when asked what the main symptoms of mpox were, and 18.7% to 24.2% selecting “don’t know” for transmission modes (except for having sex with someone who has mpox, where 13.9% selected “don’t know”). This is similar to other surveys of the general population, where 24% of people were not sure if mpox usually spreads by close contact with an infected person.(41)

There were no effects of motivational messaging on our behavioural outcomes, except for in the Meta sample for intention to self-isolate, share details of all recent sexual contacts, and be vaccinated for smallpox if advised. For these outcomes, intention to engage with protective behaviours was higher in the control group. In practice, however, intentions were very similar (means changed by 0.1 to 0.4) and this is unlikely to represent a meaningful difference. Another study investigating gonorrhoea reinfection in young adults has also found that those in the control group (who received monthly texts reminding them to update their contact details) were less likely to be reinfected than those in the intervention group (who received a series of texts that were educational and used behaviour change techniques).(42) These messages were specifically designed to decrease stigma, and partner numbers were higher in the intervention arm. A systematic review of intervention communications during the COVID-19 pandemic found that messages about the personal and collective benefits of vaccination had mixed effects on vaccination intention, with some suggestions that this approach may be more effective in more strongly hesitant individuals.(28)

Generally, higher intentions were seen in the Grindr and Meta samples, except for self-isolation. However, rates should be interpreted with caution, as intended behaviour does not always translate to enacted behaviour.(43) This was shown to be the case for intended and actual engagement with the UK contact tracing programme during the COVID-19 pandemic.(11) Rates of having already been vaccinated for smallpox in 2022 were especially high (32% Grindr, 41% Meta), again suggesting our sample may have been particularly interested in the topic of the survey, especially given issues with access to vaccination.(44) In the UK, the public health agency (UK Health Security Agency) has been working in conjunction with community-based organisations and charities to raise awareness of mpox and how to prevent the transmission in GBMSM as the population most affected. Higher rates of knowledge and behavioural intentions suggest that this targeted messaging has been effective in increasing knowledge and driving protective behaviours. Evidence suggests that while one-off sexual encounters may make up only a minority of sexual interactions, they could account for a large proportion of mpox transmission.(45, 46) Furthermore, if encounters are anonymous, this will affect contact tracing efforts. Therefore, very high rates of intending to completely stop sexual contact with others if symptomatic (91% to 93% in Grindr and Meta samples respectively) are encouraging.

It is notable that 19% (Grindr, Meta) to 30% of people (general population, Savanta GBMSM) agreed that is it best to avoid physical contact with GBMSM because of the mpox outbreak, perhaps also reflecting a change in behaviour in this population. Recent decreases in some sexually transmitted infections in GBMSM, and the decline in mpox transmission in England, suggest this may be the case.(6)

Within the general population, women, people who were older, and those with lower financial hardship were more likely to intend to carry out protective behaviours. This pattern was also widely seen during the COVID-19 pandemic (11, 47, 48) and previous outbreaks.(49) This highlights the importance of considering health equity issues to ensure effective outbreak control. Offering financial support for protective behaviours, especially those that may affect people’s ability to earn an income such as self-isolation, is likely to increase engagement, especially for those in lower income settings. Few socio-demographic characteristics were associated with outcomes in the Grindr sample. This could be because the Grindr sample differed from the general population sample on key characteristics (more educated, higher socio-economic grade, less financial hardship), or be a function of the way analyses were conducted with all variables being entered into a single regression model. The Grindr sample, being smaller, also had about half the statistical power of the general population sample.

Intention to engage in protective behaviours in the general population was also associated with psychological factors such as greater worry about mpox, perceived risk of mpox to others (but there was little evidence of an association with perceived risk to oneself), perceived susceptibility and severity of mpox, and greater knowledge about transmission. This pattern of results was also seen during the COVID-19 and aH1N1 influenza pandemics.(12, 47, 50–52) COVID-19 vaccination intention was also associated with perceived risk to others, but not to oneself.(33) Few associations were significant in the Grindr sample. In addition to the decreased statistical power, this may also reflect the different context and experience that participants in this group had. While for our general population sample the questions about mpox risk were somewhat hypothetical, the Grindr sample were GBMSM and thus more likely to be impacted by mpox (as illustrated by the high vaccination rates). Factors relating to their personal experience of the outbreak may have superseeded any effect of the psychological variables that we assessed. Taken together, these results suggest that clear communications about the level of risk of infection may encourage people to enact protective behaviours when appropriate.

Agreeing that the risks of mpox were being exaggerated was associated with lower intention to engage with protective behaviours. This was also the case in previous pandemics.(47, 53) Other psychological factors associated with decreased intention to engage with protective behaviours were believing that you were already immune to mpox and that your life had been negatively affected by changes made in response to mpox. Attending an important event was one of the reasons given for breaking self-isolation during the aH1N1 pandemic.(54) The peak of the mpox outbreak in the UK occurred in July 2022, a period coinciding with the summer holiday and festival season, including Pride events. It is important to be conscious of asking people to self-isolate or quarantine over periods encompassing key events that may be happening.

Behaviour-specific beliefs were also associated with intentions. For self-isolation, greater intention was associated with greater social norms (thinking that others would also self-isolate). Greater social norms were associated with fewer outings during lockdown (14) and increased vaccination uptake and intention (in oneself and one’s child) (15, 55) during the COVID-19 pandemic. Social norms were also associated with enacting protective behaviours, including self-isolation, during previous outbreaks.(49, 56) Lower self-isolation intention was associated with thinking that self-isolation would have a negative impact on your work. In this study, as in previous research,(11) the main reasons for not being able to self-isolate were needing to go out for essentials (food / medicines), for a walk or some other exercise, and for work. In the UK, people with suspected mpox were directed to call sexual health services, likely explaining this association. Some people may have found this stigmatising as mpox is not a sexually transmitted infection.(57) Stigma surrounding having other sexually transmitted infections is associated with not seeking help.(58) In our study, immediate help seeking was associated with being willing to contact a sexual health clinic. These findings suggest that widening recommended points of contact with health services for suspected mpox cases or contacts may be beneficial. Taken together, results suggest that communications emphasising that others are also engaging with the protective behaviour and recommending non-stigmatising routes into health systems may improve engagement with self-isolation and help seeking.

In the Grindr sample, having been vaccinated for smallpox in 2022 was associated with having ever taken PrEP for HIV. This is probably due to how the vaccine was rolled out in the UK. GBMSM identified by sexual health services as being at highest risk of exposure – using markers similar to those used to assess eligibility for PrEP – were invited to be vaccinated.(59) Being vaccinated was also strongly associated with thinking that you were already immune to mpox in this survey. As this is a cross-sectional survey, we cannot infer the direction of results, but it is likely that respondents believed they were immune to mpox because of their recent vaccination. This is interesting given limited information about the effectiveness of the vaccine at that time.

Strengths of the study include the collection of data from four large samples, including the general population and the population most affected by the recent mpox outbreak (GBMSM). We would also caution readers about several caveats for this study, including that participant socio-demographic characteristics differed by the recruitment method. Participants recruited through Grindr and Meta were more likely to be highly educated, higher socio-economic grade, and have less financial hardship. High rates of smallpox vaccination in these samples suggest that these groups are likely to be interested in and have personal experience relating to the outbreak and may have been more likely to engage with protective behaviours. We do not know if survey respondents are representative in terms of their beliefs, knowledge and intended behaviours with reference to the general population sample, GBMSM generally, and GBMSM who use Grindr and Meta. While overall rates of beliefs, knowledge and intended behaviours may be affected by sampling, associations within the data are likely to hold true.(60)

Other limitations relate to the survey and statistical measures used. The use of a cross-sectional survey means that answers may have been influenced by social desirability and recall bias, although the anonymity of a written survey may have mitigated these effects to some degree. Rates of intended behaviour are often higher than enacted behaviours.(11, 43) Therefore, rates of intended behaviours should be taken with caution. Our motivational messages were very brief and only repeated once, before measuring intended behaviours and behaviour-specific attitudes. A fully developed public communications campaign where people are exposed to co-produced and repeated messages may have more influence. There were high rates of actual and intended vaccination in the Grindr sample (94%). This may have affected our ability to detect associations. Variables were entered into regression models in blocks, with results reported for the final block entered (essentially all variables entered together). Therefore, we investigated the independent effect of a variable, accounting for all other variables included.

This study investigated mpox beliefs, knowledge and intended behaviours in a general population sample and in GBMSM (those most affected by the current outbreak). Intended uptake of protective behaviours differed by behaviour. GBMSM generally had higher intention to engage with protective behaviours, apart from self-isolation. This may have been a function of sampling. Higher knowledge about mpox symptoms and transmission in GBMSM samples suggests that the public health messaging carried out by multiple stakeholders including the UK Health Security Agency, charities and via grass-roots community efforts has been successful. There was no impact of additional motivational messaging on intended uptake of protective behaviours. Associations between increased financial hardship and lower intention to enact protective behaviours suggest that providing financial support to those affected in future outbreaks may increase uptake.

## Data Availability

Anonymised data will be made available online when the manuscript is published in a peer-reviewed journal.

## DATA SHARING STATEMENT

Anonymised data are available online.(61)

## AUTHOR CONTRIBUTION STATEMENT

All authors conceptualised the study and contributed to survey materials. LS completed analyses with guidance from HWWP, JB and GJR. LS wrote the first draft of the manuscript. All authors contributed to, and approved, the final manuscript. LS is guarantor. The corresponding author attests that all listed authors meet authorship criteria and that no others meeting the criteria have been omitted.

## DISSEMINATION DECLARATION

Dissemination of survey results to participants is not possible due to the anonymous nature of data collection.

## ACKNOWLEDGEMENTS

We would like to thank the Terrence Higgins Trust for their support with participant recruitment, and Savanta for hosting the survey. We would also like to thank study participants, for taking their time to complete the survey.

## Supplementary materials 1. Full survey materials and top-line results for general population and GBMSM samples separately

For categorical data, n (%) are shown. Percentages are column percentages. Where totals do not add up to 100%, it is due to rounding errors.

For continuous data, n (where different to total sample), mean (M), standard deviation (SD), and range are shown.

Total *n*s are: General population, n=3050; Savanta GBMSM, n=247; Grindr, n=831; Meta [Facebook and Instagram], n=1036.

### Screening questions

ASK ALL

Postcode What is your full postcode?

*We will only use your postcode to allocate you to groups based on your location (e.g. region). Your full postcode will not be passed on to the research team*.

Type your answer below

OPEN END

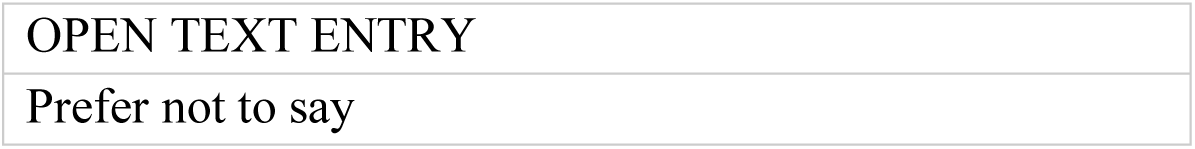

*New screen*

ASK ALL

Gender What is your gender?

SINGLE CODE

|  | General population | Savanta GBMSM | Grindr | Meta |
| --- | --- | --- | --- | --- |
| Male (including trans man) | 1282 (42.0) | 247 (100.0) | 827 (99.5) | 1030 (99.4) |
| Female (including trans woman) | 1729 (56.7) | 0 (0.0) | 0 (0.0) | 0 (0.0) |
| Non-binary | 17 (0.6) | 0 (0.0) | 0 (0.0) | 0 (0.0) |
| Other, please state | 15 (0.5) | 0 (0.0) | 4 (0.5) | 6 (0.6) |
| Prefer not to say | 7 (0.2) | 0 (0.0) | 0 (0.0) | 0 (0.0) |

\* For main analyses, where possible, text answers from “other, please state” were used to assign participants to a group. Therefore, numbers presented in the main manuscript may differ slightly.

*New screen*

ASK ALL

SexBirth Is your gender the same as your gender assigned at birth?

SINGLE CODE

|  | General population | Savanta GBMSM | Grindr | Meta |
| --- | --- | --- | --- | --- |
| Yes | 3022 (99.1) | 238 (96.4) | 828 (99.6) | 1013 (97.8) |
| No | 22 (0.7) | 9 (3.6) | 2 (0.2) | 18 (1.7) |
| Prefer not to say | 6 (0.2) | 0 (0.0) | 1 (0.1) | 5 (0.5) |

*New screen*

ASK ALL

Age How old are you (in years)?

Type your answer below

NUMERIC - OPEN END

|  | General population | Savanta GBMSM | Grindr | Meta |
| --- | --- | --- | --- | --- |
| OPEN TEXT ENTRY – number [cap at 110] | M=48.6, SD=17.4, range 18 to 98. | M=47.1, SD=16.5, range 18 to 77. | M=44.2, SD=12.5, range 18 to 82. | M=47.6, SD=11.9, range 18 to 79. |

*SCREEN OUT if under 18 years*.

*New screen*

ASK ALL

Sexuality Which of the following best describes your sexual orientation?

SINGLE CODE, RANDOMISE

|  | General population | Savanta GBMSM | Grindr | Meta |
| --- | --- | --- | --- | --- |
| Straight or heterosexual | 2795 (91.6) | 0 (0.0) | 0 (0.0) | 0 (0.0) |
| Gay or lesbian | 78 (2.6) | 230 (93.1) | 706 (85.0) | 967 (93.3) |
| Bisexual | 135 (4.4) | 16 (6.5) | 118 (14.2) | 55 (5.3) |
| Other sexual orientation, please state [ANCHOR] | 18 (0.6) | 1 (0.4) | 7 (0.8) | 14 (1.4) |
| Prefer not to say [ANCHOR] | 24 (0.8) | 0 (0.0) | 0 (0.0) | 0 (0.0) |

*\* For main analyses, where possible, text answers from “other, please state” were used to assign participants to a group. Therefore, numbers presented in the main manuscript may differ slightly.*

*New screen*

ASK ALL

SEG Which of the following best describes the profession of the chief income earner in your household?

SINGLE CODE

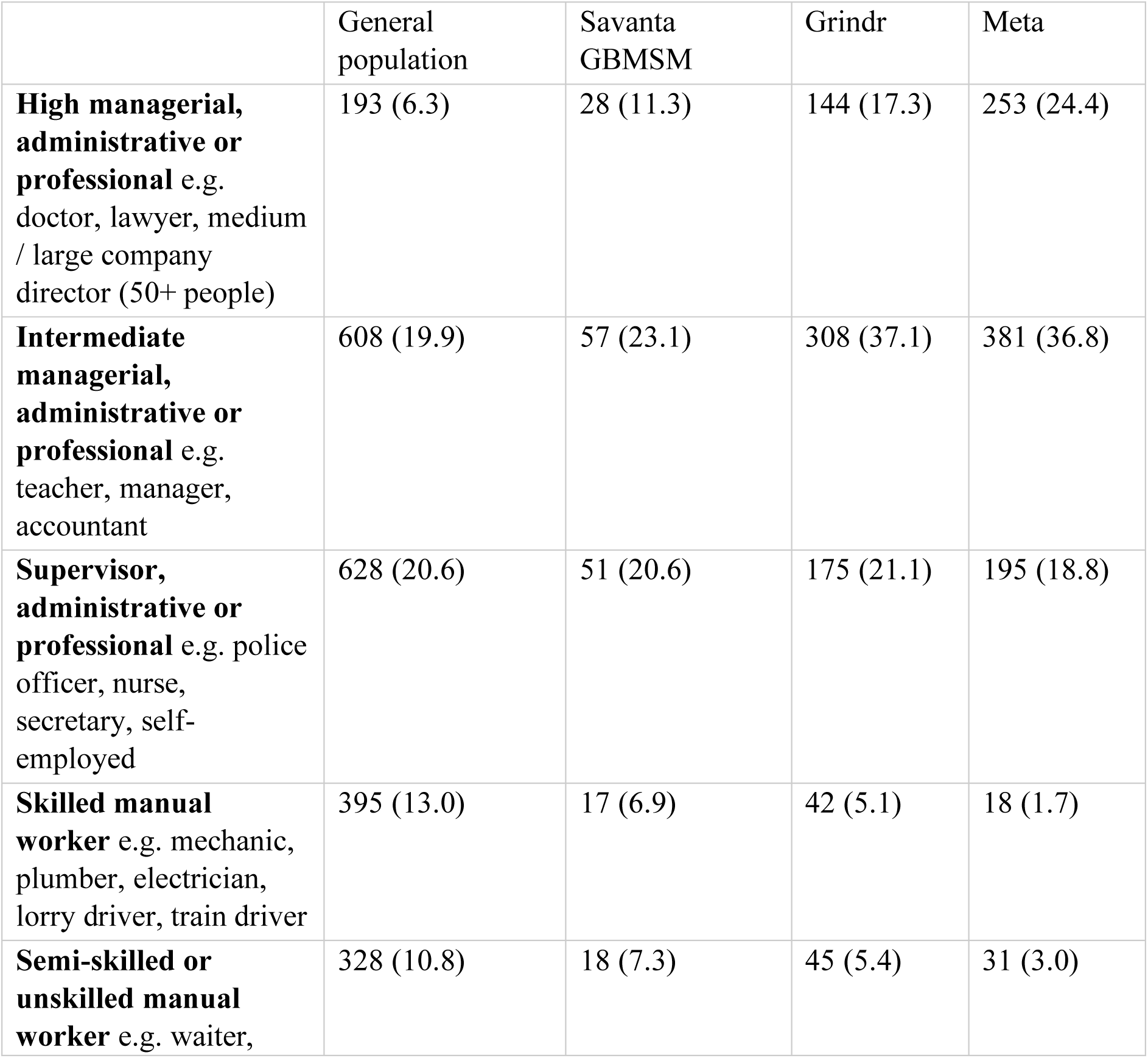
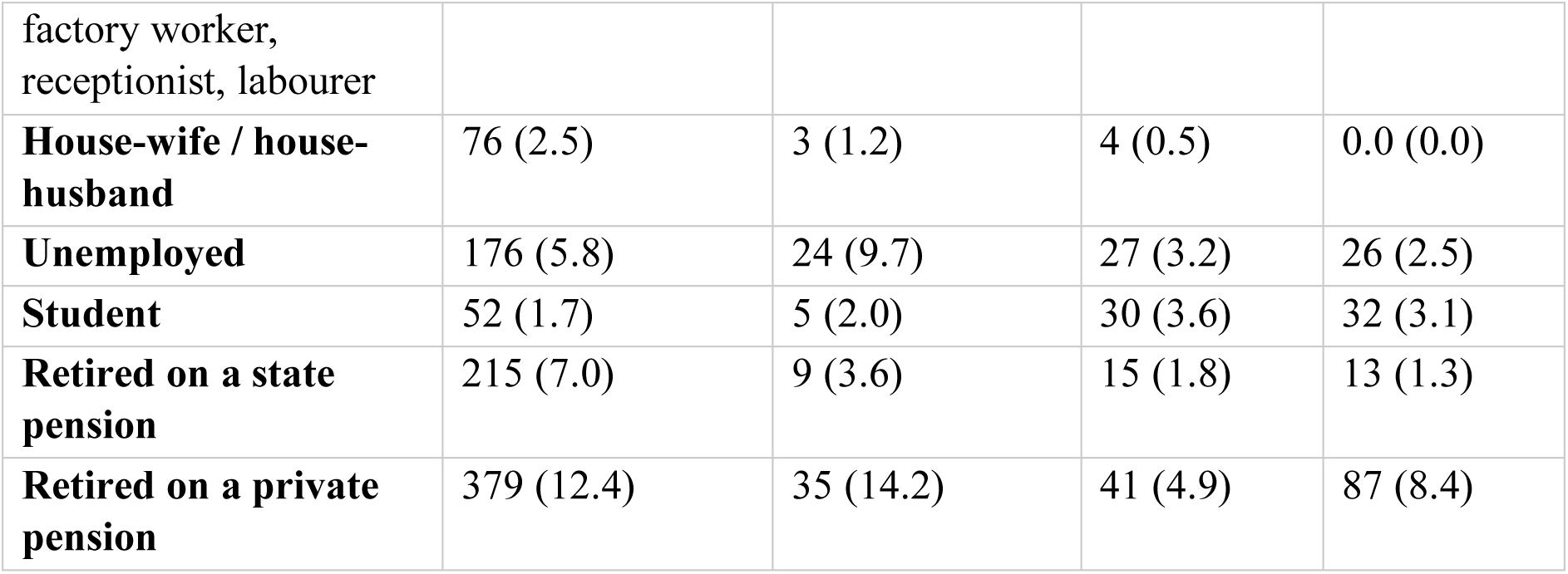

*New screen*

ASK ALL

QC This is a quality control question.

What colour is grass usually?

SINGLE CODE, RANDOMISE ORDER

|  | General population | Savanta GBMSM | Grindr | Meta |
| --- | --- | --- | --- | --- |
| Purple | 0 (0.0) | 0 (0.0) | 0 (0.0) | 0 (0.0) |
| Pink | 0 (0.0) | 0 (0.0) | 0 (0.0) | 0 (0.0) |
| Orange | 0 (0.0) | 0 (0.0) | 0 (0.0) | 0 (0.0) |
| Green | 3050 (100.0) | 247 (100.0) | 831 (100.0) | 1036 (100.0) |
| White | 0 (0.0) | 0 (0.0) | 0 (0.0) | 0 (0.0) |
| Black | 0 (0.0) | 0 (0.0) | 0 (0.0) | 0 (0.0) |

*SCREEN OUT if do not select “green”*.

### Beliefs and attitudes

*New screen*

There is an infection called “monkeypox” that has affected some people in the UK. We are interested to know your personal opinion about monkeypox, based on what you currently know.

For each question, please select the answer that reflects your opinion about monkeypox. Do not worry if you do not know what the best answer might be or if you are not at all familiar with monkeypox. We only ask that you try to give your answer based on what you think you know or what you would honestly decide to do in the situations described. If you are really not sure, please answer “don’t know”.

*New screen*

Q1. Before today, how much have you seen or heard about monkeypox?

|  | General population | Savanta GBMSM | Grindr | Meta |
| --- | --- | --- | --- | --- |
| I have not seen or heard anything | 117 (3.8) | 6 (2.4) | 7 (0.8) | 2 (0.2) |
| I have seen or heard a little | 2164 (71.0) | 136 (55.1) | 385 (46.3) | 287 (27.7) |
| I have seen or heard a lot | 759 (24.9) | 103 (41.7) | 439 (52.8) | 747 (72.1) |
| Don't know | 10 (0.3) | 2 (0.8) | 0 (0.0) | 0 (0.0) |

*New screen*

ASK ALL

Q2. Overall, how worried are you about monkeypox?

SINGLE CODE

|  | General population | Savanta GBMSM | Grindr | Meta |
| --- | --- | --- | --- | --- |
| Not at all worried | 635 (20.8) | 40 (16.2) | 77 (9.3) | 47 (4.5) |
| Not very worried | 1763 (57.8) | 125 (50.6) | 469 (56.4) | 500 (48.3) |
| Very worried | 452 (14.8) | 61 (24.7) | 234 (28.2) | 410 (39.6) |
| Extremely worried | 129 (4.2) | 13 (5.3) | 38 (4.6) | 72 (6.9) |
| Don't know | 71 (2.3) | 8 (3.2) | 13 (1.6) | 7 (0.7) |

*New screen*

ASK ALL

Q3. How much risk do you think monkeypox currently poses to:

Please select one option for each answer

RANDOMISE order of presentation of statements

People in the UK?

|  | General population | Savanta GBMSM | Grindr | Meta |
| --- | --- | --- | --- | --- |
| No risk at all | 106 (3.5) | 9 (3.6) | 11 (1.3) | 3 (0.3) |
| Low risk | 1537 (50.4) | 102 (41.3) | 357 (43.0) | 427 (41.2) |
| Medium risk | 1058 (34.7) | 97 (39.3) | 337 (40.6) | 426 (41.1) |
| High risk | 186 (6.1) | 28 (11.3) | 92 (11.1) | 153 (14.8) |
| Very high risk | 75 (2.5) | 6 (2.4) | 17 (2.0) | 22 (2.1) |
| Don't know | 88 (2.9) | 5 (2.0) | 17 (2.0) | 5 (0.5) |

You personally?

|  | General population | Savanta GBMSM | Grindr | Meta |
| --- | --- | --- | --- | --- |
| No risk at all | 700 (23.0) | 48 (19.4) | 42 (5.1) | 44 (4.2) |
| Low risk | 1619 (53.1) | 96 (38.9) | 291 (35.0) | 266 (25.7) |
| Medium risk | 441 (14.5) | 71 (28.7) | 303 (36.5) | 332 (32.0) |
| High risk | 136 (4.5) | 15 (6.1) | 148 (17.8) | 299 (28.9) |
| Very high risk | 58 (1.9) | 13 (5.3) | 37 (4.5) | 93 (9.0) |
| Don't know | 96 (3.1) | 4 (1.6) | 10 (1.2) | 2 (0.2) |

*New screen*

ASK ALL

Q4. How much, if at all, have you seen or heard about monkeypox? From:

The news (TV, online news websites or apps, printed newspapers, or radio)

|  | General population | Savanta GBMSM | Grindr | Meta |
| --- | --- | --- | --- | --- |
| Not at all | 210 (6.9) | 14 (5.7) | 61 (7.3) | 54 (5.2) |
| A little | 2125 (69.7) | 149 (60.3) | 533 (64.1) | 652 (62.9) |
| A lot | 693 (22.7) | 82 (33.2) | 235 (28.3) | 330 (31.9) |
| Don't know | 22 (0.7) | 2 (0.8) | 2 (0.2) | 0 (0.0) |

Official websites or helplines (e.g. NHS, GOV.UK), or an NHS GP practice, clinic or hospital

|  | General population | Savanta GBMSM | Grindr | Meta |
| --- | --- | --- | --- | --- |
| Not at all | 1547 (50.7) | 98 (39.7) | 198 (23.8) | 188 (18.1) |
| A little | 1113 (36.5) | 97 (39.3) | 450 (54.2) | 601 (58.0) |
| A lot | 316 (10.4) | 48 (19.4) | 169 (20.3) | 241 (23.3) |
| Don't know | 74 (2.4) | 4 (1.6) | 14 (1.7) | 6 (0.6) |

Speaking to friends, family, colleagues or other people you know (in person, by phone, text, WhatsApp, email, or in other ways)

|  | General population | Savanta GBMSM | Grindr | Meta |
| --- | --- | --- | --- | --- |
| Not at all | 1469 (48.2) | 111 (44.9) | 263 (31.6) | 218 (21.0) |
| A little | 1235 (40.5) | 87 (35.2) | 388 (46.7) | 466 (45.0) |
| A lot | 300 (9.8) | 44 (17.8) | 175 (21.1) | 350 (33.8) |
| Don't know | 46 (1.5) | 5 (2.0) | 5 (0.6) | 2 (0.2) |

Workers in community groups, community or faith leaders, charities, or volunteers who help improve the health and wellbeing of others

|  | General population | Savanta GBMSM | Grindr | Meta |
| --- | --- | --- | --- | --- |
| Not at all | 2111 (69.2) | 165 (66.8) | 553 (66.5) | 633 (61.1) |
| A little | 633 (20.8) | 53 (21.5) | 162 (19.5) | 253 (24.4) |
| A lot | 220 (7.2) | 25 (10.1) | 58 (7.0) | 113 (10.9) |
| Don't know | 86 (2.8) | 4 (1.6) | 58 (7.0) | 37 (3.6) |

Other places, including blogs, social media sites (e.g. Facebook, Twitter, Instagram), online communities or other websites

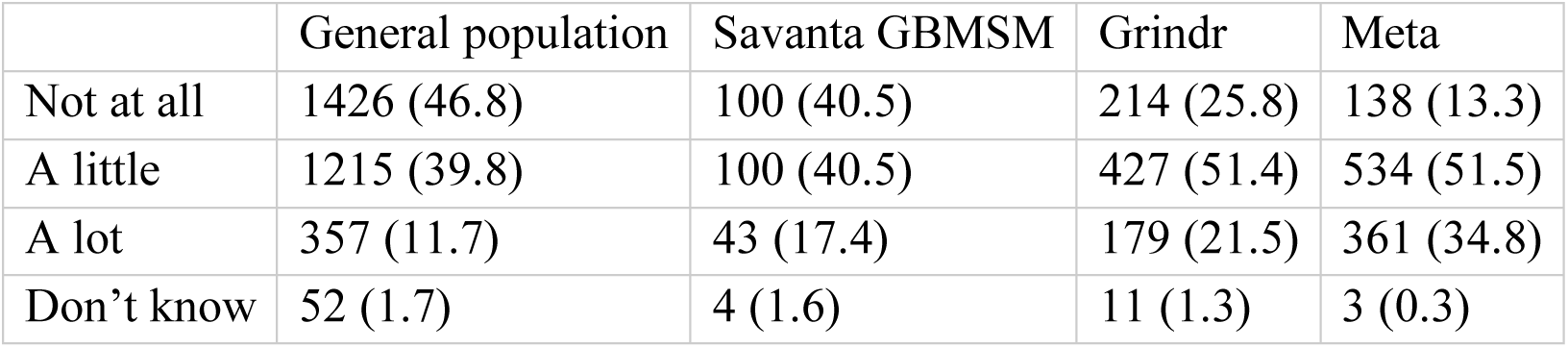

*New screen*

Q4_other. Please state any other places that you have seen or heard about monkeypox from.

Open ended, text entry

None button

*New screen*

For each question, please select the answer that reflects your opinion about monkeypox. Do not worry if you do not know what the best answer might be or if you are not at all familiar with monkeypox. We only ask that you try to give your answer based on what you think you know. If you are really not sure, please answer “don’t know”.

*New screen*

ASK ALL

Q5. How much do you agree or disagree with the following statements:

RANDOMISE statements

I have a good idea of how people catch monkeypox

|  | General population | Savanta GBMSM | Grindr | Meta |
| --- | --- | --- | --- | --- |
| Strongly disagree | 252 (8.2) | 14 (5.6) | 27 (3.2) | 15 (1.4) |
| Disagree | 727 (23.8) | 35 (14.1) | 46 (5.5) | 31 (2.9) |
| Neither agree nor disagree | 590 (19.3) | 32 (12.9) | 82 (9.8) | 57 (5.5) |
| Agree | 1159 (38.0) | 121 (48.9) | 468 (56.3) | 559 (53.9) |
| Strongly agree | 217 (7.1) | 41 (16.5) | 196 (23.5) | 371 (35.8) |
| Don't know | 105 (3.4) | 4 (1.6) | 12 (1.4) | 3 (0.2) |

I know what the main symptoms of monkeypox are

|  | General population | Savanta GBMSM | Grindr | Meta |
| --- | --- | --- | --- | --- |
| Strongly disagree | 263 (8.6) | 16 (6.5) | 38 (4.6) | 17 (1.6) |
| Disagree | 820 (26.9) | 44 (17.8) | 100 (12.0) | 60 (5.8) |
| Neither agree nor disagree | 590 (19.3) | 41 (16.6) | 106 (12.8) | 108 (10.4) |
| Agree | 1113 (36.5) | 120 (48.6) | 493 (59.3) | 642 (62.0) |
| Strongly agree | 152 (5.0) | 23 (9.3) | 80 (9.6) | 204 (19.7) |
| Don't know | 112 (3.7) | 3 (1.2) | 14 (1.7) | 5 (0.5) |

It would be easy for me to tell if someone I meet has monkeypox

|  | General population | Savanta GBMSM | Grindr | Meta |
| --- | --- | --- | --- | --- |
| Strongly disagree | 485 (15.9) | 33 (13.4) | 149 (17.9) | 205 (19.8) |
| Disagree | 1123 (36.8) | 71 (28.7) | 369 (44.4) | 498 (48.1) |
| Neither agree nor disagree | 663 (21.7) | 64 (25.9) | 181 (21.8) | 209 (20.2) |
| Agree | 504 (16.5) | 54 (21.9) | 88 (10.6) | 92 (8.9) |
| Strongly agree | 119 (3.9) | 14 (5.7) | 15 (1.8) | 15 (1.4) |
| Don't know | 156 (5.1) | 11 (4.5) | 29 (3.5) | 17 (1.6) |

*New screen*

ASK ALL

Q6. How much do you agree or disagree with the following statements:

RANDOMISE statements

In the near future, it is likely that some of the people I come into physical contact with (touch) will have monkeypox

|  | General population | Savanta GBMSM | Grindr | Meta |
| --- | --- | --- | --- | --- |
| Strongly disagree | 275 (9.0) | 13 (5.3) | 23 (2.8) | 23 (2.2) |
| Disagree | 743 (24.4) | 56 (22.7) | 144 (17.3) | 145 (14.0) |
| Neither agree nor disagree | 912 (29.9) | 65 (26.3) | 237 (28.5) | 238 (23.0) |
| Agree | 632 (20.7) | 73 (29.6) | 297 (35.7) | 470 (45.4) |
| Strongly agree | 112 (3.7) | 12 (4.9) | 43 (5.2) | 118 (11.4) |
| Don't know | 376 (12.3) | 28 (11.3) | 87 (10.5) | 42 (4.1) |

If I come into physical contact with (touch) someone who has monkeypox, it is likely that I will catch it

|  | General population | Savanta GBMSM | Grindr | Meta |
| --- | --- | --- | --- | --- |
| Strongly disagree | 108 (3.5) | 5 (2.0) | 20 (2.4) | 9 (0.9) |
| Disagree | 467 (15.3) | 24 (9.7) | 93 (11.2) | 145 (14.0) |
| Neither agree nor disagree | 814 (26.7) | 59 (23.9) | 167 (20.1) | 207 (20.0) |
| Agree | 944 (31.0) | 99 (40.1) | 376 (45.2) | 497 (48.0) |
| Strongly agree | 207 (6.8) | 25 (10.1) | 98 (11.8) | 135 (13.0) |
| Don't know | 510 (16.7) | 35 (14.2) | 77 (9.3) | 43 (4.2) |

Monkeypox would be a serious illness for me

|  | General population | Savanta GBMSM | Grindr | Meta |
| --- | --- | --- | --- | --- |
| Strongly disagree | 224 (7.3) | 16 (6.5) | 30 (3.6) | 26 (2.5) |
| Disagree | 546 (17.9) | 35 (14.2) | 144 (17.3) | 160 (15.4) |
| Neither agree nor disagree | 744 (24.4) | 58 (23.5) | 182 (21.9) | 220 (21.2) |
| Agree | 787 (25.8) | 85 (34.4) | 290 (34.9) | 413 (39.9) |
| Strongly agree | 274 (9.0) | 28 (11.3) | 116 (14.0) | 166 (16.0) |
| Don't know | 475 (15.6) | 25 (10.1) | 69 (8.3) | 51 (4.9) |

People who catch monkeypox usually make a full recovery, even if they do not receive any treatment

|  | General population | Savanta GBMSM | Grindr | Meta |
| --- | --- | --- | --- | --- |
| Strongly disagree | 52 (1.7) | 0 (0.0) | 11 (1.3) | 6 (0.6) |
| Disagree | 264 (8.7) | 25 (10.1) | 77 (9.3) | 62 (6.0) |
| Neither agree nor disagree | 844 (27.7) | 56 (22.7) | 117 (14.1) | 144 (13.9) |
| Agree | 977 (32.0) | 96 (38.9) | 383 (46.1) | 530 (51.2) |
| Strongly agree | 228 (7.5) | 29 (11.7) | 102 (12.3) | 210 (20.3) |
| Don't know | 685 (22.5) | 41 (16.6) | 141 (17.0) | 84 (8.1) |

My personal behaviour has an impact on how monkeypox spreads

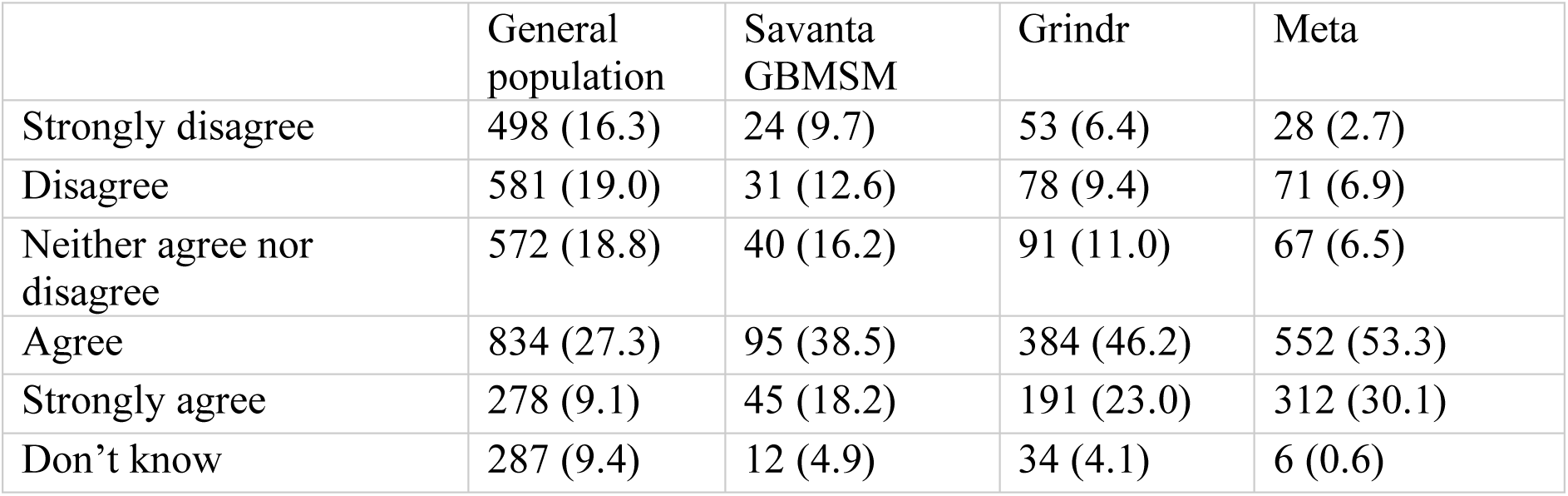

|  | General population | Savanta GBMSM | Grindr | Meta |
| --- | --- | --- | --- | --- |
| Strongly disagree | 498 (16.3) | 24 (9.7) | 53 (6.4) | 28 (2.7) |
| Disagree | 581 (19.0) | 31 (12.6) | 78 (9.4) | 71 (6.9) |
| Neither agree nor disagree | 572 (18.8) | 40 (16.2) | 91 (11.0) | 67 (6.5) |
| Agree | 834 (27.3) | 95 (38.5) | 384 (46.2) | 552 (53.3) |
| Strongly agree | 278 (9.1) | 45 (18.2) | 191 (23.0) | 312 (30.1) |
| Don't know | 287 (9.4) | 12 (4.9) | 34 (4.1) | 6 (0.6) |

My life has been negatively affected by changes made in response to the monkeypox outbreak

|  | General population | Savanta GBMSM | Grindr | Meta |
| --- | --- | --- | --- | --- |
| Strongly disagree | 1181 (38.7) | 90 (36.4) | 167 (20.1) | 160 (15.4) |
| Disagree | 981 (32.2) | 75 (30.4) | 289 (34.8) | 300 (29.0) |
| Neither agree nor disagree | 420 (13.8) | 37 (15.0) | 168 (20.2) | 217 (20.9) |
| Agree | 246 (8.1) | 24 (9.7) | 144 (17.3) | 263 (25.4) |
| Strongly agree | 115 (3.8) | 12 (4.9) | 42 (5.1) | 88 (8.5) |
| Don't know | 107 (3.5) | 9 (3.6) | 21 (2.5) | 8 (0.8) |

*New screen*

Below are some claims that some people have made about monkeypox. For each one, please say whether you strongly disagree, disagree, neither agree nor disagree, agree, or strongly agree.

ASK ALL

Q7. How much do you agree or disagree with the following claims:

SINGLE CODE

RANDOMISE statements

The risks of monkeypox are being exaggerated

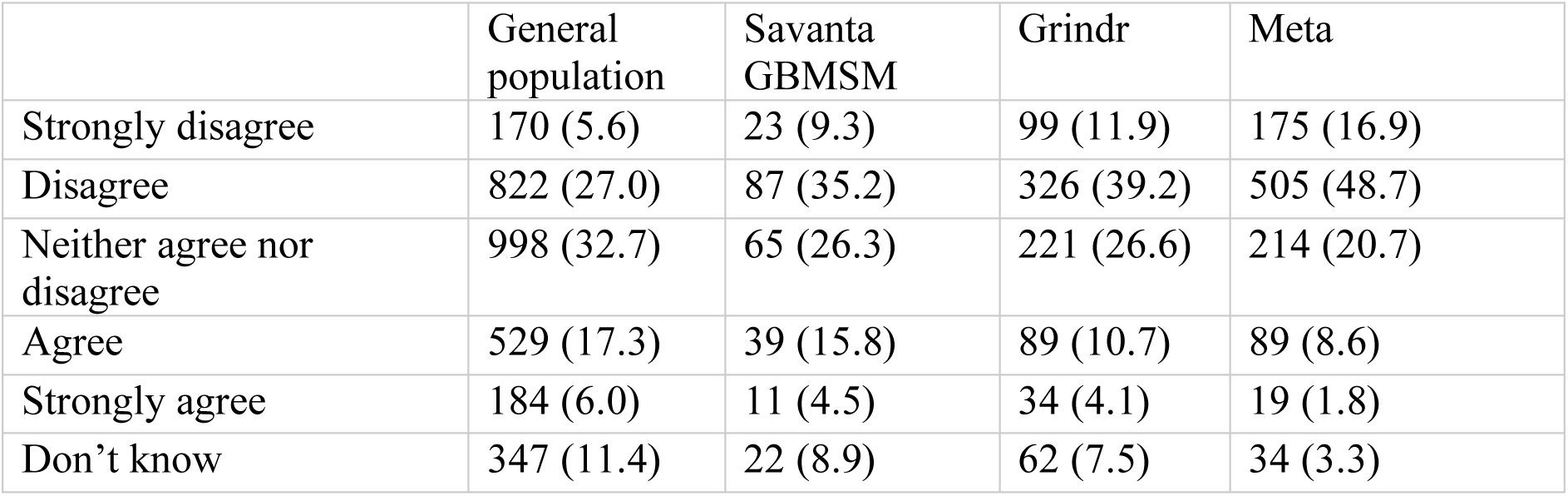

|  | General population | Savanta GBMSM | Grindr | Meta |
| --- | --- | --- | --- | --- |
| Strongly disagree | 170 (5.6) | 23 (9.3) | 99 (11.9) | 175 (16.9) |
| Disagree | 822 (27.0) | 87 (35.2) | 326 (39.2) | 505 (48.7) |
| Neither agree nor disagree | 998 (32.7) | 65 (26.3) | 221 (26.6) | 214 (20.7) |
| Agree | 529 (17.3) | 39 (15.8) | 89 (10.7) | 89 (8.6) |
| Strongly agree | 184 (6.0) | 11 (4.5) | 34 (4.1) | 19 (1.8) |
| Don't know | 347 (11.4) | 22 (8.9) | 62 (7.5) | 34 (3.3) |

Because of the monkeypox outbreak, it is best to avoid physical contact with (touch) men who are gay, bisexual or have sex with men

|  | General population | Savanta GBMSM | Grindr | Meta |
| --- | --- | --- | --- | --- |
| Strongly disagree | 422 (13.8) | 50 (20.2) | 237 (28.5) | 294 (28.4) |
| Disagree | 707 (23.2) | 64 (25.9) | 267 (32.1) | 357 (34.5) |
| Neither agree nor disagree | 667 (21.9) | 44 (17.8) | 147 (17.7) | 172 (16.6) |
| Agree | 611 (20.0) | 51 (20.6) | 132 (15.9) | 161 (15.5) |
| Strongly agree | 305 (10.0) | 22 (8.9) | 22 (2.6) | 37 (3.6) |
| Don't know | 338 (11.1) | 16 (6.5) | 26 (3.1) | 15 (1.4) |

Because of the monkeypox outbreak, it is best to avoid physical contact with (touch) people from Africa

|  | General population | Savanta GBMSM | Grindr | Meta |
| --- | --- | --- | --- | --- |
| Strongly disagree | 462 (15.1) | 52 (21.1) | 327 (39.4) | 431 (41.6) |
| Disagree | 845 (27.7) | 69 (27.9) | 273 (32.9) | 399 (38.5) |
| Neither agree nor disagree | 768 (25.2) | 57 (23.1) | 118 (14.2) | 127 (12.3) |
| Agree | 454 (14.9) | 40 (16.2) | 59 (7.1) | 43 (4.2) |
| Strongly agree | 175 (5.7) | 10 (4.0) | 14 (1.7) | 11 (1.1) |
| Don't know | 346 (11.3) | 19 (7.7) | 40 (4.8) | 25 (2.4) |

Monkeypox is only a risk to men who are gay, bisexual or have sex with men

|  | General population | Savanta GBMSM | Grindr | Meta |
| --- | --- | --- | --- | --- |
| Strongly disagree | 536 (17.6) | 77 (31.2) | 375 (45.1) | 431 (41.6) |
| Disagree | 909 (29.8) | 75 (30.4) | 302 (36.3) | 449 (43.3) |
| Neither agree nor disagree | 586 (19.2) | 35 (14.2) | 56 (6.7) | 63 (6.1) |
| Agree | 521 (17.1) | 30 (12.1) | 61 (7.3) | 65 (6.3) |
| Strongly agree | 161 (5.3) | 15 (6.1) | 14 (1.7) | 21 (2.0) |
| Don't know | 337 (11.0) | 15 (6.1) | 23 (2.8) | 7 (0.7) |

*New screen*

ASK ALL

Q8. What do you think the main symptoms of monkeypox are?

Please select up to 4

MULTICODE – MAX 4

RANDOMISE

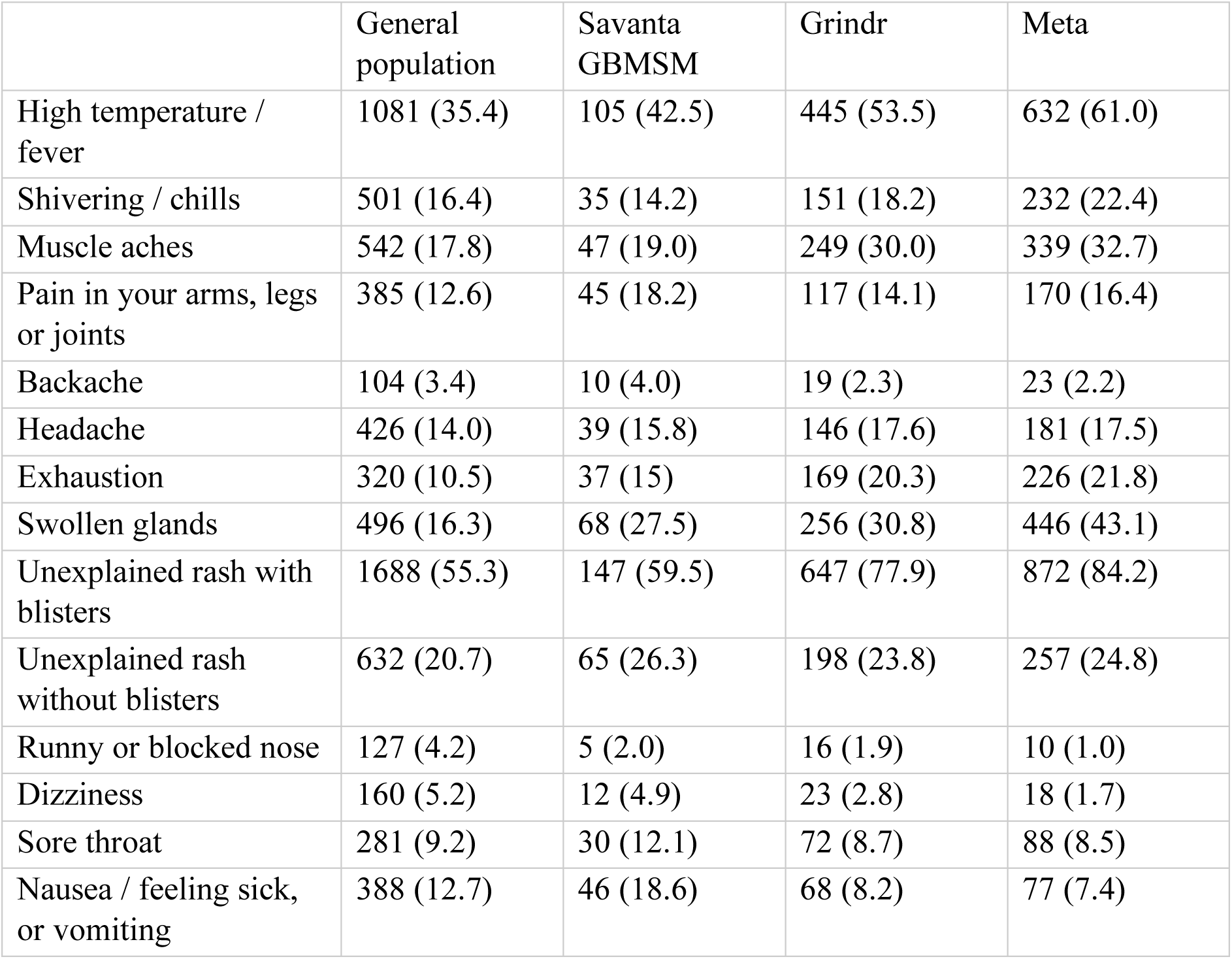
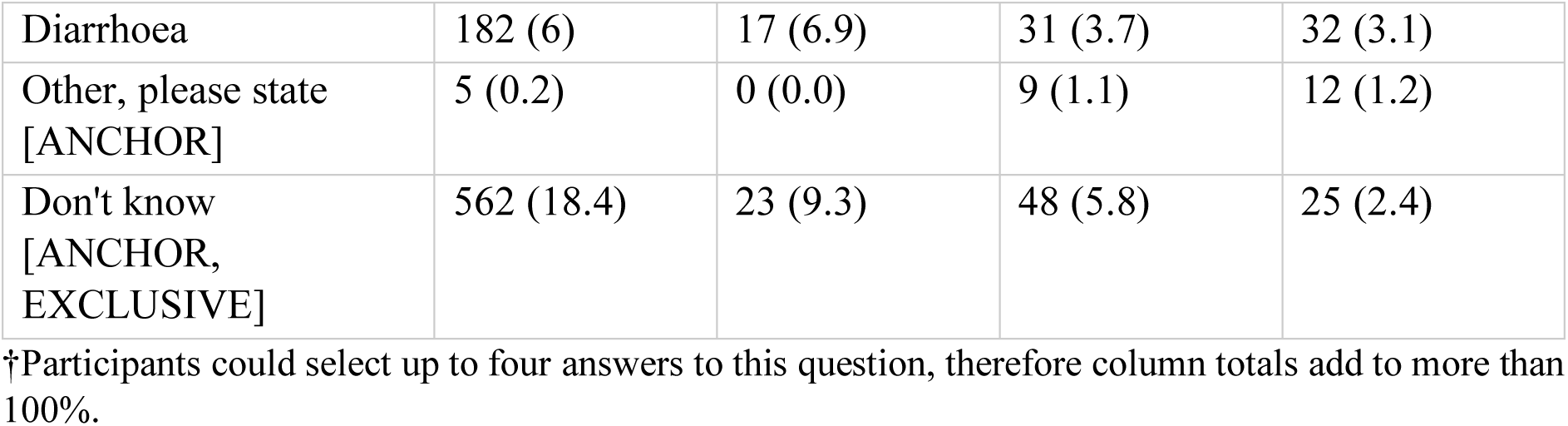

|  | General population | Savanta GBMSM | Grindr | Meta |
| --- | --- | --- | --- | --- |
| High temperature / fever | 1081 (35.4) | 105 (42.5) | 445 (53.5) | 632 (61.0) |
| Shivering / chills | 501 (16.4) | 35 (14.2) | 151 (18.2) | 232 (22.4) |
| Muscle aches | 542 (17.8) | 47 (19.0) | 249 (30.0) | 339 (32.7) |
| Pain in your arms, legs or joints | 385 (12.6) | 45 (18.2) | 117 (14.1) | 170 (16.4) |
| Backache | 104 (3.4) | 10 (4.0) | 19 (2.3) | 23 (2.2) |
| Headache | 426 (14.0) | 39 (15.8) | 146 (17.6) | 181 (17.5) |
| Exhaustion | 320 (10.5) | 37 (15) | 169 (20.3) | 226 (21.8) |
| Swollen glands | 496 (16.3) | 68 (27.5) | 256 (30.8) | 446 (43.1) |
| Unexplained rash with blisters | 1688 (55.3) | 147 (59.5) | 647 (77.9) | 872 (84.2) |
| Unexplained rash without blisters | 632 (20.7) | 65 (26.3) | 198 (23.8) | 257 (24.8) |
| Runny or blocked nose | 127 (4.2) | 5 (2.0) | 16 (1.9) | 10 (1.0) |
| Dizziness | 160 (5.2) | 12 (4.9) | 23 (2.8) | 18 (1.7) |
| Sore throat | 281 (9.2) | 30 (12.1) | 72 (8.7) | 88 (8.5) |
| Nausea / feeling sick, or vomiting | 388 (12.7) | 46 (18.6) | 68 (8.2) | 77 (7.4) |
| Diarrhoea | 182 (6) | 17 (6.9) | 31 (3.7) | 32 (3.1) |
| Other, please state<br>[ANCHOR] | 5 (0.2) | 0 (0.0) | 9 (1.1) | 12 (1.2) |
| Don't know<br>[ANCHOR,<br>EXCLUSIVE] | 562 (18.4) | 23 (9.3) | 48 (5.8) | 25 (2.4) |

†Participants could select up to four answers to this question, therefore column totals add to more than 100%.

*New screen*

ASK ALL

Q9. You can catch monkeypox if:

Please select one option for each answer

RANDOMISE statements

You touch the rash of a person who has monkeypox

|  | General population | Savanta GBMSM | Grindr | Meta |
| --- | --- | --- | --- | --- |
| Strongly disagree | 68 (2.2) | 3 (1.2) | 11 (1.3) | 6 (0.6) |
| Disagree | 176 (5.8) | 10 (4.0) | 23 (2.8) | 15 (1.4) |
| Neither agree nor disagree | 516 (16.9) | 28 (11.3) | 48 (5.8) | 38 (3.7) |
| Agree | 1326 (43.5) | 112 (45.3) | 394 (47.4) | 424 (40.9) |
| Strongly agree | 394 (12.9) | 65 (26.3) | 293 (35.3) | 519 (50.1) |
| Don't know | 570 (18.7) | 29 (11.7) | 62 (7.5) | 34 (3.3) |

You are coughed or sneezed on by a person who has monkeypox

|  | General population | Savanta GBMSM | Grindr | Meta |
| --- | --- | --- | --- | --- |
| Strongly disagree | 82 (2.7) | 17 (6.9) | 50 (6.0) | 82 (7.9) |
| Disagree | 356 (11.7) | 39 (15.8) | 191 (23.0) | 253 (24.4) |
| Neither agree nor disagree | 634 (20.8) | 41 (16.6) | 127 (15.3) | 160 (15.4) |
| Agree | 1024 (33.6) | 87 (35.2) | 251 (30.2) | 344 (33.2) |
| Strongly agree | 273 (9.0) | 22 (8.9) | 74 (8.9) | 82 (7.9) |
| Don't know | 681 (22.3) | 41 (16.6) | 138 (16.6) | 115 (11.1) |

You touch a person who has monkeypox, even if they do not have a rash

|  | General population | Savanta GBMSM | Grindr | Meta |
| --- | --- | --- | --- | --- |
| Strongly disagree | 91 (3.0) | 4 (1.6) | 26 (3.1) | 20 (1.9) |
| Disagree | 419 (13.7) | 28 (11.3) | 139 (16.7) | 189 (18.2) |
| Neither agree nor disagree | 634 (20.8) | 61 (24.7) | 125 (15.0) | 198 (19.1) |
| Agree | 932 (30.6) | 91 (36.8) | 337 (40.6) | 410 (39.6) |
| Strongly agree | 237 (7.8) | 24 (9.7) | 73 (8.8) | 95 (9.2) |
| Don't know | 737 (24.2) | 39 (15.8) | 131 (15.8) | 124 (12) |

You have sex with someone who has monkeypox

|  | General population | Savanta GBMSM | Grindr | Meta |
| --- | --- | --- | --- | --- |
| Strongly disagree | 72 (2.4) | 5 (2.0) | 12 (1.4) | 10 (1.0) |
| Disagree | 99 (3.2) | 4 (1.6) | 5 (0.6) | 7 (0.7) |
| Neither agree nor disagree | 359 (11.8) | 17 (6.9) | 24 (2.9) | 16 (1.5) |
| Agree | 1293 (42.4) | 120 (48.6) | 387 (46.6) | 441 (42.6) |
| Strongly agree | 802 (26.3) | 89 (36.0) | 377 (45.4) | 551 (53.2) |
| Don't know | 425 (13.9) | 12 (4.9) | 26 (3.1) | 11 (1.1) |

You come within 1 metre of someone who has monkeypox

|  | General population | Savanta GBMSM | Grindr | Meta |
| --- | --- | --- | --- | --- |
| Strongly disagree | 357 (11.7) | 59 (23.9) | 269 (32.4) | 407 (39.3) |
| Disagree | 972 (31.9) | 93 (37.7) | 379 (45.6) | 438 (42.3) |
| Neither agree nor disagree | 598 (19.6) | 29 (11.7) | 72 (8.7) | 84 (8.1) |
| Agree | 358 (11.7) | 34 (13.8) | 39 (4.7) | 50 (4.8) |
| Strongly agree | 131 (4.3) | 4 (1.6) | 10 (1.2) | 11 (1.1) |
| Don't know | 634 (20.8) | 28 (11.3) | 62 (7.5) | 46 (4.4) |

You touch something (e.g. towels, bedding or clothing) that has been touched by a person who has monkeypox

|  | General population | Savanta GBMSM | Grindr | Meta |
| --- | --- | --- | --- | --- |
| Strongly disagree | 87 (2.9) | 7 (2.8) | 21 (2.5) | 18 (1.7) |
| Disagree | 414 (13.6) | 30 (12.1) | 85 (10.2) | 72 (6.9) |
| Neither agree nor disagree | 653 (21.4) | 46 (18.6) | 100 (12.0) | 89 (8.6) |
| Agree | 961 (31.5) | 96 (38.9) | 379 (45.6) | 585 (56.5) |
| Strongly agree | 248 (8.1) | 35 (14.2) | 142 (17.1) | 204 (19.7) |
| Don't know | 687 (22.5) | 33 (13.4) | 104 (12.5) | 68 (6.6) |

You touch a pet animal that has monkeypox

|  | General population | Savanta GBMSM | Grindr | Meta |
| --- | --- | --- | --- | --- |
| Strongly disagree | 130 (4.3) | 14 (5.7) | 61 (7.3) | 67 (6.5) |
| Disagree | 469 (15.4) | 43 (17.4) | 140 (16.8) | 151 (14.6) |
| Neither agree nor disagree | 693 (22.7) | 57 (23.1) | 159 (19.1) | 208 (20.1) |
| Agree | 740 (24.3) | 62 (25.1) | 219 (26.4) | 341 (32.9) |
| Strongly agree | 193 (6.3) | 24 (9.7) | 46 (5.5) | 82 (7.9) |
| Don't know | 825 (27.0) | 47 (19) | 206 (24.8) | 187 (18.1) |

### Intended behaviour

*New screen*

[For Savanta gen pop sample, randomise people to four groups. Ppts should see EITHER group A OR group B OR group C OR group D.

For Savanta GBMSM, Grindr, Meta samples, randomise people to two groups. Ppts should see EITHER group E OR group F.]

[Savanta gen pop] GROUP A (risk + perceived necessity/efficacy): Monkeypox causes mild illness for most people, but it can be very severe and can even kill some people. Pets may also be able to catch monkeypox. People who have recently been in contact with someone who might have monkeypox and get an unexplained rash should get tested quickly. If they test positive, they should avoid close contact with others to stop monkeypox from spreading further.

[Savanta gen pop] GROUP B (risk + perceived benefits): Monkeypox causes mild illness for most people, but it can be very severe and can even kill some people. Pets may also be able to catch monkeypox. People who test positive for monkeypox can take action to protect their friends, family, and pets from infection.

[Savanta gen pop] GROUP C (risk + perceived costs): Monkeypox causes mild illness for most people, but it can be very severe and can even kill some people. Pets may also be able to catch monkeypox. Imagine that people who have to self-isolate because they test positive for monkeypox will be provided with the practical and financial support that they need to do that.

[Savanta gen pop] GROUP D (control message): Monkeypox is a rare infection that is mainly found in parts of west or central Africa. Recently, there has been an outbreak of monkeypox in other countries. Since early May, more than 40,000 monkeypox cases have been reported in over 80 countries where the virus is not normally seen, including the UK.

[Savanta GBMSM / Grindr / Meta] GROUP E (all messages): Monkeypox causes mild illness for most people, but it can be very severe and can even kill some people. Pets may also be able to catch monkeypox.

People who have recently been in contact with someone who might have monkeypox and get an unexplained rash should get tested quickly. If they test positive, they should avoid close contact with others to stop monkeypox from spreading further. People who test positive for monkeypox can take action to protect their friends, family, and pets from infection. Imagine that people who have to self-isolate because they test positive for monkeypox will be provided with the practical and financial support that they need to do that.

[Savanta GBMSM / Grindr / Meta] GROUP F (control message): Monkeypox is a rare infection that is mainly found in parts of west or central Africa. Recently, there has been an outbreak of monkeypox in other countries.

There is a new outbreak of monkeypox in the UK. Since early May, more than 40,000 monkeypox cases have been reported in over 80 countries where the virus is not normally seen, including the UK. The current outbreak is the first time that the virus has been passed from person to person in the UK, and so the UK Health Security Agency (UKHSA) is working with partners to put in place guidance and measures that will prevent the spread of infection.

*New screen*

*Self-isolation*

People who have monkeypox are being asked to self-isolate for 21 days (3 weeks). They are also being asked to tell public health officials about other people they have been in contact with.

Self-isolation means staying at home and not going to work, school or public areas. It means avoiding close contact with people you live with, spending time in separate rooms as much as possible, and not having visitors to your home.

[Randomise order of Q10 and Q11]

*New screen*

ASK ALL

Q10. Imagine you are contacted by public health officials after testing and told that **you have monkeypox**. They tell you that you need to self-isolate for 21 days.

Realistically, would you self-isolate for 21 days?

SINGLE CODE

Answer Options

|  | General population | Savanta GBMSM | Grindr | Meta |
| --- | --- | --- | --- | --- |
| Definitely would not | 106 (3.5) | 10 (4.0) | 56 (6.7) | 41 (4.0) |
| Probably would not | 283 (9.3) | 24 (9.7) | 116 (14.0) | 144 (13.9) |
| Not sure | 367 (12.0) | 19 (7.7) | 74 (8.9) | 90 (8.7) |
| Probably would | 799 (26.2) | 76 (30.8) | 260 (31.3) | 327 (31.6) |
| Definitely would | 1495 (49.0) | 118 (47.8) | 325 (39.1) | 434 (41.9) |

*New screen*

ASK ALL

Q11. Imagine you are contacted by public health officials and told that **you have come into high-risk contact with someone who has monkeypox**. They tell you that you need to self-isolate for 21 days.

Realistically, would you self-isolate for 21 days?

SINGLE CODE

Answer Options

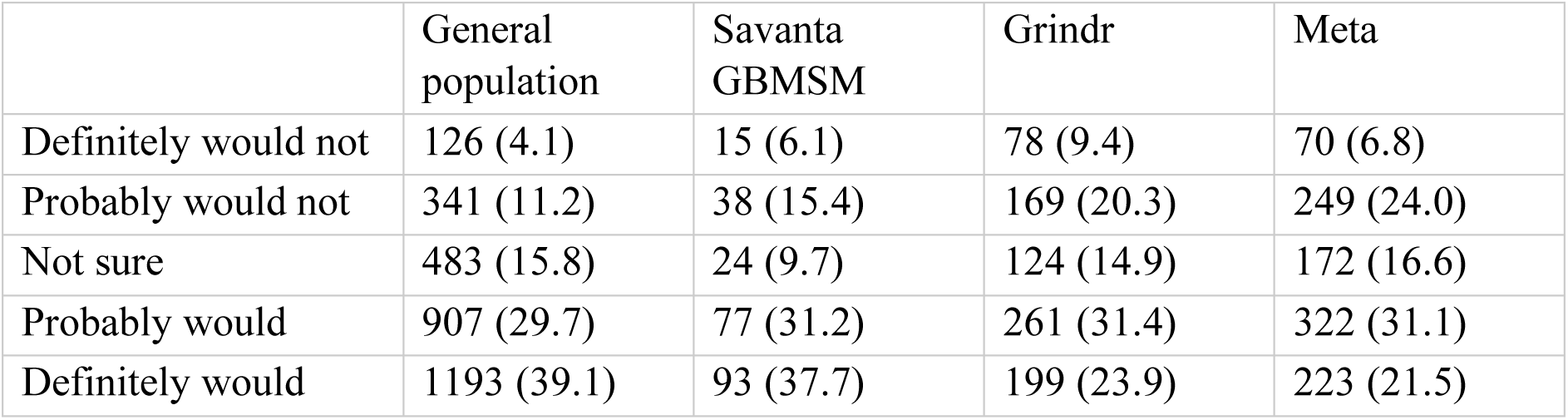

*New screen*

ASK ALL

Q12. What would stop you from being able to self-isolate for 21 days?

Please select all that apply

MULTI CODE, RANDOMISE ORDER

Answer Options

|  | General population | Savanta GBMSM | Grindr | Meta |
| --- | --- | --- | --- | --- |
| I would need to go out to get food, medicines, or other essentials | 992 (32.5) | 119 (48.2) | 477 (57.4) | 604 (58.3) |
| I would need to go out to work | 542 (17.8) | 52 (21.1) | 309 (37.2) | 323 (31.2) |
| I would need to go out to provide help or care to friends or relatives | 238 (7.8) | 22 (8.9) | 53 (6.4) | 72 (6.9) |
| I would need to go out as part of looking after my children (e.g. to take them to school, or a playground) | 396 (13.0) | 9 (3.6) | 21 (2.5) | 15 (1.4) |
| I would need to go out for a walk or some other exercise | 765 (25.1) | 72 (29.1) | 388 (46.7) | 550 (53.1) |
| My home is too small | 192 (6.3) | 17 (6.9) | 103 (12.4) | 109 (10.5) |
| I would be too bored or lonely | 344 (11.3) | 39 (15.8) | 170 (20.5) | 242 (23.4) |
| I don't think it's necessary for me to self-isolate | 207 (6.8) | 12 (4.9) | 54 (6.5) | 85 (8.2) |
| Other, please state | 40 (1.3) | 3 (1.2) | 40 (4.8) | 52 (5.0) |
| Nothing, I would stay at home for 21 days [EXCLUSIVE] | 1166 (38.2) | 76 (30.8) | 153 (18.4) | 185 (17.9) |

†Participants could select more than one item for this question, therefore column totals add to more than 100%.

*New screen*

Now we would like you to think about a different scenario.

*Help seeking*

*New screen*

Imagine that tomorrow morning **you develop an unexplained rash with blisters**. You also learn that **you have come into contact with someone who might have monkeypox**.

ASK ALL

Q13 If **you** got these symptoms, which of these things would you do?

RANDOMISE order

Wait a day or two to see if they get better or clear up on their own

|  | General population | Savanta GBMSM | Grindr | Meta |
| --- | --- | --- | --- | --- |
| Definitely would not | 344 (11.3) | 27 (10.9) | 130 (15.6) | 195 (18.8) |
| Probably would not | 546 (17.9) | 47 (19.0) | 175 (21.1) | 248 (23.9) |
| Not sure | 809 (26.5) | 58 (23.5) | 145 (17.4) | 217 (20.9) |
| Probably would | 1080 (35.4) | 92 (37.2) | 323 (38.9) | 326 (31.5) |
| Definitely would | 271 (8.9) | 23 (9.3) | 58 (7.0) | 50 (4.8) |

Try to book an appointment with a GP

|  | General population | Savanta GBMSM | Grindr | Meta |
| --- | --- | --- | --- | --- |
| Definitely would not | 307 (10.1) | 28 (11.3) | 154 (18.5) | 183 (17.7) |
| Probably would not | 409 (13.4) | 48 (19.4) | 167 (20.1) | 295 (28.5) |
| Not sure | 586 (19.2) | 40 (16.2) | 125 (15.0) | 148 (14.3) |
| Probably would | 1032 (33.8) | 86 (34.8) | 221 (26.6) | 222 (21.4) |
| Definitely would | 716 (23.5) | 45 (18.2) | 164 (19.7) | 188 (18.1) |

Visit a Pharmacist/Chemist for advice

|  | General population | Savanta GBMSM | Grindr | Meta |
| --- | --- | --- | --- | --- |
| Definitely would not | 634 (20.8) | 49 (19.8) | 284 (34.2) | 393 (37.9) |
| Probably would not | 749 (24.6) | 69 (27.9) | 329 (39.6) | 438 (42.3) |
| Not sure | 727 (23.8) | 64 (25.9) | 97 (11.7) | 125 (12.1) |
| Probably would | 701 (23.0) | 48 (19.4) | 88 (10.6) | 63 (6.1) |
| Definitely would | 239 (7.8) | 17 (6.9) | 33 (4.0) | 17 (1.6) |

Go to A&E or visit another NHS service such as a walk-in centre or minor injuries unit

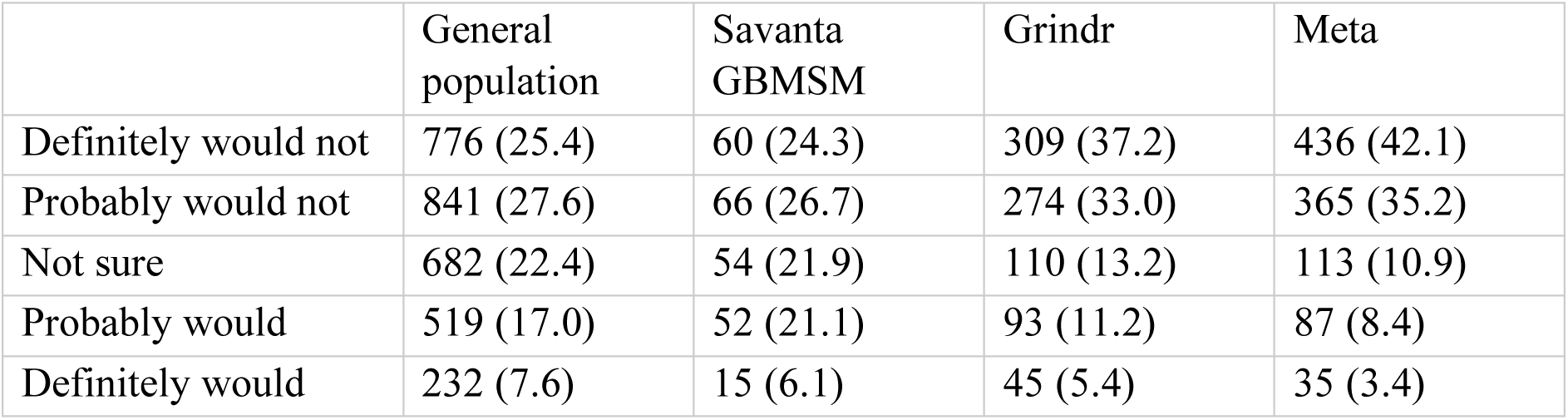

Call NHS 111 or 999 / ambulance service

|  | General population | Savanta GBMSM | Grindr | Meta |
| --- | --- | --- | --- | --- |
| Definitely would not | 246 (8.1) | 22 (8.9) | 145 (17.4) | 185 (17.9) |
| Probably would not | 358 (11.7) | 30 (12.1) | 153 (18.4) | 233 (22.5) |
| Not sure | 556 (18.2) | 36 (14.6) | 108 (13.0) | 118 (11.4) |
| Probably would | 1084 (35.5) | 95 (38.5) | 243 (29.2) | 283 (27.3) |
| Definitely would | 806 (26.4) | 64 (25.9) | 182 (21.9) | 217 (20.9) |

Visit a walk-in sexual health service

|  | General population | Savanta GBMSM | Grindr | Meta |
| --- | --- | --- | --- | --- |
| Definitely would not | 906 (29.7) | 50 (20.2) | 164 (19.7) | 207 (20.0) |
| Probably would not | 783 (25.7) | 56 (22.7) | 192 (23.1) | 203 (19.6) |
| Not sure | 727 (23.8) | 51 (20.6) | 125 (15.0) | 130 (12.5) |
| Probably would | 454 (14.9) | 65 (26.3) | 205 (24.7) | 262 (25.3) |
| Definitely would | 180 (5.9) | 25 (10.1) | 145 (17.4) | 234 (22.6) |

Call a sexual health service [fix below 6]

|  | General population | Savanta GBMSM | Grindr | Meta |
| --- | --- | --- | --- | --- |
| Definitely would not | 692 (22.7) | 27 (10.9) | 33 (4.0) | 25 (2.4) |
| Probably would not | 690 (22.6) | 38 (15.4) | 68 (8.2) | 57 (5.5) |
| Not sure | 697 (22.9) | 35 (14.2) | 60 (7.2) | 60 (5.8) |
| Probably would | 630 (20.7) | 87 (35.2) | 278 (33.5) | 320 (30.9) |
| Definitely would | 341 (11.2) | 60 (24.3) | 392 (47.2) | 574 (55.4) |

Let people you have recently been in close contact with know that you have symptoms

|  | General population | Savanta GBMSM | Grindr | Meta |
| --- | --- | --- | --- | --- |
| Definitely would not | 81 (2.7) | 8 (3.2) | 34 (4.1) | 32 (3.1) |
| Probably would not | 220 (7.2) | 20 (8.1) | 75 (9.0) | 90 (8.7) |
| Not sure | 495 (16.2) | 47 (19.0) | 152 (18.3) | 177 (17.1) |
| Probably would | 1185 (38.9) | 96 (38.9) | 298 (35.9) | 396 (38.2) |
| Definitely would | 1069 (35) | 76 (30.8) | 272 (32.7) | 341 (32.9) |

Search for information from official websites (e.g. NHS, GOV.UK)

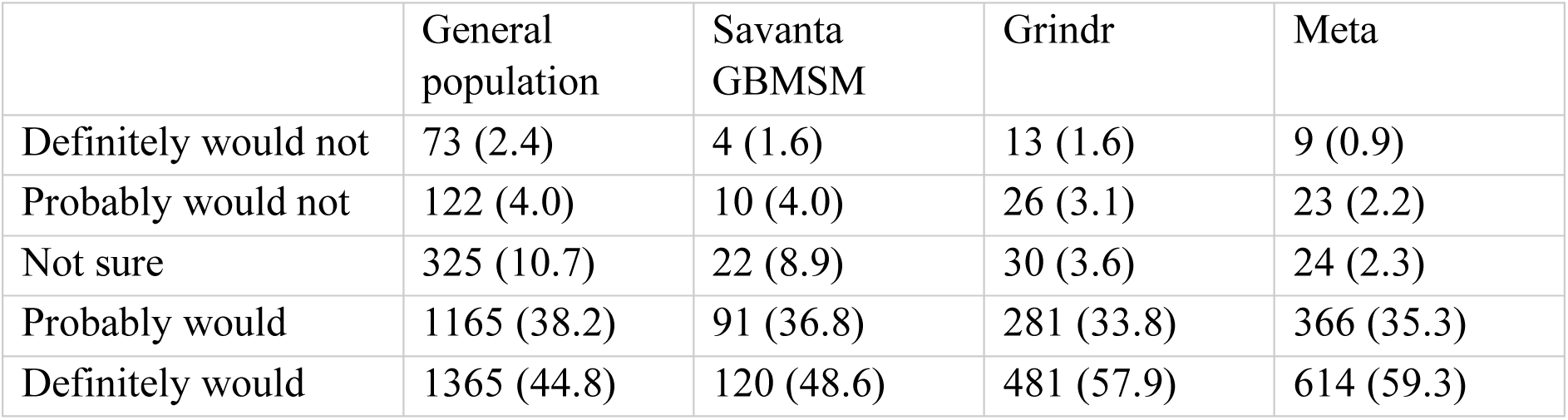

|  | General population | Savanta GBMSM | Grindr | Meta |
| --- | --- | --- | --- | --- |
| Definitely would not | 73 (2.4) | 4 (1.6) | 13 (1.6) | 9 (0.9) |
| Probably would not | 122 (4.0) | 10 (4.0) | 26 (3.1) | 23 (2.2) |
| Not sure | 325 (10.7) | 22 (8.9) | 30 (3.6) | 24 (2.3) |
| Probably would | 1165 (38.2) | 91 (36.8) | 281 (33.8) | 366 (35.3) |
| Definitely would | 1365 (44.8) | 120 (48.6) | 481 (57.9) | 614 (59.3) |

Search for information from other places, including blogs, social media sites (e.g. Facebook, Twitter, Instagram), online communities or other websites [fix below 9]

|  | General population | Savanta GBMSM | Grindr | Meta |
| --- | --- | --- | --- | --- |
| Definitely would not | 533 (17.5) | 49 (19.8) | 144 (17.3) | 135 (13.0) |
| Probably would not | 647 (21.2) | 54 (21.9) | 231 (27.8) | 277 (26.7) |
| Not sure | 647 (21.2) | 39 (15.8) | 113 (13.6) | 155 (15.0) |
| Probably would | 789 (25.9) | 70 (28.3) | 206 (24.8) | 301 (29.1) |
| Definitely would | 434 (14.2) | 35 (14.2) | 137 (16.5) | 168 (16.2) |

*New screen*

Imagine that tomorrow morning you develop an unexplained rash with blisters. You also learn that you have come into contact with someone who might have monkeypox.

Q13_other. Please state any other things you would do.

Open ended text

Nothing button

*Contact behaviour*

*New screen*

Imagine that tomorrow morning you develop an unexplained rash with blisters. You also learn that you have come into contact with someone who might have monkeypox.

ASK ALL

Q14 If **you** got these symptoms, in the following 21 days, realistically how much, if at all, would you:

RANDOMISE order

Come into skin-to-skin contact with other people

|  | General population | Savanta GBMSM | Grindr | Meta |
| --- | --- | --- | --- | --- |
| I would completely stop doing this | 1883 (61.7) | 153 (61.9) | 648 (78) | 858 (82.8) |
| Less than normal | 412 (13.5) | 41 (16.6) | 127 (15.3) | 135 (13.0) |
| Same as normal | 249 (8.2) | 21 (8.5) | 15 (1.8) | 12 (1.2) |
| More than normal | 107 (3.5) | 8 (3.2) | 3 (0.4) | 1 (0.1) |
| Not applicable, I wouldn't do this anyway | 280 (9.2) | 16 (6.5) | 22 (2.6) | 21 (2.0) |
| Don't know | 109 (3.6) | 7 (2.8) | 15 (1.8) | 9 (0.9) |
| Prefer not to say | 10 (0.3) | 1 (0.4) | 1 (0.1) | 0 (0.0) |

Have sexual contact (from kissing to intercourse) with other people

|  | General population | Savanta GBMSM | Grindr | Meta |
| --- | --- | --- | --- | --- |
| I would completely stop doing this | 1923 (63.0) | 171 (69.2) | 721 (86.8) | 922 (89.0) |
| Less than normal | 238 (7.8) | 20 (8.1) | 54 (6.5) | 61 (5.9) |
| Same as normal | 216 (7.1) | 15 (6.1) | 14 (1.7) | 7 (0.7) |
| More than normal | 113 (3.7) | 10 (4.0) | 1 (0.1) | 1 (0.1) |
| Not applicable, I wouldn't do this anyway | 451 (14.8) | 24 (9.7) | 27 (3.2) | 30 (2.9) |
| Don't know | 93 (3.0) | 6 (2.4) | 13 (1.6) | 13 (1.3) |
| Prefer not to say | 16 (0.5) | 1 (0.4) | 1 (0.1) | 2 (0.2) |

Have sex without using a condom

|  | General population | Savanta GBMSM | Grindr | Meta |
| --- | --- | --- | --- | --- |
| I would completely stop doing this | 1729 (56.7) | 155 (62.8) | 623 (75) | 788 (76.1) |
| Less than normal | 146 (4.8) | 17 (6.9) | 33 (4.0) | 37 (3.6) |
| Same as normal | 235 (7.7) | 15 (6.1) | 27 (3.2) | 29 (2.8) |
| More than normal | 124 (4.1) | 11 (4.5) | 3 (0.4) | 2 (0.2) |
| Not applicable, I wouldn't do this anyway | 680 (22.3) | 42 (17.0) | 123 (14.8) | 161 (15.5) |
| Don't know | 111 (3.6) | 7 (2.8) | 17 (2.0) | 14 (1.4) |
| Prefer not to say | 25 (0.8) | 0 (0.0) | 5 (0.6) | 5 (0.5) |

Share bedding, towels, or clothes with other people

|  | General population | Savanta GBMSM | Grindr | Meta |
| --- | --- | --- | --- | --- |
| I would completely stop doing this | 1839 (60.3) | 147 (59.5) | 604 (72.7) | 807 (77.9) |
| Less than normal | 286 (9.4) | 23 (9.3) | 62 (7.5) | 96 (9.3) |
| Same as normal | 279 (9.1) | 21 (8.5) | 24 (2.9) | 19 (1.8) |
| More than normal | 124 (4.1) | 12 (4.9) | 2 (0.2) | 1 (0.1) |
| Not applicable, I wouldn't do this anyway | 415 (13.6) | 41 (16.6) | 126 (15.2) | 104 (10.0) |
| Don't know | 98 (3.2) | 3 (1.2) | 12 (1.4) | 8 (0.8) |
| Prefer not to say | 9 (0.3) | 0 (0.0) | 1 (0.1) | 1 (0.1) |

Go to a crowded place

|  | General population | Savanta GBMSM | Grindr | Meta |
| --- | --- | --- | --- | --- |
| I would completely stop doing this | 1872 (61.4) | 141 (57.1) | 552 (66.4) | 663 (64.0) |
| Less than normal | 418 (13.7) | 50 (20.2) | 183 (22) | 297 (28.7) |
| Same as normal | 248 (8.1) | 23 (9.3) | 57 (6.9) | 39 (3.8) |
| More than normal | 123 (4.0) | 9 (3.6) | 1 (0.1) | 1 (0.1) |
| Not applicable, I wouldn't do this anyway | 275 (9.0) | 16 (6.5) | 25 (3.0) | 23 (2.2) |
| Don't know | 103 (3.4) | 8 (3.2) | 13 (1.6) | 12 (1.2) |
| Prefer not to say | 11 (0.4) | 0 (0.0) | 0 (0.0) | 1 (0.1) |

Help or provide care for a vulnerable person

|  | General population | Savanta GBMSM | Grindr | Meta |
| --- | --- | --- | --- | --- |
| I would completely stop doing this | 1674<br>(54.9) | 131<br>(53.0) | 516<br>(62.1) | 686 (66.2) |
| Less than normal | 304 (10.0) | 16 (6.5) | 81 (9.7) | 81 (7.8) |
| Same as normal | 255 (8.4) | 24 (9.7) | 23 (2.8) | 24 (2.3) |
| More than normal | 160 (5.2) | 14 (5.7) | 8 (1.0) | 6 (0.6) |
| Not applicable, I wouldn't do this anyway | 513 (16.8) | 50 (20.2) | 181<br>(21.8) | 217 (20.9) |
| Don't know | 134 (4.4) | 11 (4.5) | 22 (2.6) | 20 (1.9) |
| Prefer not to say | 10 (0.3) | 1 (0.4) | 0 (0.0) | 2 (0.2) |

Go to a public place where you may come into physical contact with (touch) someone else

|  | General population | Savanta GBMSM | Grindr | Meta |
| --- | --- | --- | --- | --- |
| I would completely stop doing this | 1818<br>(59.6) | 146<br>(59.1) | 525<br>(63.2) | 653 (63.0) |
| Less than normal | 469 (15.4) | 51 (20.6) | 236<br>(28.4) | 318 (30.7) |
| Same as normal | 249 (8.2) | 20 (8.1) | 33 (4.0) | 24 (2.3) |
| More than normal | 143 (4.7) | 9 (3.6) | 2 (0.2) | 2 (0.2) |
| Not applicable, I wouldn't do this anyway | 259 (8.5) | 11 (4.5) | 24 (2.9) | 24 (2.3) |
| Don't know | 104 (3.4) | 9 (3.6) | 10 (1.2) | 14 (1.4) |
| Prefer not to say | 8 (0.3) | 1 (0.4) | 1 (0.1) | 1 (0.1) |

*New screen*

Imagine that tomorrow morning you develop an unexplained rash with blisters. You also learn that you have come into contact with someone who might have monkeypox.

Q15 How many days after the start of these symptoms would you:

Please give an approximate number if you are unsure. If you would not do this, please put 0. Type your answer below

|  |  |  | General population | Savanta GBMSM | Grindr | Meta |
| --- | --- | --- | --- | --- | --- | --- |
| Have sexual contact (from kissing to intercourse) with someone else | OPEN TEXT ENTRY – number [cap at 100] | ASK ONLY to those who answer 1-4 for similar question above | N=2021, M=15.7, SD=15.3, range 0 to 100. | N=190, M=16.2, SD=14.4, range 0 to 100. | N=746, M=20.4, SD=15.8, range 0 to 100. | N=947, M=20.0, SD=12.3, range 0 to 100. |
| Prefer not to say |  |  | 469 (18.8) | 26 (12.0) | 44 (5.6) | 44 (4.4) |
| Have sex without using a condom | OPEN TEXT ENTRY – number [cap at 100] | ASK ONLY to those who answer 1-4 for similar question above | N=1807, M=15.7, SD=16.8, range 0 to 100. | N=175, M=16.8, SD=18.9, range 0 to 100. | N=649, M=21.2, SD=19.5, range 0 to 100. | N=821, M=21.3, SD=18.0, range 0 to 100. |
| Prefer not to say |  |  | 427 (19.1) | 23 (11.6) | 37 (5.4) | 35 (4.1) |

*Contact sharing*

*New screen*

People with suspected or confirmed monkeypox are being asked by public health officials to share contact details of people they have been in close contact with, so that they can be offered appropriate care and advice. Details of contacts are kept completely confidential.

ASK ALL

Q16 If **you** were asked for this information, would you try to share all the contact details of every …

RANDOMISE statements

Person who had been in your home (household members and visitors) in the last seven days

|  | General population | Savanta GBMSM | Grindr | Meta |
| --- | --- | --- | --- | --- |
| Definitely would not | 135 (4.4) | 14 (5.7) | 51 (6.1) | 56 (5.4) |
| Probably would not | 168 (5.5) | 19 (7.7) | 101 (12.2) | 118 (11.4) |
| Not sure | 392 (12.9) | 30 (12.1) | 97 (11.7) | 120 (11.6) |
| Probably would | 758 (24.9) | 60 (24.3) | 247 (29.7) | 325 (31.4) |
| Definitely would | 1552 (50.9) | 120 (48.6) | 331 (39.8) | 414 (40.0) |
| Prefer not to say | 45 (1.5) | 4 (1.6) | 4 (0.5) | 3 (0.3) |

Person you had sexual contact with (from kissing to intercourse) in the last seven days

|  | General population | Savanta GBMSM | Grindr | Meta |
| --- | --- | --- | --- | --- |
| Definitely would not | 169 (5.5) | 17 (6.9) | 49 (5.9) | 54 (5.2) |
| Probably would not | 162 (5.3) | 16 (6.5) | 59 (7.1) | 67 (6.5) |
| Not sure | 366 (12.0) | 27 (10.9) | 59 (7.1) | 83 (8.0) |
| Probably would | 629 (20.6) | 56 (22.7) | 236 (28.4) | 282 (27.2) |
| Definitely would | 1671 (54.8) | 127 (51.4) | 420 (50.5) | 545 (52.6) |
| Prefer not to say | 53 (1.7) | 4 (1.6) | 8 (1.0) | 5 (0.5) |

Place you had sexual contact with someone (from kissing to intercourse) in the last seven days

|  | General population | Savanta GBMSM | Grindr | Meta |
| --- | --- | --- | --- | --- |
| Definitely would not | 165 (5.4) | 17 (6.9) | 51 (6.1) | 52 (5) |
| Probably would not | 188 (6.2) | 18 (7.3) | 63 (7.6) | 72 (6.9) |
| Not sure | 415 (13.6) | 28 (11.3) | 72 (8.7) | 91 (8.8) |
| Probably would | 655 (21.5) | 64 (25.9) | 261 (31.4) | 306 (29.5) |
| Definitely would | 1559 (51.1) | 116 (47) | 375 (45.1) | 510 (49.2) |
| Prefer not to say | 68 (2.2) | 4 (1.6) | 9 (1.1) | 5 (0.5) |

Person you had skin-to-skin contact with (including hugging or kissing) in the last seven days

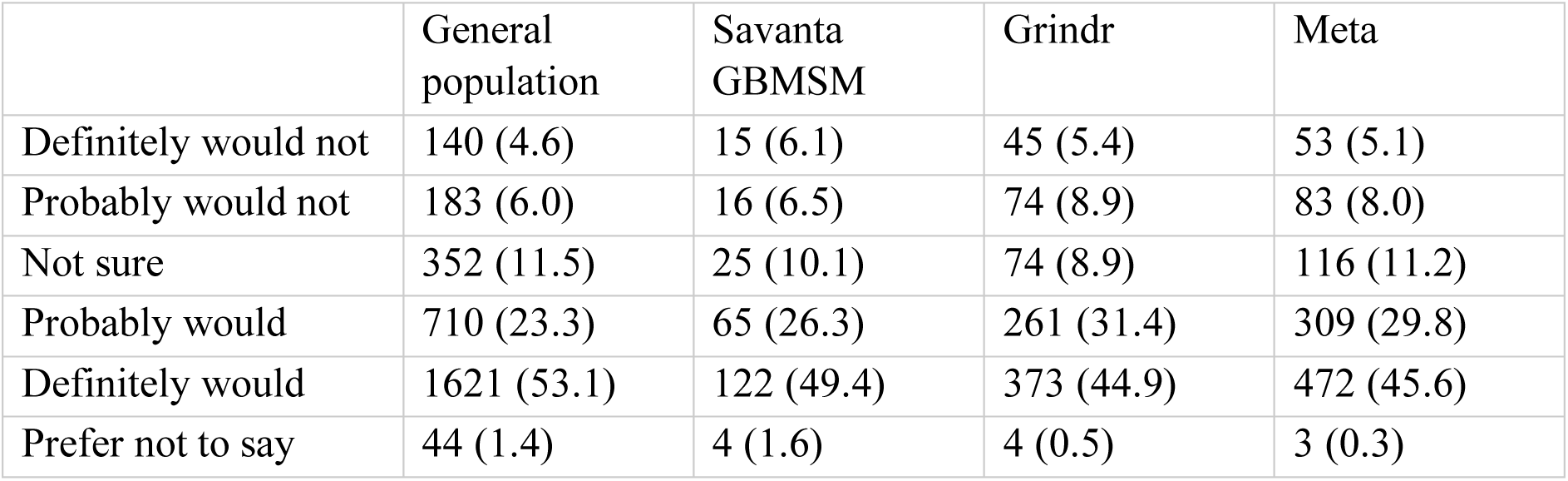

Person you had shared bedding, towels, or clothes with in the last seven days

|  | General population | Savanta GBMSM | Grindr | Meta |
| --- | --- | --- | --- | --- |
| Definitely would not | 148 (4.9) | 15 (6.1) | 48 (5.8) | 47 (4.5) |
| Probably would not | 180 (5.9) | 16 (6.5) | 57 (6.9) | 70 (6.8) |
| Not sure | 352 (11.5) | 26 (10.5) | 69 (8.3) | 81 (7.8) |
| Probably would | 746 (24.5) | 65 (26.3) | 236 (28.4) | 282 (27.2) |
| Definitely would | 1574 (51.6) | 122 (49.4) | 415 (49.9) | 551 (53.2) |
| Prefer not to say | 50 (1.6) | 3 (1.2) | 6 (0.7) | 5 (0.5) |

*Attitudes and beliefs*

*New screen*

ASK ALL

Q17 How much do you agree or disagree with the following statements:

RANDOMISE statements

I wouldn’t want to know the results of a monkeypox test

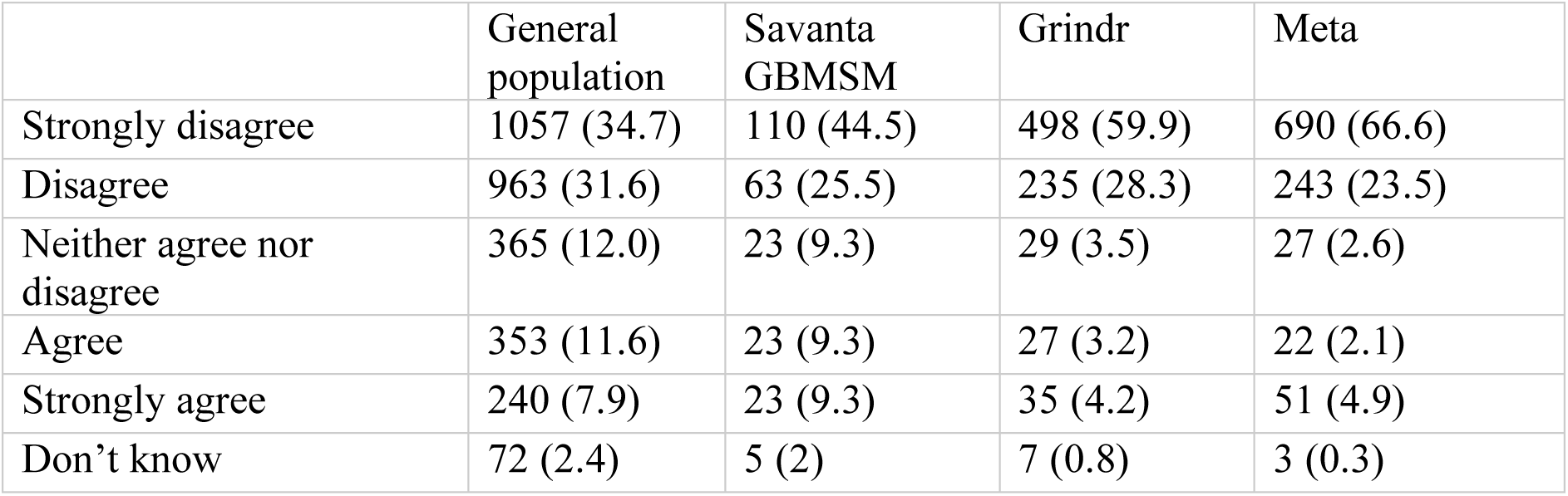

|  | General population | Savanta GBMSM | Grindr | Meta |
| --- | --- | --- | --- | --- |
| Strongly disagree | 1057 (34.7) | 110 (44.5) | 498 (59.9) | 690 (66.6) |
| Disagree | 963 (31.6) | 63 (25.5) | 235 (28.3) | 243 (23.5) |
| Neither agree nor disagree | 365 (12.0) | 23 (9.3) | 29 (3.5) | 27 (2.6) |
| Agree | 353 (11.6) | 23 (9.3) | 27 (3.2) | 22 (2.1) |
| Strongly agree | 240 (7.9) | 23 (9.3) | 35 (4.2) | 51 (4.9) |
| Don't know | 72 (2.4) | 5 (2) | 7 (0.8) | 3 (0.3) |

I would be worried what my friends or family would think about me if they thought I had monkeypox

|  | General population | Savanta GBMSM | Grindr | Meta |
| --- | --- | --- | --- | --- |
| Strongly disagree | 428 (14.0) | 35 (14.2) | 83 (10.0) | 125 (12.1) |
| Disagree | 624 (20.5) | 43 (17.4) | 114 (13.7) | 147 (14.2) |
| Neither agree nor disagree | 627 (20.6) | 42 (17.0) | 99 (11.9) | 104 (10.0) |
| Agree | 865 (28.4) | 73 (29.6) | 271 (32.6) | 325 (31.4) |
| Strongly agree | 402 (13.2) | 52 (21.1) | 263 (31.6) | 333 (32.1) |
| Don't know | 104 (3.4) | 2 (0.8) | 1 (0.1) | 2 (0.2) |

I would be worried about how colleagues / my employer would react if they thought I had monkeypox

|  | General population | Savanta GBMSM | Grindr | Meta |
| --- | --- | --- | --- | --- |
| Strongly disagree | 366 (12) | 31 (12.6) | 79 (9.5) | 99 (9.6) |
| Disagree | 508 (16.7) | 40 (16.2) | 95 (11.4) | 112 (10.8) |
| Neither agree nor disagree | 649 (21.3) | 37 (15.0) | 82 (9.9) | 109 (10.5) |
| Agree | 914 (30.0) | 67 (27.1) | 249 (30.0) | 309 (29.8) |
| Strongly agree | 462 (15.1) | 64 (25.9) | 316 (38.0) | 391 (37.7) |
| Don't know | 151 (5.0) | 8 (3.2) | 10 (1.2) | 16 (1.5) |

I don’t want to have a monkeypox test result on my medical record

I don't want to have a monkeypox test result on my medical record
|  | General population | Savanta GBMSM | Grindr | Meta |
| --- | --- | --- | --- | --- |
| Strongly disagree | 541 (17.7) | 50 (20.2) | 161 (19.4) | 229 (22.1) |
| Disagree | 801 (26.3) | 63 (25.5) | 193 (23.2) | 260 (25.1) |
| Neither agree nor disagree | 837 (27.4) | 58 (23.5) | 183 (22.0) | 205 (19.8) |
| Agree | 494 (16.2) | 42 (17.0) | 142 (17.1) | 151 (14.6) |
| Strongly agree | 257 (8.4) | 28 (11.3) | 142 (17.1) | 171 (16.5) |
| Don't know | 120 (3.9) | 6 (2.4) | 10 (1.2) | 20 (1.9) |

An effective way to prevent the spread of monkeypox is for people who have symptoms to contact healthcare services

|  | General population | Savanta GBMSM | Grindr | Meta |
| --- | --- | --- | --- | --- |
| Strongly disagree | 67 (2.2) | 7 (2.8) | 16 (1.9) | 19 (1.8) |
| Disagree | 131 (4.3) | 9 (3.6) | 25 (3.0) | 19 (1.8) |
| Neither agree nor disagree | 459 (15.0) | 24 (9.7) | 58 (7.0) | 64 (6.2) |
| Agree | 1185 (38.9) | 88 (35.6) | 311 (37.4) | 378 (36.5) |
| Strongly agree | 1105 (36.2) | 113 (45.7) | 408 (49.1) | 549 (53) |
| Don't know | 103 (3.4) | 6 (2.4) | 13 (1.6) | 7 (0.7) |

I would be willing to contact a sexual health clinic if I thought I had monkeypox symptoms or had come into contact with someone who had monkeypox

|  | General population | Savanta GBMSM | Grindr | Meta |
| --- | --- | --- | --- | --- |
| Strongly disagree | 149 (4.9) | 7 (2.8) | 21 (2.5) | 20 (1.9) |
| Disagree | 291 (9.5) | 20 (8.1) | 24 (2.9) | 17 (1.6) |
| Neither agree nor disagree | 590 (19.3) | 28 (11.3) | 36 (4.3) | 39 (3.8) |
| Agree | 1047 (34.3) | 73 (29.6) | 260 (31.3) | 247 (23.8) |
| Strongly agree | 800 (26.2) | 110 (44.5) | 481 (57.9) | 709 (68.4) |
| Don't know | 173 (5.7) | 9 (3.6) | 9 (1.1) | 4 (0.4) |

*New screen*

ASK ALL

Q18 How much do you agree or disagree with the following statements:

RANDOMISE statements

If I had monkeypox symptoms, I wouldn’t want to tell anyone as I don’t want to self-isolate

|  | General population | Savanta GBMSM | Grindr | Meta |
| --- | --- | --- | --- | --- |
| Strongly disagree | 1055 (34.6) | 79 (32.0) | 291 (35.0) | 374 (36.1) |
| Disagree | 949 (31.1) | 74 (30.0) | 322 (38.7) | 414 (40.0) |
| Neither agree nor disagree | 431 (14.1) | 33 (13.4) | 98 (11.8) | 119 (11.5) |
| Agree | 365 (12.0) | 32 (13.0) | 72 (8.7) | 78 (7.5) |
| Strongly agree | 174 (5.7) | 21 (8.5) | 36 (4.3) | 35 (3.4) |
| Don't know | 76 (2.5) | 8 (3.2) | 12 (1.4) | 16 (1.5) |

If I had monkeypox symptoms, I wouldn’t want to tell anyone so that others don’t have to self-isolate

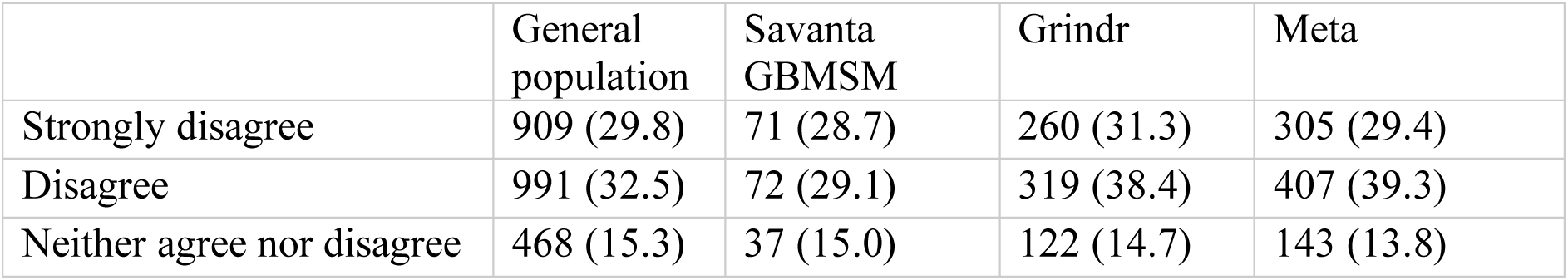
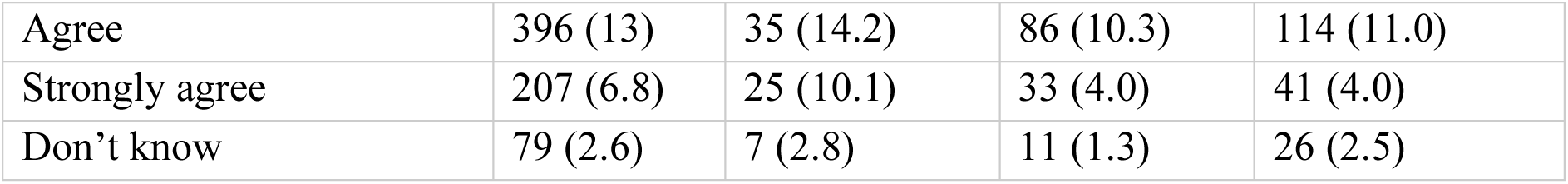

Most people would self-isolate if they were told to

|  | General population | Savanta GBMSM | Grindr | Meta |
| --- | --- | --- | --- | --- |
| Strongly disagree | 132 (4.3) | 14 (5.7) | 84 (10.1) | 90 (8.7) |
| Disagree | 561 (18.4) | 42 (17.0) | 233 (28.0) | 338 (32.6) |
| Neither agree nor disagree | 634 (20.8) | 51 (20.6) | 180 (21.7) | 222 (21.4) |
| Agree | 1127 (37.0) | 93 (37.7) | 239 (28.8) | 311 (30.0) |
| Strongly agree | 461 (15.1) | 32 (13.0) | 51 (6.1) | 36 (3.5) |
| Don't know | 135 (4.4) | 15 (6.1) | 44 (5.3) | 39 (3.8) |

I have the support I need to self-isolate for 21 days

|  | General population | Savanta GBMSM | Grindr | Meta |
| --- | --- | --- | --- | --- |
| Strongly disagree | 173 (5.7) | 26 (10.5) | 142 (17.1) | 132 (12.7) |
| Disagree | 348 (11.4) | 28 (11.3) | 185 (22.3) | 245 (23.6) |
| Neither agree nor disagree | 458 (15.0) | 41 (16.6) | 129 (15.5) | 127 (12.3) |
| Agree | 1062 (34.8) | 80 (32.4) | 226 (27.2) | 352 (34.0) |
| Strongly agree | 942 (30.9) | 66 (26.7) | 133 (16.0) | 168 (16.2) |
| Don't know | 67 (2.2) | 6 (2.4) | 16 (1.9) | 12 (1.2) |

An effective way to prevent the spread of monkeypox is for people who have tested positive to self-isolate

|  | General population | Savanta GBMSM | Grindr | Meta |
| --- | --- | --- | --- | --- |
| Strongly disagree | 51 (1.7) | 2 (0.8) | 17 (2.0) | 21 (2.0) |
| Disagree | 107 (3.5) | 13 (5.3) | 16 (1.9) | 34 (3.3) |
| Neither agree nor disagree | 361 (11.8) | 24 (9.7) | 45 (5.4) | 64 (6.2) |
| Agree | 1018 (33.4) | 68 (27.5) | 319 (38.4) | 362 (34.9) |
| Strongly agree | 1424 (46.7) | 132 (53.4) | 416 (50.1) | 543 (52.4) |
| Don't know | 89 (2.9) | 8 (3.2) | 18 (2.2) | 12 (1.2) |

*New screen*

ASK ALL

Q19 How much do you agree or disagree with the following statements: If I had to self-isolate because I had tested positive for monkeypox…

RANDOMISE statements

…I would lose touch with my friends and relatives

|  | General population | Savanta GBMSM | Grindr | Meta |
| --- | --- | --- | --- | --- |
| Strongly disagree | 567 (18.6) | 44 (17.8) | 151 (18.2) | 180 (17.4) |
| Disagree | 1039 (34.1) | 78 (31.6) | 320 (38.5) | 408 (39.4) |
| Neither agree nor disagree | 614 (20.1) | 47 (19.0) | 148 (17.8) | 180 (17.4) |
| Agree | 539 (17.7) | 52 (21.1) | 134 (16.1) | 200 (19.3) |
| Strongly agree | 225 (7.4) | 19 (7.7) | 61 (7.3) | 65 (6.3) |
| Don't know | 66 (2.2) | 7 (2.8) | 17 (2) | 3 (0.3) |

…it would have a severe impact on my family’s wellbeing

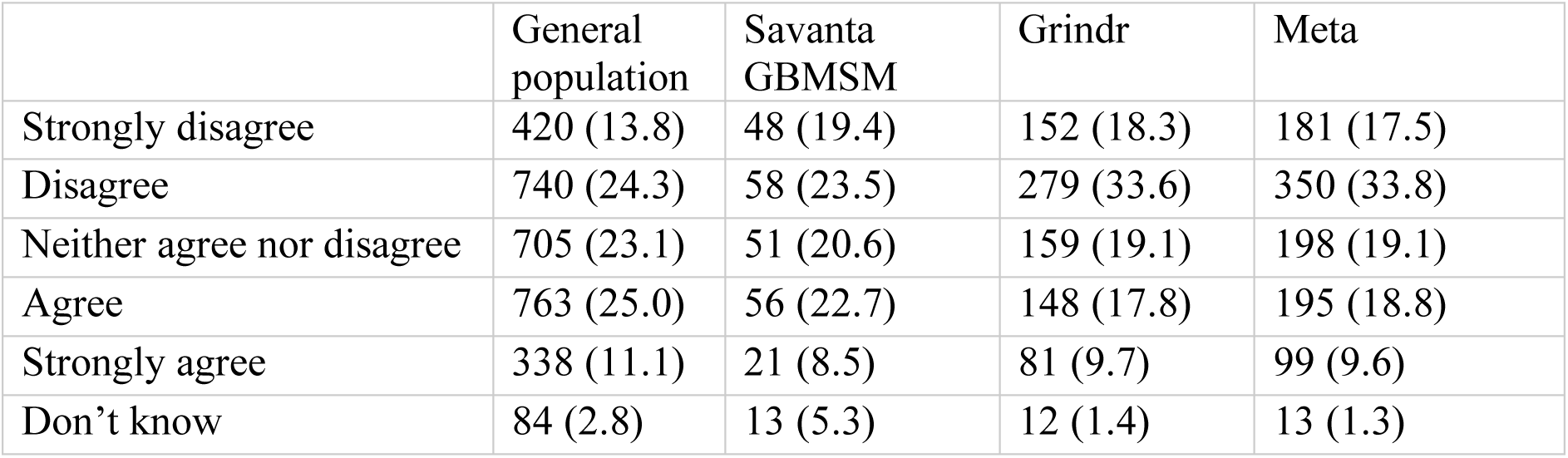

…it would have a negative impact on how much money I have

...it would have a negative impact on how much money I have
|  | General population | Savanta GBMSM | Grindr | Meta |
| --- | --- | --- | --- | --- |
| Strongly disagree | 498 (16.3) | 52 (21.1) | 132 (15.9) | 184 (17.8) |
| Disagree | 791 (25.9) | 62 (25.1) | 231 (27.8) | 357 (34.5) |
| Neither agree nor disagree | 566 (18.6) | 27 (10.9) | 99 (11.9) | 153 (14.8) |
| Agree | 694 (22.8) | 56 (22.7) | 162 (19.5) | 169 (16.3) |
| Strongly agree | 414 (13.6) | 43 (17.4) | 196 (23.6) | 165 (15.9) |
| Don't know | 87 (2.9) | 7 (2.8) | 11 (1.3) | 8 (0.8) |

…it would have a negative impact on my work

...it would have a negative impact on my work
|  | General population | Savanta GBMSM | Grindr | Meta |
| --- | --- | --- | --- | --- |
| Strongly disagree | 545 (17.9) | 52 (21.1) | 76 (9.1) | 98 (9.5) |
| Disagree | 664 (21.8) | 38 (15.4) | 136 (16.4) | 191 (18.4) |
| Neither agree nor disagree | 622 (20.4) | 49 (19.8) | 100 (12) | 136 (13.1) |
| Agree | 697 (22.9) | 55 (22.3) | 260 (31.3) | 325 (31.4) |
| Strongly agree | 405 (13.3) | 46 (18.6) | 251 (30.2) | 264 (25.5) |
| Don't know | 117 (3.8) | 7 (2.8) | 8 (1.0) | 22 (2.1) |

…I would miss out on events and activities that I want to attend

...I would miss out on events and activities that I want to attend
|  | General population | Savanta GBMSM | Grindr | Meta |
| --- | --- | --- | --- | --- |
| Strongly disagree | 340 (11.1) | 32 (13.0) | 77 (9.3) | 46 (4.4) |
| Disagree | 471 (15.4) | 33 (13.4) | 106 (12.8) | 107 (10.3) |
| Neither agree nor disagree | 607 (19.9) | 37 (15.0) | 97 (11.7) | 117 (11.3) |
| Agree | 1097 (36.0) | 84 (34.0) | 354 (42.6) | 457 (44.1) |
| Strongly agree | 460 (15.1) | 55 (22.3) | 185 (22.3) | 306 (29.5) |
| Don't know | 75 (2.5) | 6 (2.4) | 12 (1.4) | 3 (0.3) |

### Vaccination

*New screen*

People who have come into significant contact with someone who has tested positive for monkeypox are being asked to have a smallpox vaccine to reduce the risk of getting seriously ill. As monkeypox is caused by a virus similar to the one that causes smallpox, vaccines designed for smallpox are considered effective in preventing or reducing the severity of monkeypox.

Like all medicines, this vaccine can cause side effects, but not everyone gets them. The most common side effects are pain and itching at the injection site. Most side effects are mild and clear fully without any treatment within 7 days.

*New screen*

For each question, please select the answer that reflects your opinion. Do not worry if you do not know what the best answer might be or if you are not at all familiar with monkeypox or vaccination. We only ask that you try to give your answer based on what you think you know or what you would honestly decide to do in the situations described. If you are really not sure, please answer “don’t know”.

*New screen*

ASK ALL

Q20 How much do you agree or disagree with the following statements:

RANDOMISE statements

In general, vaccination is a good thing

|  | General population | Savanta GBMSM | Grindr | Meta |
| --- | --- | --- | --- | --- |
| Strongly disagree | 65 (2.1) | 9 (3.6) | 15 (1.8) | 6 (0.6) |
| Disagree | 119 (3.9) | 4 (1.6) | 5 (0.6) | 3 (0.3) |
| Neither agree nor disagree | 361 (11.8) | 21 (8.5) | 33 (4.0) | 15 (1.4) |
| Agree | 1019 (33.4) | 59 (23.9) | 189 (22.7) | 142 (13.7) |
| Strongly agree | 1400 (45.9) | 148 (59.9) | 584 (70.3) | 869 (83.9) |
| Don't know | 86 (2.8) | 6 (2.4) | 5 (0.6) | 1 (0.1) |

Most people like me will get a smallpox vaccination if advised

|  | General population | Savanta GBMSM | Grindr | Meta |
| --- | --- | --- | --- | --- |
| Strongly disagree | 82 (2.7) | 5 (2.0) | 24 (2.9) | 15 (1.4) |
| Disagree | 248 (8.1) | 14 (5.7) | 41 (4.9) | 43 (4.2) |
| Neither agree nor disagree | 533 (17.5) | 31 (12.6) | 93 (11.2) | 122 (11.8) |
| Agree | 1270 (41.6) | 111 (44.9) | 404 (48.6) | 470 (45.4) |
| Strongly agree | 717 (23.5) | 69 (27.9) | 223 (26.8) | 365 (35.2) |
| Don't know | 200 (6.6) | 17 (6.9) | 46 (5.5) | 21 (2.0) |

If I get a smallpox vaccination, I will be protected against monkeypox

|  | General population | Savanta GBMSM | Grindr | Meta |
| --- | --- | --- | --- | --- |
| Strongly disagree | 100 (3.3) | 9 (3.6) | 27 (3.2) | 6 (0.6) |
| Disagree | 385 (12.6) | 19 (7.7) | 71 (8.5) | 64 (6.2) |
| Neither agree nor disagree | 927 (30.4) | 52 (21.1) | 158 (19.0) | 139 (13.4) |
| Agree | 953 (31.2) | 102 (41.3) | 391 (47.1) | 599 (57.8) |
| Strongly agree | 250 (8.2) | 29 (11.7) | 117 (14.1) | 190 (18.3) |
| Don't know | 435 (14.3) | 36 (14.6) | 67 (8.1) | 38 (3.7) |

A smallpox vaccination could give me smallpox

|  | General population | Savanta GBMSM | Grindr | Meta |
| --- | --- | --- | --- | --- |
| Strongly disagree | 614 (20.1) | 70 (28.3) | 400 (48.1) | 668 (64.5) |
| Disagree | 931 (30.5) | 65 (26.3) | 277 (33.3) | 264 (25.5) |
| Neither agree nor disagree | 662 (21.7) | 38 (15.4) | 69 (8.3) | 51 (4.9) |
| Agree | 392 (12.9) | 32 (13.0) | 27 (3.2) | 17 (1.6) |
| Strongly agree | 136 (4.5) | 9 (3.6) | 8 (1.0) | 6 (0.6) |
| Don't know | 315 (10.3) | 33 (13.4) | 50 (6.0) | 30 (2.9) |

I might regret getting the smallpox vaccination if I later experienced side effects from it

|  | General population | Savanta GBMSM | Grindr | Meta |
| --- | --- | --- | --- | --- |
| Strongly disagree | 420 (13.8) | 48 (19.4) | 259 (31.2) | 410 (39.6) |
| Disagree | 825 (27.0) | 74 (30.0) | 350 (42.1) | 421 (40.6) |
| Neither agree nor disagree | 710 (23.3) | 43 (17.4) | 88 (10.6) | 121 (11.7) |
| Agree | 655 (21.5) | 50 (20.2) | 89 (10.7) | 61 (5.9) |
| Strongly agree | 268 (8.8) | 19 (7.7) | 31 (3.7) | 17 (1.6) |
| Don't know | 172 (5.6) | 13 (5.3) | 14 (1.7) | 6 (0.6) |

I am already immune to monkeypox

|  | General population | Savanta GBMSM | Grindr | Meta |
| --- | --- | --- | --- | --- |
| Strongly disagree | 676 (22.2) | 77 (31.2) | 269 (32.4) | 366 (35.3) |
| Disagree | 800 (26.2) | 51 (20.6) | 213 (25.6) | 251 (24.2) |
| Neither agree nor disagree | 532 (17.4) | 30 (12.1) | 106 (12.8) | 132 (12.7) |
| Agree | 228 (7.5) | 24 (9.7) | 57 (6.9) | 99 (9.6) |
| Strongly agree | 138 (4.5) | 13 (5.3) | 26 (3.1) | 44 (4.2) |
| Don't know | 676 (22.2) | 52 (21.1) | 160 (19.3) | 144 (13.9) |

I would be worried about experiencing side effects from a smallpox vaccination

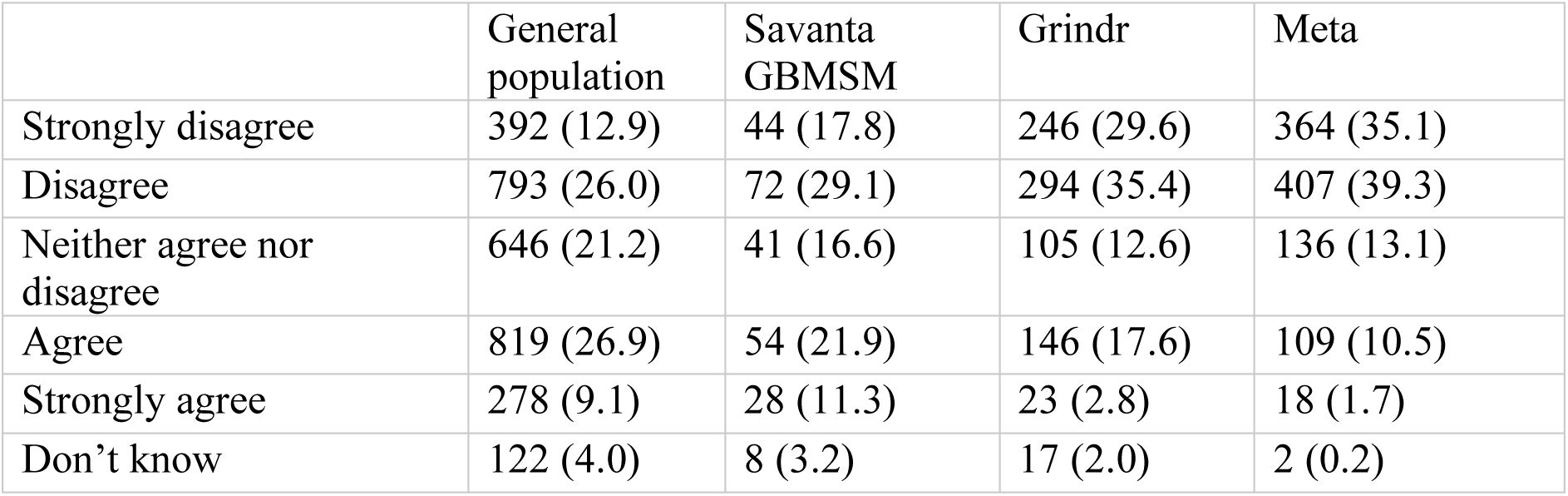

I would be worried that having a smallpox vaccine might make me infectious to others

|  | General population | Savanta GBMSM | Grindr | Meta |
| --- | --- | --- | --- | --- |
| Strongly disagree | 653 (21.4) | 74 (30.0) | 454 (54.6) | 693 (66.9) |
| Disagree | 1050 (34.4) | 72 (29.1) | 280 (33.7) | 274 (26.4) |
| Neither agree nor disagree | 605 (19.8) | 33 (13.4) | 46 (5.5) | 32 (3.1) |
| Agree | 378 (12.4) | 31 (12.6) | 18 (2.2) | 15 (1.4) |
| Strongly agree | 159 (5.2) | 18 (7.3) | 5 (0.6) | 7 (0.7) |
| Don't know | 205 (6.7) | 19 (7.7) | 28 (3.4) | 15 (1.4) |

People who are likely to come into high-risk contact with monkeypox should have a smallpox vaccine

|  | General population | Savanta GBMSM | Grindr | Meta |
| --- | --- | --- | --- | --- |
| Strongly disagree | 63 (2.1) | 5 (2.0) | 13 (1.6) | 9 (0.9) |
| Disagree | 158 (5.2) | 15 (6.1) | 8 (1.0) | 5 (0.5) |
| Neither agree nor disagree | 554 (18.2) | 32 (13) | 57 (6.9) | 32 (3.1) |
| Agree | 1134 (37.2) | 89 (36) | 256 (30.8) | 234 (22.6) |
| Strongly agree | 873 (28.6) | 89 (36) | 465 (56) | 745 (71.9) |
| Don't know | 268 (8.8) | 17 (6.9) | 32 (3.9) | 11 (1.1) |

*New screen*

ASK ALL

Q21 To the best of your knowledge, have you received a smallpox vaccine in 2022?

SINGLE CODE

Answer Options

|  | General population | Savanta GBMSM | Grindr | Meta |
| --- | --- | --- | --- | --- |
| Yes | 162 (5.3) | 26 (10.5) | 265 (31.9) | 422 (40.7) |
| No. I was offered the vaccine, but didn't want to have it | 116 (3.8) | 9 (3.6) | 11 (1.3) | 16 (1.5) |
| No, I haven't been offered the vaccine | 2609 (85.5) | 202 (81.8) | 536 (64.5) | 587 (56.7) |
| Don't know / can't remember | 163 (5.3) | 10 (4.0) | 19 (2.3) | 11 (1.1) |

*New screen*

ASK IF Q21=2,3,4

Q22 If you were advised by public health officials to have a smallpox vaccine **after coming into high-risk contact with someone who has monkeypox**, how likely would you be to have one?

SINGLE CODE

Answer Options

|  | General population, n=2888 | Savanta GBMSM, n=221 | Grindr, n=566 | Meta, n=614 |
| --- | --- | --- | --- | --- |
| Definitely would not | 115 (4.0) | 6 (2.7) | 16 (2.8) | 10 (1.6) |
| Probably would not | 139 (4.8) | 15 (6.8) | 13 (2.3) | 4 (0.7) |
| Not sure | 497 (17.2) | 22 (10.0) | 17 (3.0) | 14 (2.3) |
| Probably would | 814 (28.2) | 48 (21.7) | 95 (16.8) | 51 (8.3) |
| Definitely would | 1323 (45.8) | 130 (58.8) | 425 (75.1) | 535 (87.1) |

‡This question was not asked to people who had already had the smallpox vaccine in 2022, therefore total *ns* are different.

*New screen*

ASK IF Q21=2,3,4

Q23 Some people who have been in contact with someone who has monkeypox are being asked to self-isolate for 21 days. If having a smallpox vaccine means that you would have to self-isolate for less time, how likely would you be to get vaccinated?

SINGLE CODE

Answer Options

|  | General population, n=2888 | Savanta GBMSM, n=221 | Grindr, n=566 | Meta, n=614 |
| --- | --- | --- | --- | --- |
| Definitely would not | 114 (3.9) | 5 (2.3) | 17 (3.0) | 9 (1.5) |
| Probably would not | 128 (4.4) | 9 (4.1) | 17 (3.0) | 8 (1.3) |
| Not sure | 487 (16.9) | 29 (13.1) | 31 (5.5) | 19 (3.1) |
| Probably would | 822 (28.5) | 45 (20.4) | 92 (16.3) | 55 (9.0) |
| Definitely would | 1337 (46.3) | 133 (60.2) | 409 (72.3) | 523 (85.2) |

‡This question was not asked to people who had already had the smallpox vaccine in 2022, therefore total ns are different.

*New screen*

The smallpox vaccine is considered effective in preventing monkeypox or reducing the severity of monkeypox. Some people who are more likely to come into contact with monkeypox are being offered this vaccine.

ASK IF Q21=2,3,4

Q24 If you were offered a smallpox vaccine, how likely would you be to have one?

SINGLE CODE

Answer Options

|  | General population, n=2888 | Savanta GBMSM, n=221 | Grindr, n=566 | Meta, n=614 |
| --- | --- | --- | --- | --- |
| Definitely would not | 111 (3.8) | 4 (1.8) | 20 (3.5) | 12 (2.0) |
| Probably would not | 157 (5.4) | 12 (5.4) | 14 (2.5) | 5 (0.8) |
| Not sure | 518 (17.9) | 21 (9.5) | 20 (3.5) | 21 (3.4) |
| Probably would | 851 (29.5) | 64 (29.0) | 95 (16.8) | 64 (10.4) |
| Definitely would | 1251 (43.3) | 120 (54.3) | 417 (73.7) | 512 (83.4) |

‡This question was not asked to people who had already had the smallpox vaccine in 2022, therefore total ns are different.

*New screen*

ASK ALL

Q25. Do you have any further thoughts you would like to share with us about monkeypox, self-isolation, vaccines or anything else to do with monkeypox?

If you do not have anything to add, please put “None”. Type your answer below

OPEN END – require at least one alphabetic/numeric character

*New screen*

### Sociodemographic variables

Now we are going to ask some questions about you. This survey is anonymous. This means that nobody can link you to your answers.

*New screen*

ASK ALL

D1A In the past month, **have you personally**: Please select one option for each answer

Gone without enough food to eat

|  | General population | Savanta GBMSM | Grindr | Meta |
| --- | --- | --- | --- | --- |
| Never | 2006 (65.8) | 171 (69.2) | 670 (80.6) | 938 (90.5) |
| Sometimes | 663 (21.7) | 48 (19.4) | 121 (14.6) | 85 (8.2) |
| Often | 300 (9.8) | 21 (8.5) | 28 (3.4) | 8 (0.8) |
| Don't know | 63 (2.1) | 7 (2.8) | 7 (0.8) | 3 (0.3) |
| Prefer not to say | 18 (0.6) | 0 (0.0) | 5 (0.6) | 2 (0.2) |

Gone without an income

|  | General population | Savanta GBMSM | Grindr | Meta |
| --- | --- | --- | --- | --- |
| Never | 2014 (66) | 172 (69.6) | 653 (78.6) | 890 (85.9) |
| Sometimes | 600 (19.7) | 43 (17.4) | 105 (12.6) | 106 (10.2) |
| Often | 335 (11.0) | 24 (9.7) | 60 (7.2) | 34 (3.3) |
| Don't know | 69 (2.3) | 6 (2.4) | 2 (0.2) | 2 (0.2) |
| Prefer not to say | 32 (1.0) | 2 (0.8) | 11 (1.3) | 4 (0.4) |

Gone without fuel for heating or to cook food

|  | General population | Savanta GBMSM | Grindr | Meta |
| --- | --- | --- | --- | --- |
| Never | 2232 (73.2) | 179 (72.5) | 730 (87.8) | 978 (94.4) |
| Sometimes | 474 (15.5) | 38 (15.4) | 72 (8.7) | 48 (4.6) |
| Often | 255 (8.4) | 25 (10.1) | 22 (2.6) | 6 (0.6) |
| Don't know | 71 (2.3) | 2 (0.8) | 1 (0.1) | 2 (0.2) |
| Prefer not to say | 18 (0.6) | 3 (1.2) | 6 (0.7) | 2 (0.2) |

*New screen*

ASK ALL

D1B Last month, how difficult was it for you to cover your expenses and pay all your bills?

SINGLE CODE

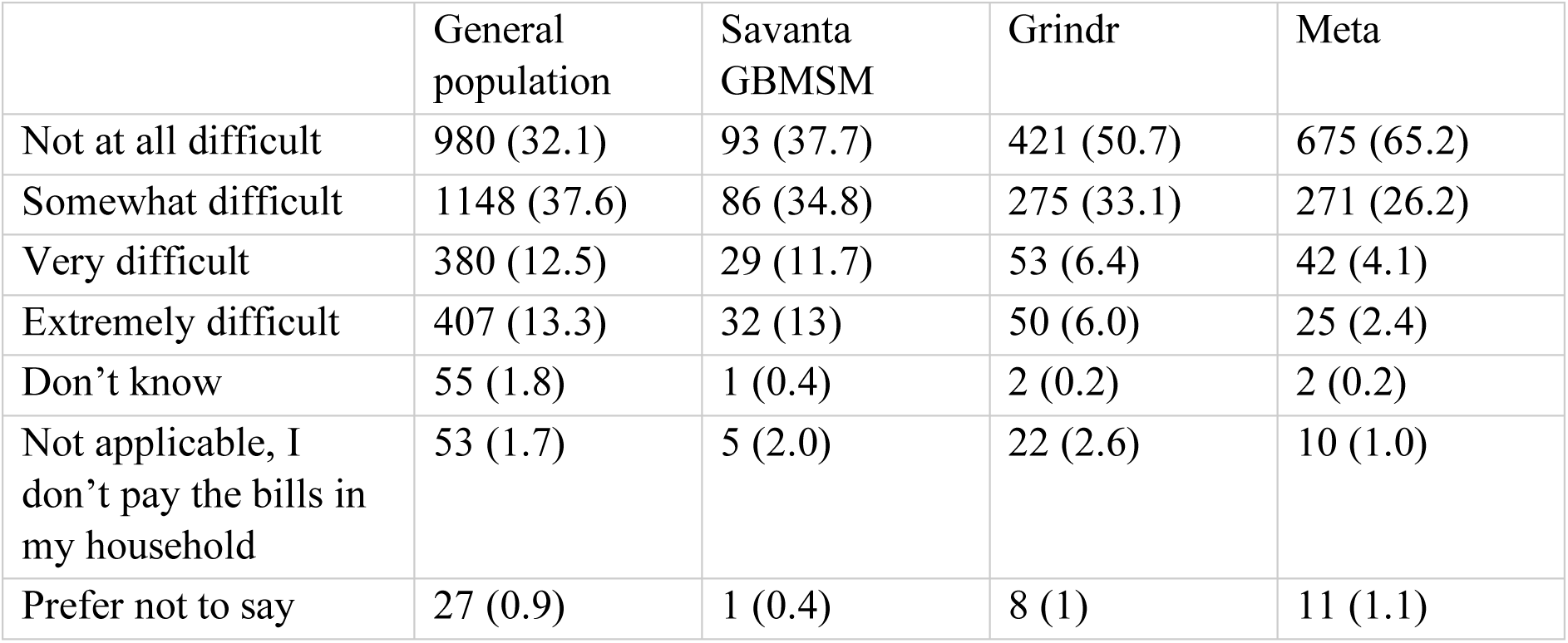

|  | General population | Savanta GBMSM | Grindr | Meta |
| --- | --- | --- | --- | --- |
| Not at all difficult | 980 (32.1) | 93 (37.7) | 421 (50.7) | 675 (65.2) |
| Somewhat difficult | 1148 (37.6) | 86 (34.8) | 275 (33.1) | 271 (26.2) |
| Very difficult | 380 (12.5) | 29 (11.7) | 53 (6.4) | 42 (4.1) |
| Extremely difficult | 407 (13.3) | 32 (13) | 50 (6.0) | 25 (2.4) |
| Don't know | 55 (1.8) | 1 (0.4) | 2 (0.2) | 2 (0.2) |
| Not applicable, I don't pay the bills in my household | 53 (1.7) | 5 (2.0) | 22 (2.6) | 10 (1.0) |
| Prefer not to say | 27 (0.9) | 1 (0.4) | 8 (1) | 11 (1.1) |

*New screen*

ASK ALL

D2 How many people currently live in your household?

Pl*ease include yourself and all adults and children – including those not related to you*

SINGLE CODE

|  | General population | Savanta GBMSM | Grindr | Meta |
| --- | --- | --- | --- | --- |
| I live alone | 663 (21.7) | 101 (40.9) | 358 (43.1) | 380 (36.7) |
| 2 | 1104 (36.2) | 90 (36.4) | 313 (37.7) | 531 (51.3) |
| 3 – 4 | 1055 (34.6) | 42 (17.0) | 131 (15.8) | 103 (9.9) |
| 5 – 6 | 204 (6.7) | 13 (5.3) | 25 (3.0) | 15 (1.4) |
| 7 + | 24 (0.8) | 1 (0.4) | 4 (0.5) | 7 (0.7) |

*New screen*

ASK ALL

D3 Are you the parent / guardian of any dependent children?

*Dependent children are those aged under 18 living in your household*.

SINGLE CODE

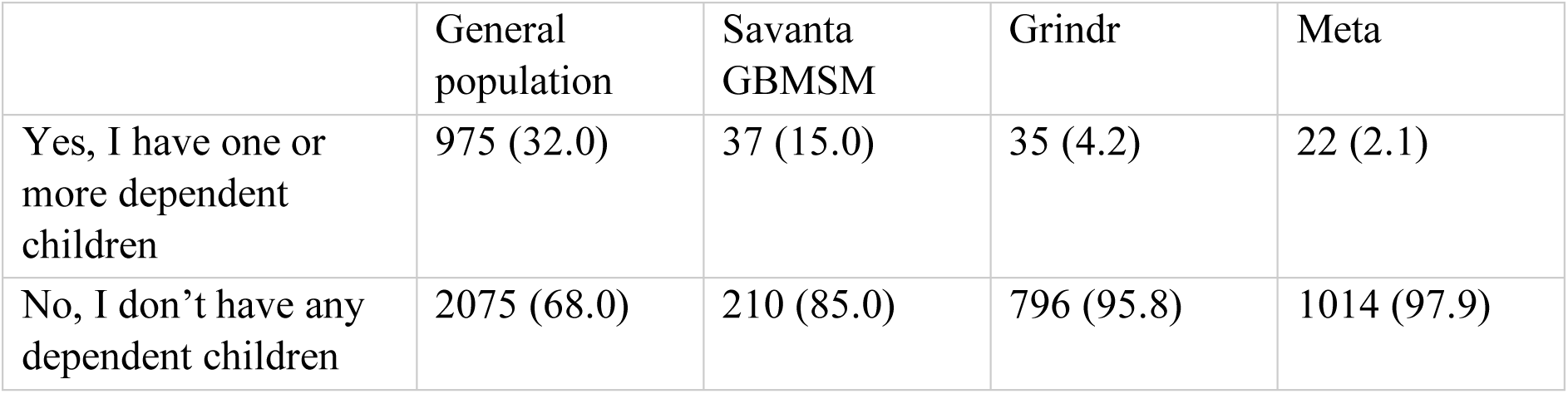

|  | General population | Savanta GBMSM | Grindr | Meta |
| --- | --- | --- | --- | --- |
| Yes, I have one or more dependent children | 975 (32.0) | 37 (15.0) | 35 (4.2) | 22 (2.1) |
| No, I don't have any dependent children | 2075 (68.0) | 210 (85.0) | 796 (95.8) | 1014 (97.9) |

*New screen*

ASK ALL

D4. Have you ever provided voluntary care for a family member or friend who needs support due to old age, physical illness, disability, mental health problems, or addiction?

Please select one option

SINGLE CODE

|  | General population | Savanta GBMSM | Grindr | Meta |
| --- | --- | --- | --- | --- |
| Yes, I currently do this | 623 (20.4) | 45 (18.2) | 158 (19.0) | 155 (15.0) |
| I have previously provided care for a friend/family member, but no longer do | 897 (29.4) | 68 (27.5) | 223 (26.8) | 315 (30.4) |
| I have never provided care for a family member/friend in this way | 1424 (46.7) | 121 (49) | 422 (50.8) | 541 (52.2) |
| Prefer not to say | 106 (3.5) | 13 (5.3) | 28 (3.4) | 25 (2.4) |

*New screen*

ASK ALL

D5 Do you currently have any pets that live in your home? MULTI CODE

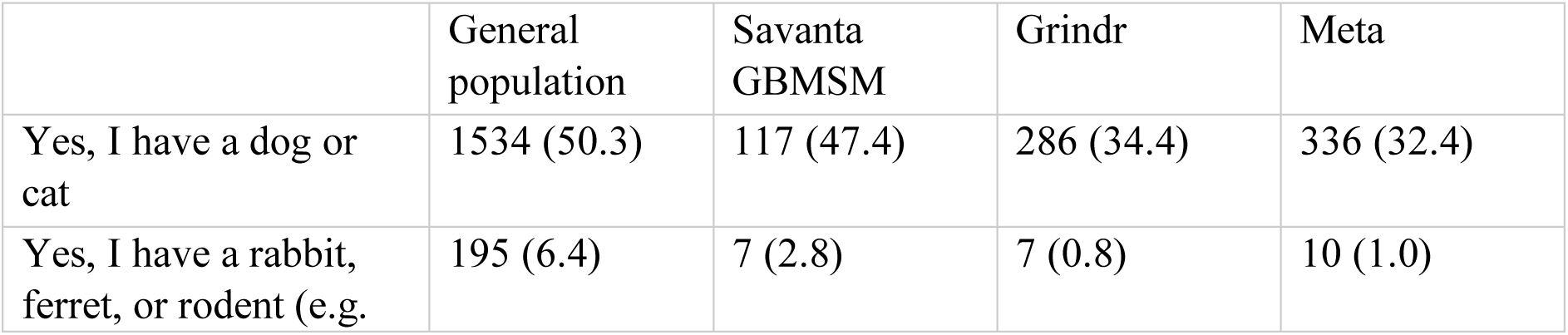
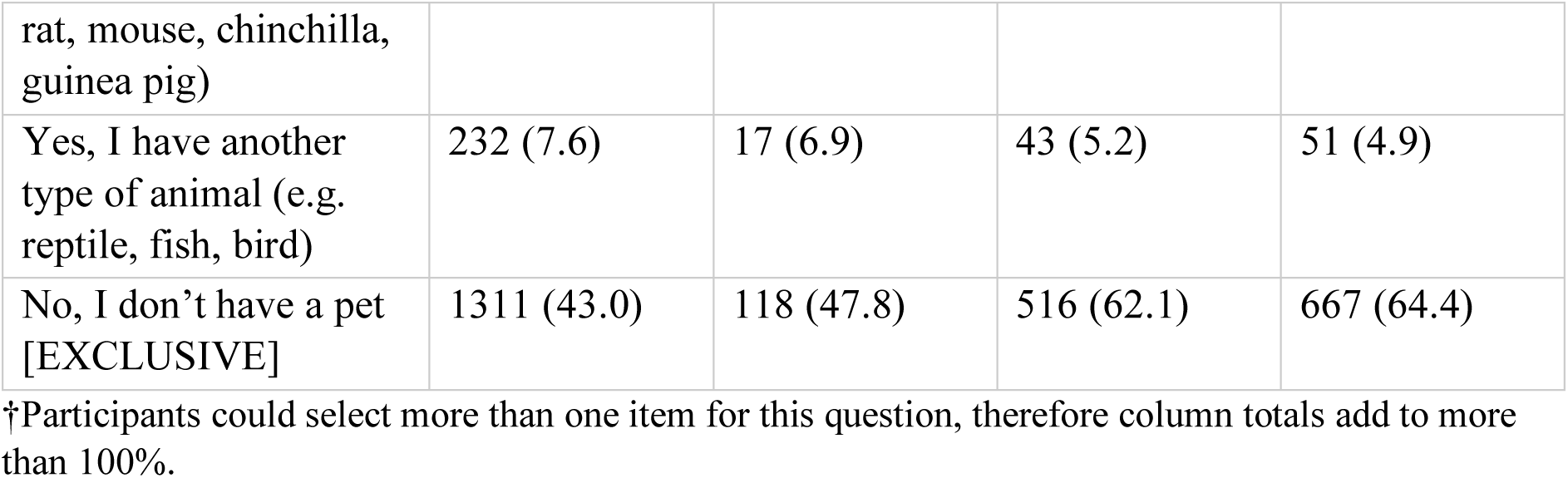

*New screen*

ASK ALL

D6 What is your employment status?

SINGLE CODE

|  | General population | Savanta GBMSM | Grindr | Meta |
| --- | --- | --- | --- | --- |
| Full time paid job (31+ hours) | 1163 (38.1) | 120 (48.6) | 529 (63.7) | 626 (60.4) |
| Part time paid job (<31 hours) | 471 (15.4) | 21 (8.5) | 58 (7.0) | 90 (8.7) |
| Doing paid work on a self-employed basis or within your own business | 102 (3.3) | 9 (3.6) | 82 (9.9) | 115 (11.1) |
| Student / On a government training programme (Nation Traineeship/Modern Apprenticeship) | 93 (3.0) | 7 (2.8) | 35 (4.2) | 32 (3.1) |
| Out of work | 120 (3.9) | 7 (2.8) | 31 (3.7) | 19 (1.8) |
| Looking after home / Homemaker | 184 (6.0) | 4 (1.6) | 3 (0.4) | 5 (0.5) |
| Retired | 707 (23.2) | 53 (21.5) | 57 (6.9) | 106 (10.2) |
| Disabled OR Long-term sick | 155 (5.1) | 21 (8.5) | 25 (3.0) | 28 (2.7) |
| Unpaid work for a business, community or voluntary organisation | 27 (0.9) | 1 (0.4) | 6 (0.7) | 11 (1.1) |
| Prefer not to say | 28 (0.9) | 4 (1.6) | 5 (0.6) | 4 (0.4) |

*New screen*

ASK IF D6= CODES 1,2, 3, 4, or 9

D6A Are you a **frontline health or social care worker**? Please include any voluntary work

*A frontline health or social care worker is someone who is directly involved in the care of patients or residents in long-stay care facilities (e.g. a care home), who has face-to-face contact with patients or clients, or who works in a laboratory, pathology or mortuary.*

SINGLE CODE

FIX ORDER

|  | General population, n=1856 | Savanta GBMSM, n=158 | Grindr, n=710 | Meta, n=874 |
| --- | --- | --- | --- | --- |
| Yes | 355 (19.1) | 23 (14.6) | 94 (13.2) | 125 (14.3) |
| No | 1474 (79.4) | 135 (85.4) | 613 (86.3) | 747 (85.5) |
| Prefer not to say | 27 (1.5) | 0 (0.0) | 3 (0.4) | 2 (0.2) |

‡This question was only asked to those who reported that they were in full-time, part-time, self-employment, a student, or did unpaid or voluntary work, therefore total *ns* are different.

*New screen*

ASK IF D6= CODES 1, 2 OR 3

D6B If you needed to, are you able to work from home?

SINGLE CODE

|  | General population, n=1736 | Savanta GBMSM, n=150 | Grindr, n=669 | Meta, n=831 |
| --- | --- | --- | --- | --- |
| Yes – full time | 632 (36.4) | 52 (34.7) | 209 (31.2) | 294 (35.4) |
| Yes – most of the time | 260 (15.0) | 26 (17.3) | 136 (20.3) | 196 (23.6) |
| Yes – but I have to go into my place of work once or twice a week | 142 (8.2) | 13 (8.7) | 67 (10.0) | 111 (13.4) |
| No – I cannot do my job from home | 702 (40.4) | 59 (39.3) | 257 (38.4) | 230 (27.7) |

‡This question was only asked to those who reported that they were in full-time, part-time, or self-employment, therefore total *ns* are different.

*New screen*

ASK ALL

D7 What is the highest level of educational qualification you have received?

SINGLE CODE

|  | General population | Savanta GBMSM | Grindr | Meta |
| --- | --- | --- | --- | --- |
| PhD/Doctorate | 43 (1.4) | 7 (2.8) | 41 (4.9) | 79 (7.6) |
| Masters | 213 (7.0) | 22 (8.9) | 204 (24.5) | 325 (31.4) |
| Bachelor's Degree or equivalent (Such as a NVQ level 5) | 729 (23.9) | 59 (23.9) | 291 (35.0) | 379 (36.6) |
| Higher education (Such as a HND or a NVQ level 4) | 348 (11.4) | 29 (11.7) | 106 (12.8) | 89 (8.6) |
| A level or equivalent (Such as Scottish Highers or NVQ level 3) | 666 (21.8) | 58 (23.5) | 110 (13.2) | 88 (8.5) |
| GCSE and below (Such as O level or an RSA Diploma) | 747 (24.5) | 53 (21.5) | 57 (6.9) | 63 (6.1) |
| Other qualifications (Such as NVQ level 1) | 119 (3.9) | 12 (4.9) | 11 (1.3) | 11 (1.1) |
| No qualifications | 185 (6.1) | 7 (2.8) | 11 (1.3) | 2 (0.2) |

*New screen*

ASK ALL

D8 Which of the following categories would best describe your ethnicity?

SINGLE CODE

|  | General population | Savanta GBMSM | Grindr | Meta |
| --- | --- | --- | --- | --- |
| **White** |  |  |  |  |
| British/English/Welsh/Scottish/Northern Irish | 2649 (86.9) | 214 (86.6) | 629 (75.7) | 799 (77.1) |
| Irish | 38 (1.2) | 5 (2.0) | 26 (3.1) | 31 (3.0) |
| Gypsy, Traveller or Irish Traveller | 7 (0.2) | 0 (0.0) | 1 (0.1) | 1 (0.1) |
| Any other White background | 71 (2.3) | 13 (5.3) | 90 (10.8) | 131 (12.6) |
| **Mixed/ Multiple ethnic groups** |  |  |  |  |
| White and Black Caribbean | 18 (0.6) | 1 (0.4) | 7 (0.8) | 4 (0.4) |
| White and Black African | 11 (0.4) | 4 (1.6) | 1 (0.1) | 3 (0.3) |
| White and Asian | 24 (0.8) | 1 (0.4) | 3 (0.4) | 9 (0.9) |
| Any other Mixed/ Multiple ethnic background | 16 (0.5) | 1 (0.4) | 12 (1.4) | 17 (1.6) |
| **Asian or Asian British** |  |  |  |  |
| Indian | 41 (1.3) | 4 (1.6) | 20 (2.4) | 1 (0.1) |
| Pakistani | 50 (1.6) | 0 (0.0) | 2 (0.2) | 0 (0.0) |
| Bangladeshi | 12 (0.4) | 0 (0.0) | 1 (0.1) | 1 (0.1) |
| Chinese | 14 (0.5) | 1 (0.4) | 7 (0.8) | 7 (0.7) |
| Any other Asian background | 12 (0.4) | 0 (0.0) | 4 (0.5) | 5 (0.5) |
| **Black or Black British** |  |  |  |  |
| African | 26 (0.9) | 1 (0.4) | 5 (0.6) | 2 (0.2) |
| Caribbean | 32 (1.0) | 0 (0.0) | 8 (1.0) | 4 (0.4) |
| Any other Black/ African/ Caribbean background | 3 (0.1) | 1 (0.4) | 1 (0.1) | 1 (0.1) |
| **Other ethnic group** |  |  |  |  |
| Arab | 7 (0.2) | 0 (0.0) | 3 (0.4) | 1 (0.1) |
| Other | 9 (0.3) | 1 (0.4) | 6 (0.7) | 8 (0.8) |
| **Don't know / Prefer not to say** |  |  |  |  |
| Don't know | 2 (0.1) | 0 (0.0) | 0 (0.0) | 1 (0.1) |
| Prefer not to say | 8 (0.3) | 0 (0.0) | 5 (0.6) | 10 (1.0) |

*New screen*

ASK ALL

D9 Do you, or anyone else in your household have any long-standing illness, disability or infirmity?

Please select all that apply MULTICODE

|  | General population | Savanta GBMSM | Grindr | Meta |
| --- | --- | --- | --- | --- |
| Yes, I do | 843 (27.6) | 80 (32.4) | 212 (25.5) | 305 (29.4) |
| Yes, another household member does | 391 (12.8) | 21 (8.5) | 88 (10.6) | 115 (11.1) |
| No | 1879 (61.6) | 149 (60.3) | 538 (64.7) | 634 (61.2) |
| Prefer not to say | 62 (2) | 4 (1.6) | 10 (1.2) | 14 (1.4) |

†Participants could select more than one item for this question, therefore column totals add to more than 100%.

*New screen*

ASK IF D9=1 (Yes, I do)

D9A Do you have any of the following health conditions?

Please select all that apply

MULTICODE

FIX ORDER

|  | General population, n=843 | Savanta GBMSM, n=80 | Grindr, n=212 | Meta, n=305 |
| --- | --- | --- | --- | --- |
| Diabetes | 184 (21.8) | 21 (26.3) | 38 (17.9) | 49 (16.1) |
| Any type of cancer | 57 (6.8) | 4 (5.0) | 10 (4.7) | 14 (4.6) |
| Heart conditions | 116 (13.8) | 8 (10.0) | 24 (11.3) | 42 (13.8) |
| Respiratory conditions (e.g. chronic obstructive pulmonary disease [COPD], asthma) | 191 (22.7) | 15 (18.8) | 27 (12.7) | 59 (19.3) |
| Mental health | 317 (37.6) | 34 (42.5) | 84 (39.6) | 102 (33.4) |
| A skin condition (e.g. atopic dermatitis / eczema) | 114 (13.5) | 9 (11.3) | 25 (11.8) | 46 (15.1) |
| A condition that makes you much more likely to get infections (e.g. SCID, homozygous sickle cell) | 20 (2.4) | 0 (0.0) | 7 (3.3) | 7 (2.3) |
| A weakened immune system | 110 (13) | 8 (10.0) | 24 (11.3) | 38 (12.5) |
| Are taking medicine that weakens your immune system (e.g. steroid tablets, chemotherapy, or antiretroviral medications) | 62 (7.4) | 4 (5.0) | 15 (7.1) | 24 (7.9) |
| HIV/AIDS | 6 (0.7) | 12 (15.0) | 52 (24.5) | 117 (38.4) |
| Other, please specify | 183 (21.7) | 11 (13.8) | 48 (22.6) | 60 (19.7) |
| Prefer not to say | 46 (5.5) | 5 (6.3) | 11 (5.2) | 8 (2.6) |

†Participants could select more than one item for this question, therefore column totals add to more than 100%.
‡This question was only asked to those who reported that had a chronic illness, therefore total *ns* are different.

*New screen*

ASK ALL

D9B Are you pregnant?

SINGLE CODE

|  | General population | Savanta GBMSM | Grindr | Meta |
| --- | --- | --- | --- | --- |
| Yes | 38 (1.2) | 1 (0.4) | 0 (0.0) | 0 (0.0) |
| No | 3001 (98.4) | 245 (99.2) | 830 (99.9) | 1035 (99.9) |
| Prefer not to say | 11 (0.4) | 1 (0.4) | 1 (0.1) | 1 (0.1) |

*New screen*

ASK ALL

D9C Have you ever taken PrEP (pre-exposure prophylaxis) for HIV?

SINGLE CODE

|  | General population | Savanta GBMSM | Grindr | Meta |
| --- | --- | --- | --- | --- |
| Yes | 42 (1.4) | 28 (11.3) | 402 (48.4) | 487 (47.0) |
| No | 2993 (98.1) | 216 (87.4) | 427 (51.4) | 547 (52.8) |
| Prefer not to say | 15 (0.5) | 3 (1.2) | 2 (0.2) | 2 (0.2) |

*New screen*

ASK ALL

D10 To the best of your knowledge, have you received a vaccine for…

Hepatitis A

|  | General population | Savanta GBMSM | Grindr | Meta |
| --- | --- | --- | --- | --- |
| Yes | 531 (17.4) | 87 (35.2) | 549 (66.1) | 773 (74.6) |
| No. I was offered the vaccine, but didn't want to have it | 128 (4.2) | 12 (4.9) | 7 (0.8) | 4 (0.4) |
| No, I haven't been offered the vaccine | 1579 (51.8) | 94 (38.1) | 129 (15.5) | 125 (12.1) |
| Don't know / can't remember | 812 (26.6) | 54 (21.9) | 146 (17.6) | 134 (12.9) |

Smallpox (before 2022)

|  | General population | Savanta GBMSM | Grindr | Meta |
| --- | --- | --- | --- | --- |
| Yes | 512 (16.8) | 51 (20.6) | 144 (17.3) | 218 (21) |
| No. I was offered the vaccine, but didn't want to have it | 117 (3.8) | 12 (4.9) | 5 (0.6) | 7 (0.7) |
| No, I haven't been offered the vaccine | 1666 (54.6) | 133 (53.8) | 477 (57.4) | 595 (57.4) |
| Don't know / can't remember | 755 (24.8) | 51 (20.6) | 205 (24.7) | 216 (20.8) |

Covid-19 (two doses or more)

|  | General population | Savanta GBMSM | Grindr | Meta |
| --- | --- | --- | --- | --- |
| Yes | 2557 (83.8) | 218 (88.3) | 796 (95.8) | 1023 (98.7) |
| No, I was offered the vaccine, but didn't want to have it | 242 (7.9) | 16 (6.5) | 27 (3.2) | 11 (1.1) |
| No, I haven't been offered the vaccine | 189 (6.2) | 9 (3.6) | 5 (0.6) | 0 (0.0) |
| Don't know / can't remember | 62 (2.0) | 4 (1.6) | 3 (0.4) | 2 (0.2) |

*New screen*

ASK ALL

D11 What is your marital status?

SINGLE CODE

|  | General population | Savanta GBMSM | Grindr | Meta |
| --- | --- | --- | --- | --- |
| Single, never married | 741 (24.3) | 123 (49.8) | 470 (56.6) | 402 (38.8) |
| Married / civil partnership | 1382 (45.3) | 62 (25.1) | 149 (17.9) | 290 (28) |
| Separated | 69 (2.3) | 3 (1.2) | 29 (3.5) | 21 (2.0) |
| Divorced | 319 (10.5) | 14 (5.7) | 57 (6.9) | 39 (3.8) |
| Widowed | 136 (4.5) | 1 (0.4) | 5 (0.6) | 24 (2.3) |
| Partnered / in a relationship | 393 (12.9) | 43 (17.4) | 111 (13.4) | 251 (24.2) |
| Prefer not to say | 10 (0.3) | 1 (0.4) | 10 (1.2) | 9 (0.9) |

*New screen*

The following questions are sensitive. You don’t need to answer if you don’t want to. If you don’t want to answer, please select “prefer not to say”.

*New screen*

ASK ALL

D12A How many **male** sexual partners have you had…?

*By “sexual partner” we mean any form of genital contact, including touching, kissing, and intercourse*.

Please give an approximate number if you are unsure. If you have not had any male sexual partners in this time, please put 0.

Type your answer below

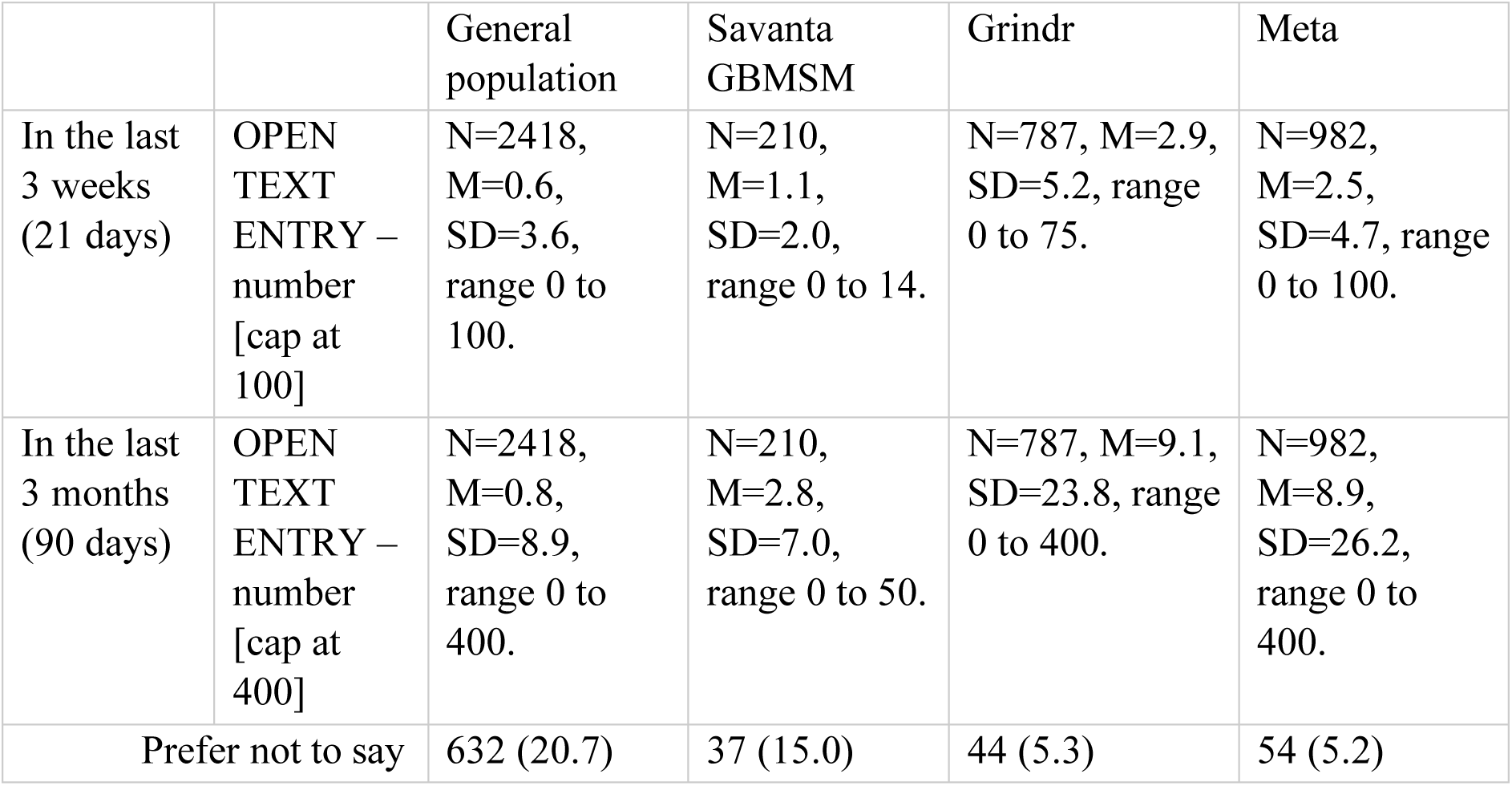

*New screen*

ASK ALL

D12B How many **female** sexual partners have you had…?

*By “sexual partner” we mean any form of genital contact, including touching, kissing, and intercourse*.

Please give an approximate number if you are unsure. If you have not had any female sexual partners in this time, please put 0.

Type your answer below

|  |  | General population | Savanta GBMSM | Grindr | Meta |
| --- | --- | --- | --- | --- | --- |
| In the last 3 weeks (21 days) | OPEN TEXT ENTRY – number [cap at 100] | N=2442, M=0.4, SD=3.1, range 0 to 100. | N=207, M=0.2, SD=0.8, range 0 to 100. | N=792, M=0.1, SD=0.6, range 0 to 10. | N=992, M=0.0, SD=0.4, range 0 to 9. |
| In the last 3 months (90 days) | OPEN TEXT ENTRY – number [cap at 400] | N=2442, M=0.8, SD=9.2, range 0 to 400. | N=207, M=2.2, SD=27.8, range 0 to 400. | N=792, M=0.2, SD=1.6, range 0 to 30. | N=992, M=0.1, SD=1.0, range 0 to 28. |
| Prefer not to say |  | 608 (19.9) | 40 (16.2) | 39 (4.7) | 44 (4.2) |

*New screen*

Thanks for completing the survey. If you would like any more information on monkeypox, please see https://www.nhs.uk/conditions/monkeypox/.

Please press the Finish button to complete the survey.

Many thanks for completing this survey, your views are extremely important to us. If you have any further comments or feedback, please use the box below.

## Supplementary materials 2. Data preparation

Some participants (3.8%, n=196) did not provide their postcode and were therefore missing data about region and index of multiple deprivation. While individual items about financial hardship had small amounts of missing data (2.0% to 3.8%, n=105 to 197), as items were summed to give a scale, missingness on the resulting scale would have been higher (7.9%, n=410). There were no apparent patterns to the missing data. We used multiple imputation to impute missing values for region, index of multiple deprivation and financial hardship items. Socio-demographic variables (age, occupation of highest earner, number of people in household, having a dependent child, employment status, education, ethnicity, marital status, and region, index of multiple deprivation and financial hardship where available) were used to predict missing values using a Markov Chain Monte Carlo (MCMC) method using linear regressions with ten iterations, to give five imputations.

We computed a single variable to indicate perceived susceptibility and severity from three items. For these items, “don’t know” was coded as the mid-point on the scale. We then multiplied responses together (range 1 to 125) and took the cubic root to give a scale from 1 (lowest perceived susceptibility and severity) to 5 (highest perceived susceptibility and severity). We also asked participants how much they agreed that they were already immune to mpox. This item was recoded into a binary item to show perceived immunity to mpox (strongly agree and agree, vs neither agree nor disagree, disagree, strongly disagree, and don’t know).

To create a single variable for perceived knowledge, we used three items (Cronbach’s α=0.74). Responses for these items were given on a five-point scale from “strongly disagree” to “strongly agree” We recoded each item as a binary item (strongly agree and agree [1], vs neither agree nor disagree, disagree, strongly disagree, and don’t know [0]), then summed responses to give a scale from 0 (lowest perceived knowledge) to 3 (highest perceived knowledge).

For knowledge about mpox symptoms, for each symptom selected, we coded participants as being correct (1, symptom listed on NHS mpox website) or incorrect (0, symptom not listed on NHS mpox website). Scores were then summed to give a scale from 0 (no selected symptoms were mpox symptoms) to 4 (all selected symptoms were mpox symptoms); responses of “don’t know” were coded as 0. A single variable denoting understanding of transmission was computed. It is unknown whether mpox can be caught from pet animals (but pet animals can catch mpox from humans (1)), therefore we did not include this item in analyses. We coded answers as correct (1) or incorrect (0; answers of “don’t know” coded as incorrect), and summed items to give a score from 0 (lowest knowledge about transmission) to 6 (highest knowledge about transmission).

To reduce the number of variables included in regression models, we used dimension reduction techniques. We conducted principal components analysis, using direct oblimin rotation as items may have been correlated, on items potentially associated with self-isolation (ten items), help seeking (six items), and vaccination (eight items) separately. The number of factors was determined using a scree plot. The item with the highest loading on to a component was then included in regression analyses. Results of the principal components analyses are reported in the supplementary materials 3.

## Supplementary materials 3. Results of principal components analyses

### Self-isolation

A scree plot suggested three components underlying factors potentially associated with self-isolation. Table 1 shows loadings of items onto each component.

**Table 1.**
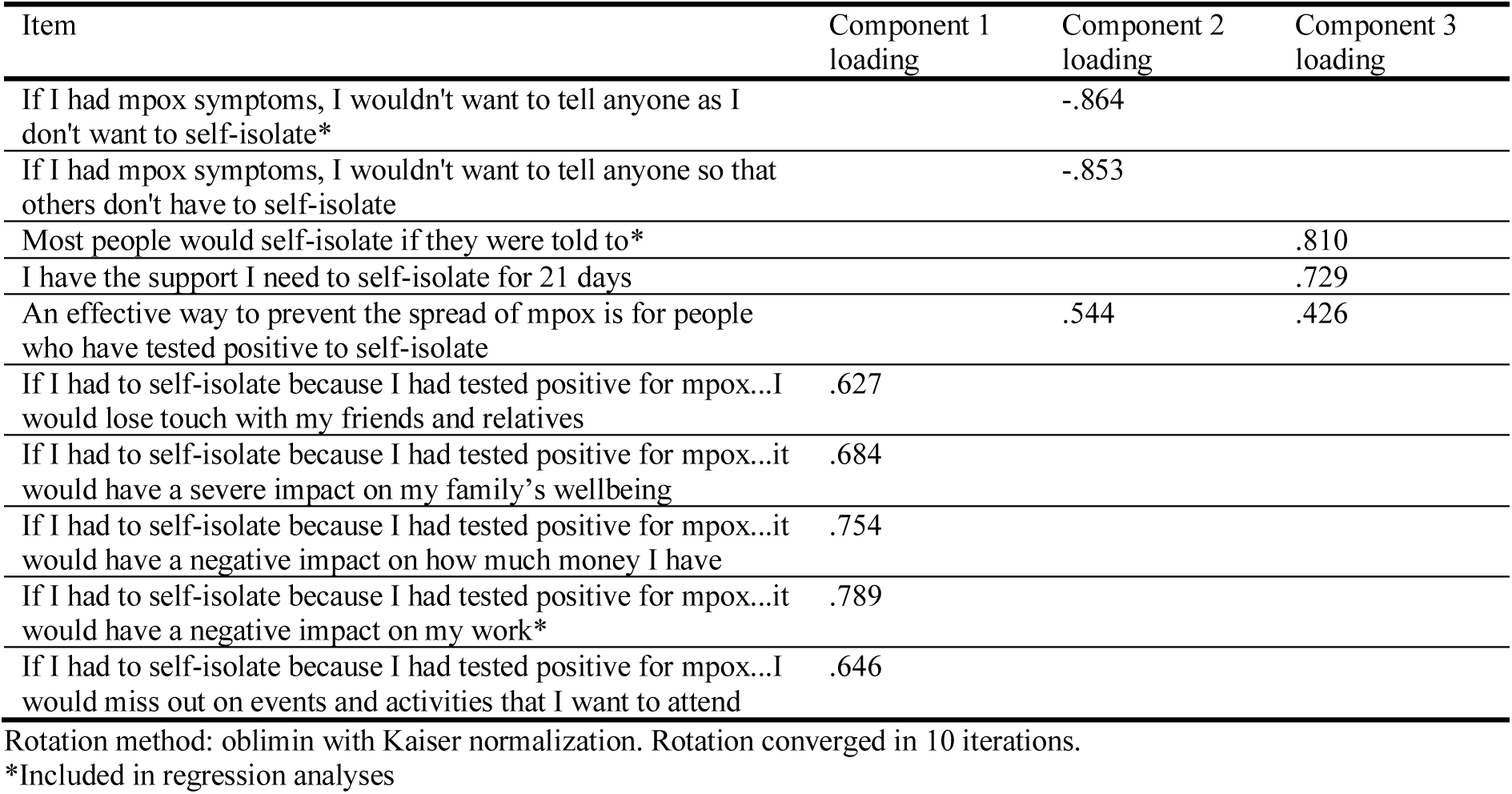
Loadings of items measuring factors potentially associated with self-isolation onto components identified (only loadings over ± .4 are shown).

### Help seeking

A scree plot suggested two components underlying factors potentially associated with seeking help immediately. Table 2 shows loadings of items onto each component.

**Table 2.**
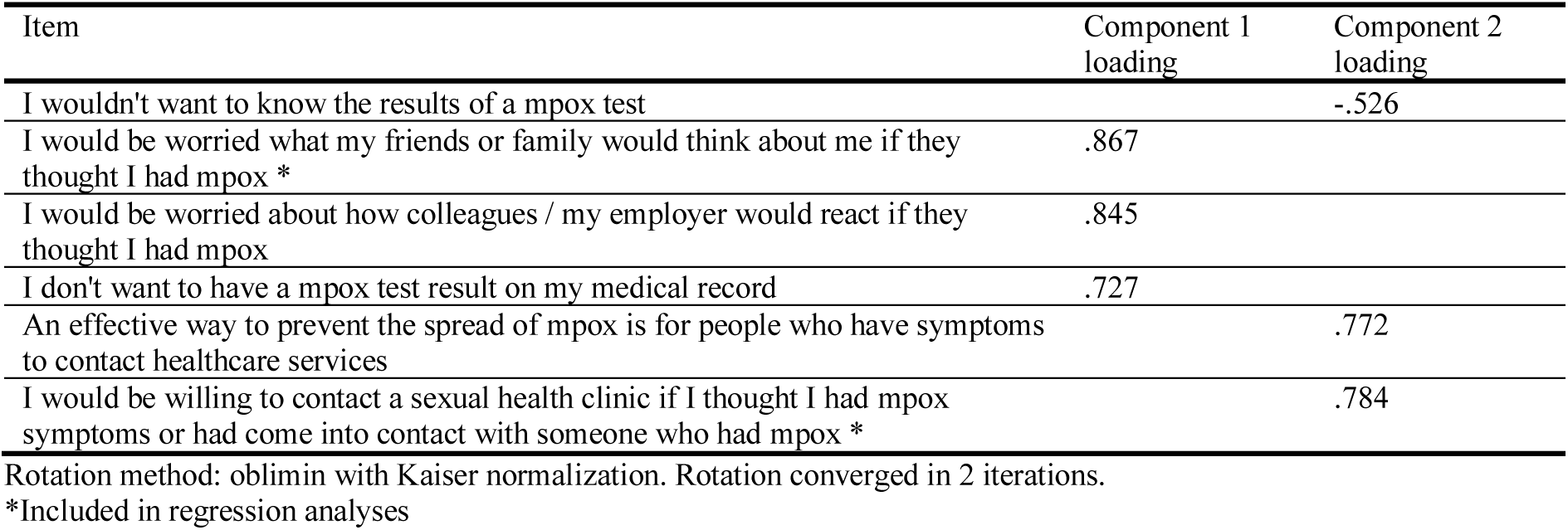
Loadings of items measuring factors potentially associated with self-isolation onto components identified (only loadings over ± .4 are shown).

### Vaccination

A scree plot suggested two components underlying factors potentially associated with vaccination. Table 3 shows loadings of items onto each component.

**Table 3.**
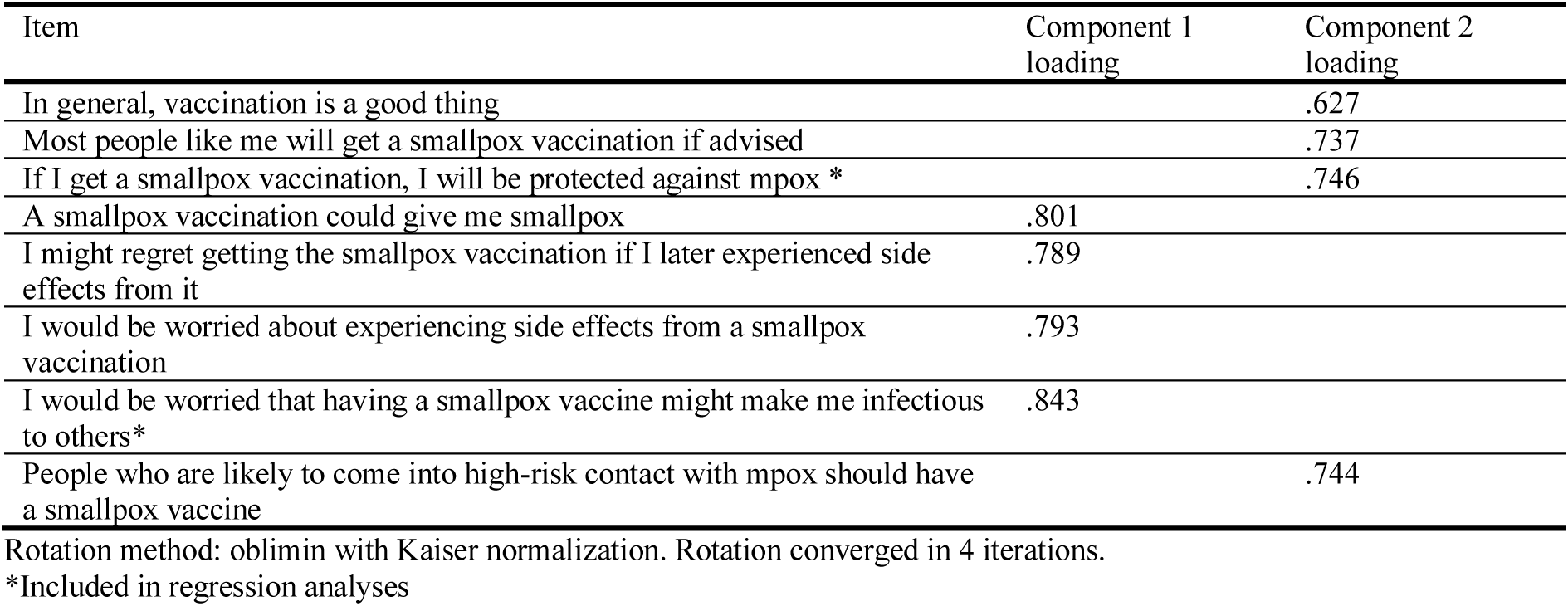
Loadings of items measuring factors potentially associated with vaccination onto components identified (only loadings over ± .4 are shown).

## Supplementary materials 4. Influence of motivational messages on behavioural outcomes

**Table 1.**
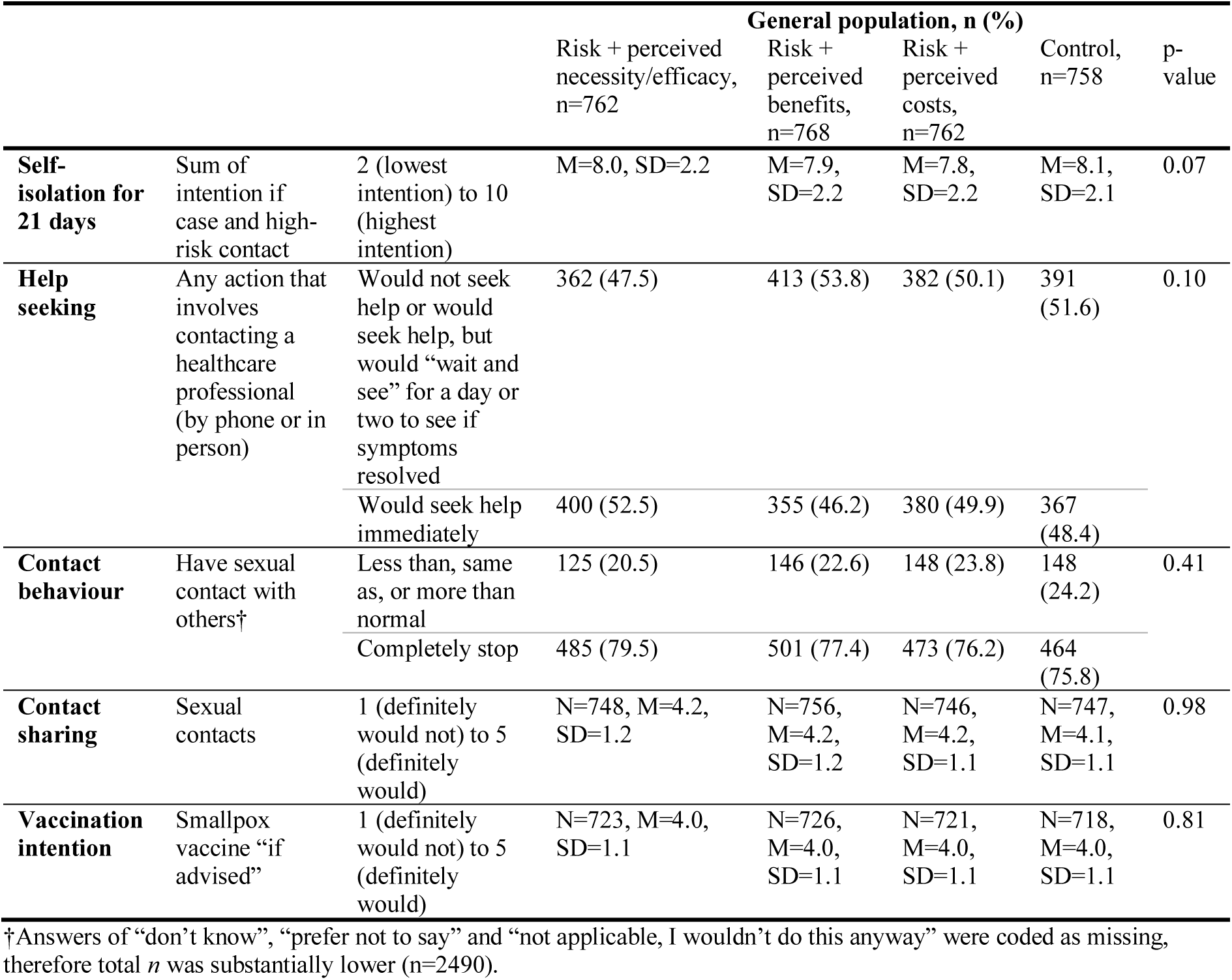
Influence of motivational messaging on behavioural outcomes, in the general population sample.

**Table 2.**
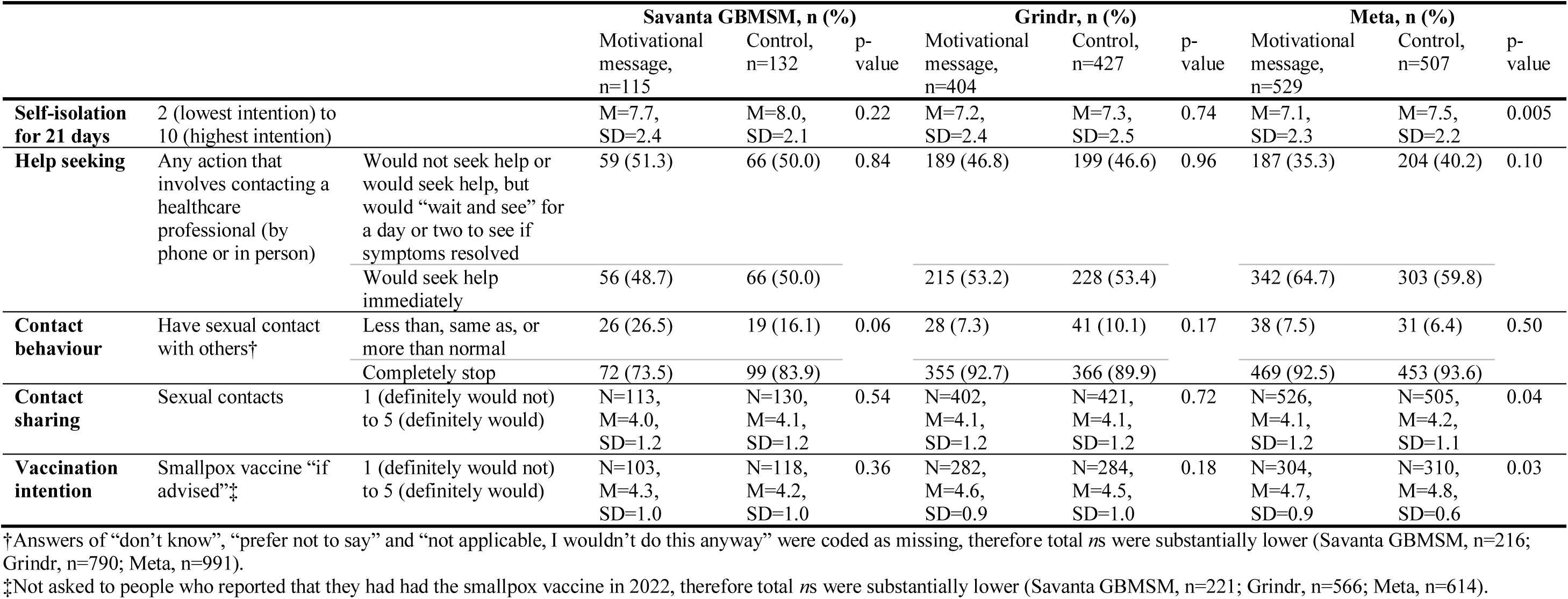
Influence of motivational messaging on behavioural outcomes, by GBMSM sample.

## Supplementary materials 5. Full results of regressions with intention to seek help immediately

**Table 1.**
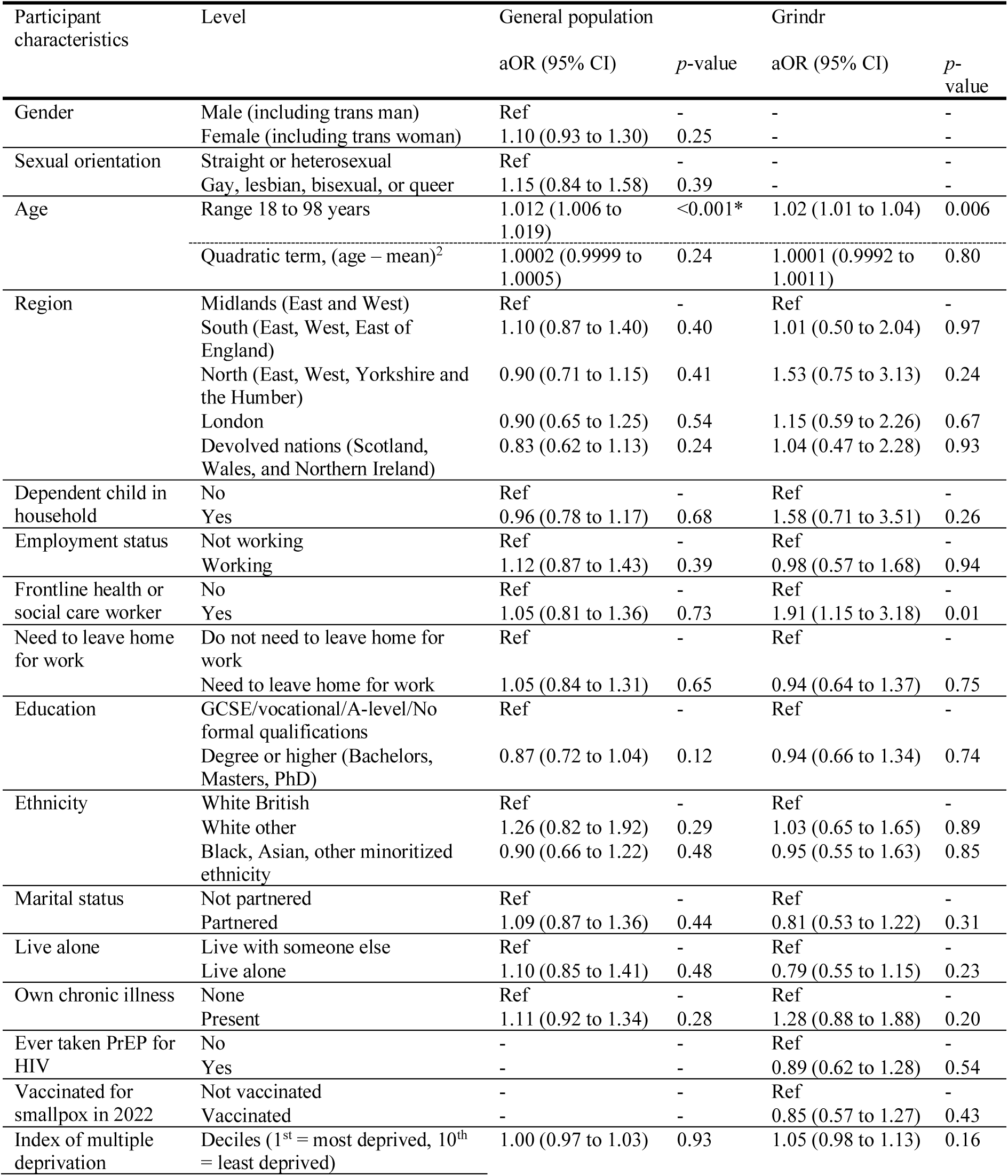
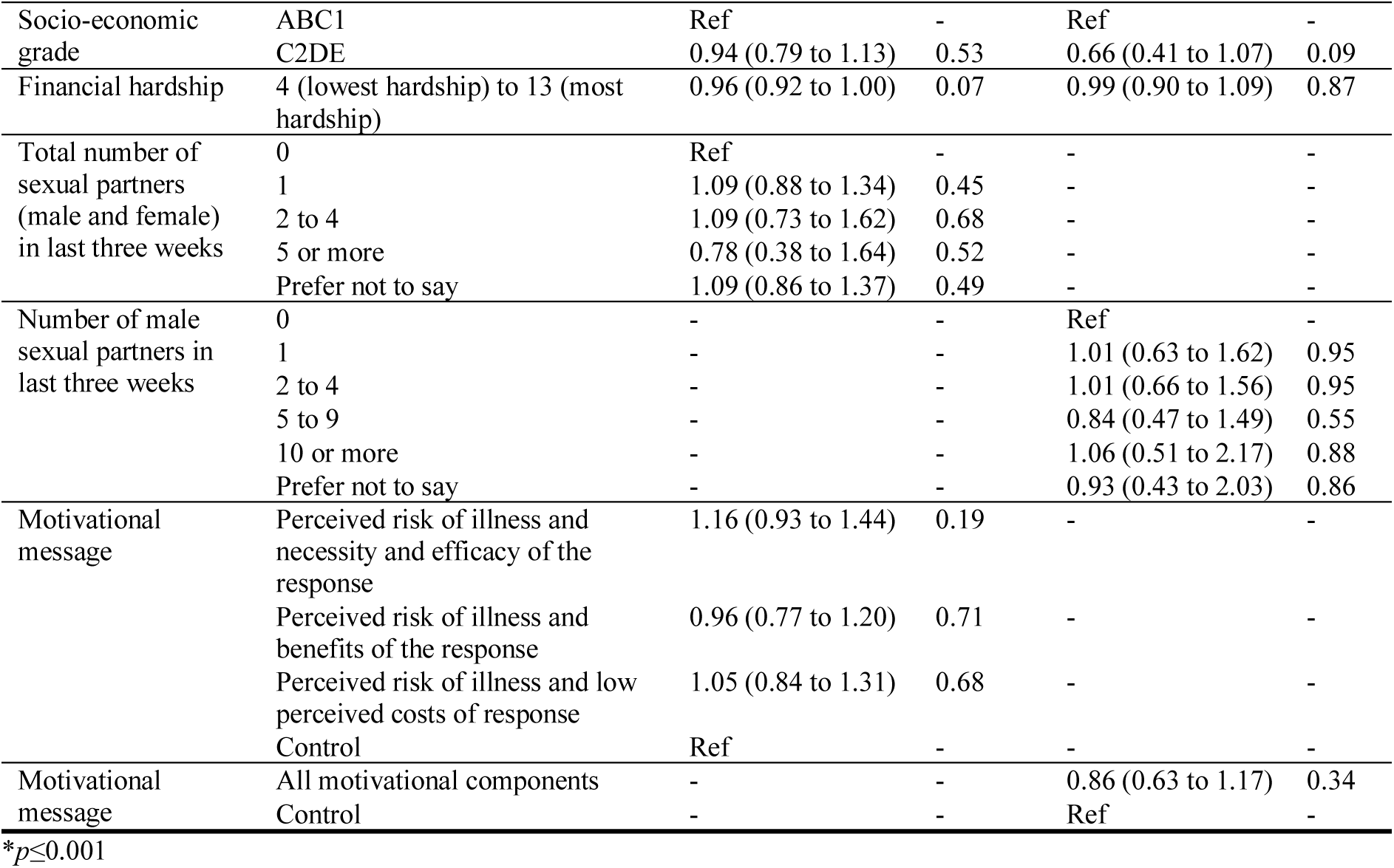
Associations between intending to seek help immediately and socio-demographic characteristics and motivational message, by sample. Variables were entered into the logistic regression model in blocks (block 1: socio-demographic variables and motivational message, block 2: psychological factors, block 3: help-seeking specific factors). Results for block 3, using pooled estimates are reported.

**Table 2.**
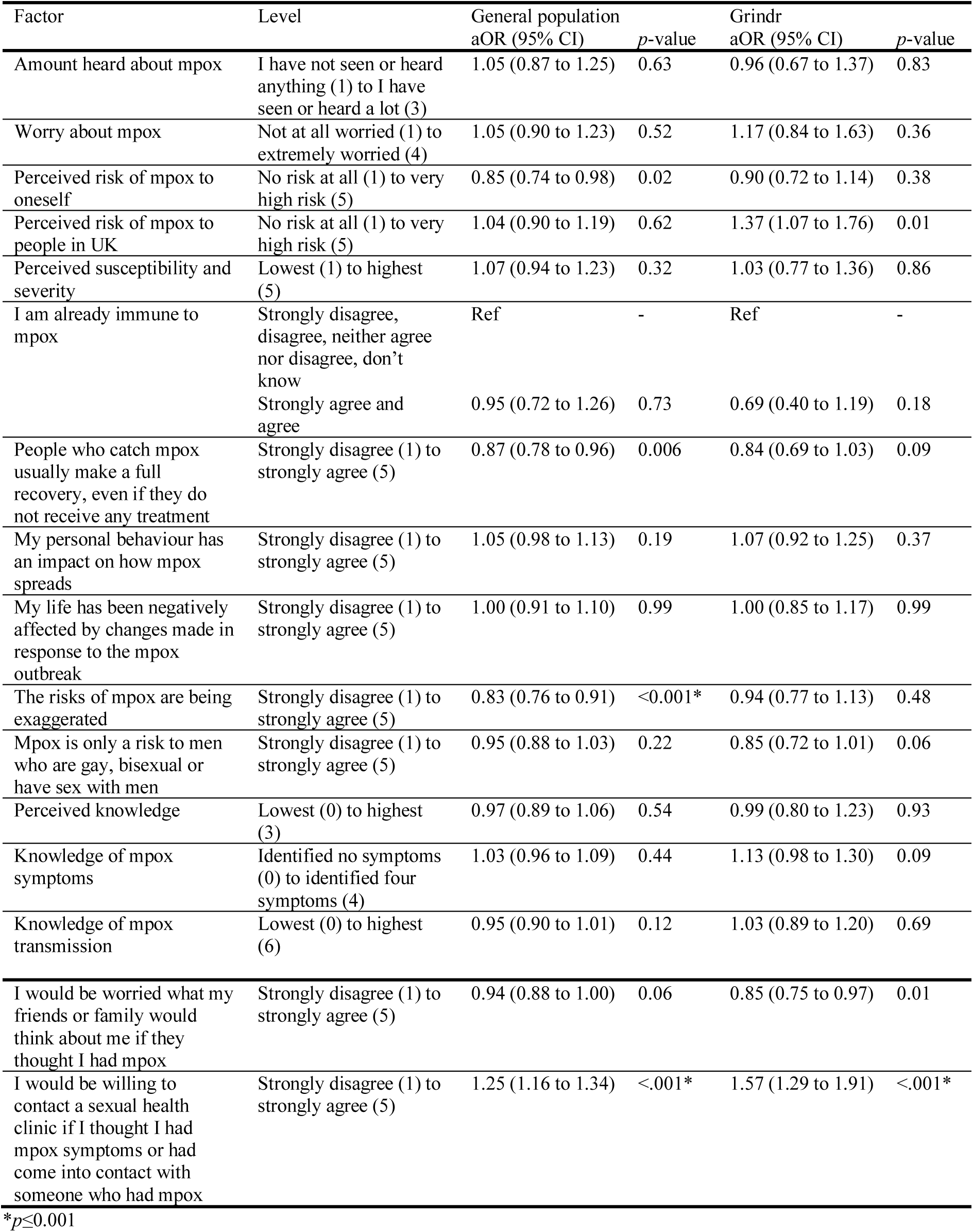
Associations between intending to seek help immediately and psychological and contextual factors, by sample. Variables were entered into the logistic regression model in blocks (block 1: socio-demographic variables and motivational message, block 2: psychological factors, block 3: help-seeking specific factors). Results for block 3, using pooled estimates are reported.

## Supplementary materials 6. Full results of regressions with smallpox vaccination

**Table 1.**
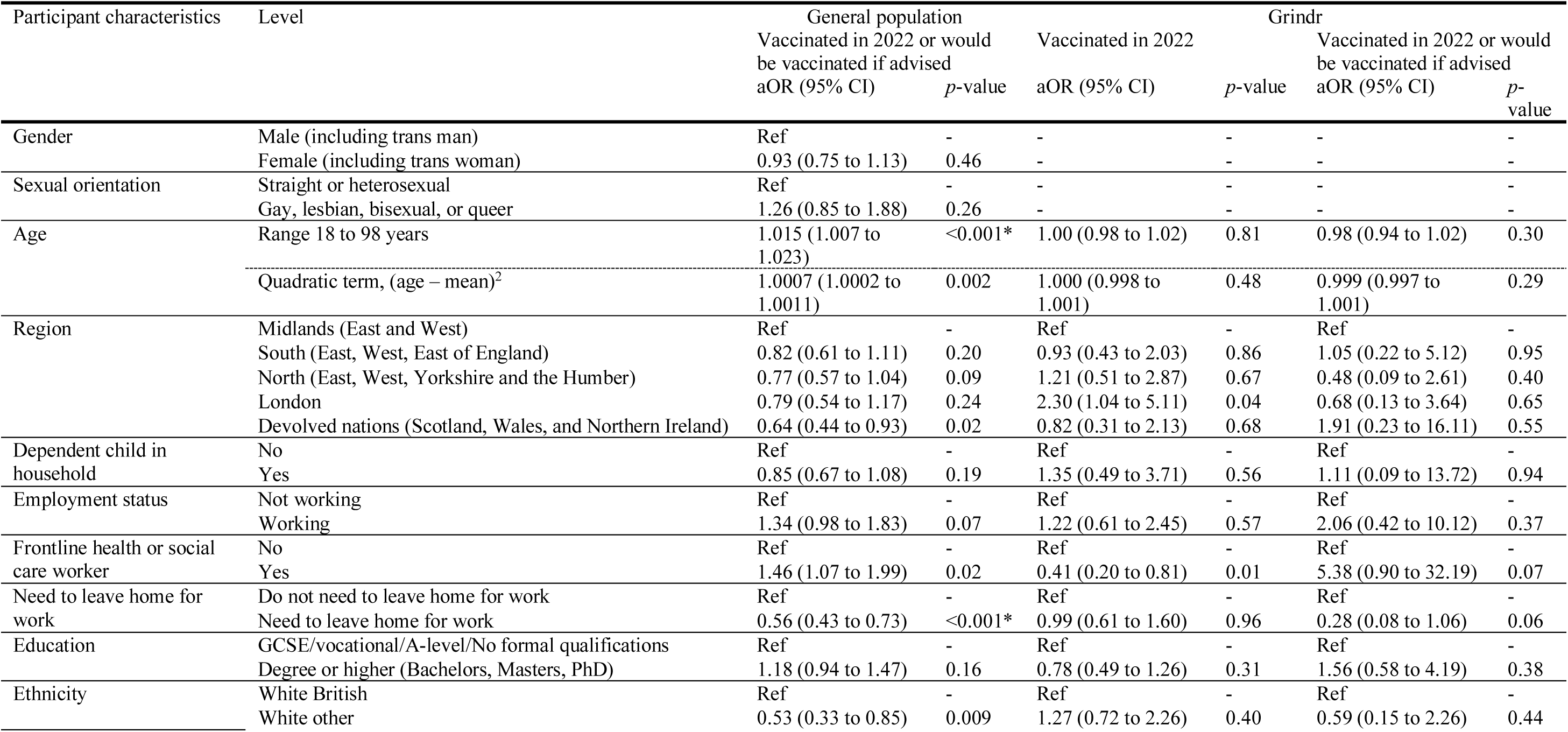
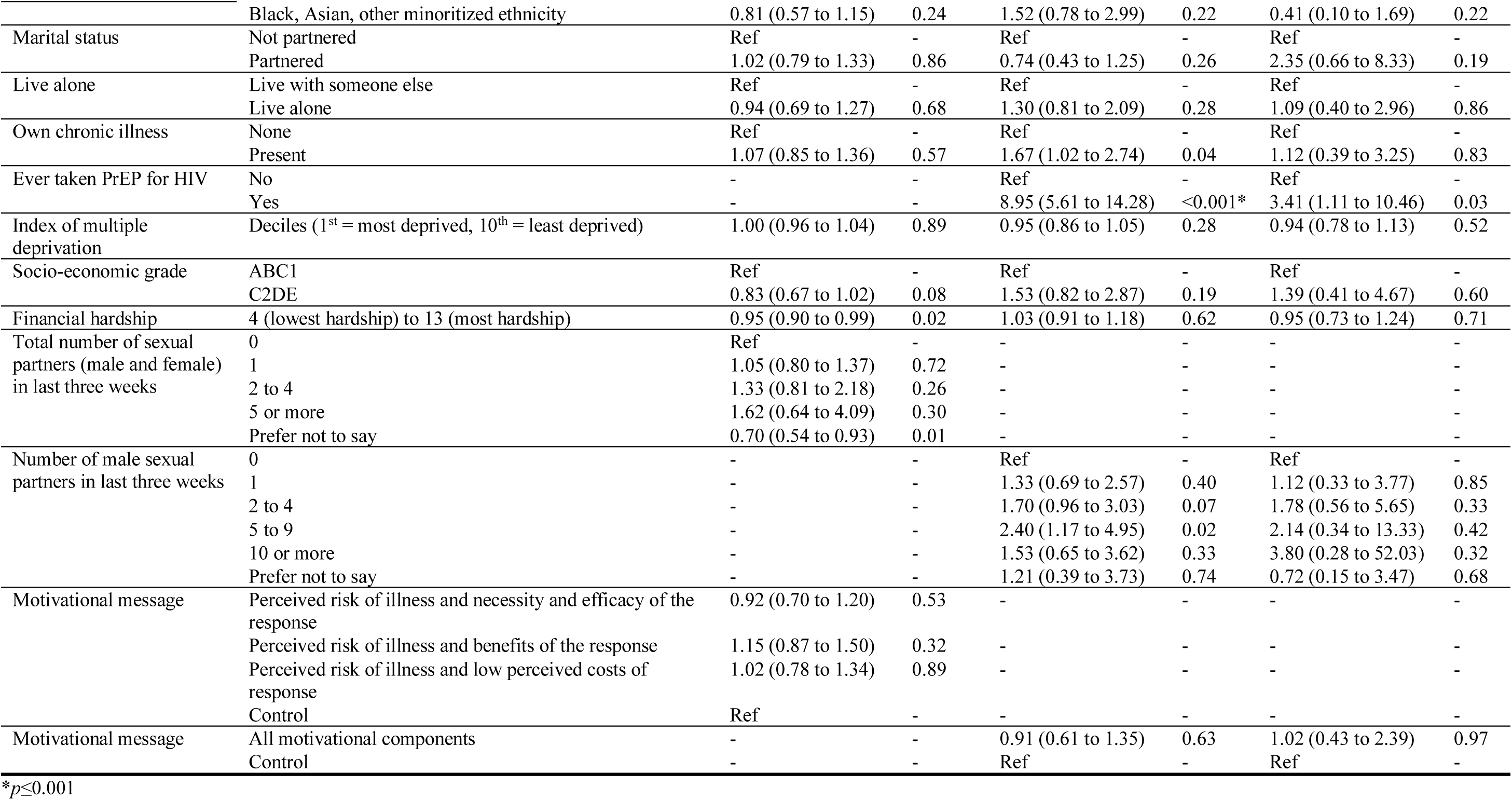
Associations between vaccine uptake (general population sample: actual and intended, GBMSM: actual, actual and intended) and socio-demographic characteristics and motivational message, by sample. Variables were entered into the logistic regression model in blocks (block 1: socio-demographic variables and motivational message, block 2: psychological factors, block 3: vaccination specific factors). Results for block 3, using pooled estimates are reported.

**Table 2.**
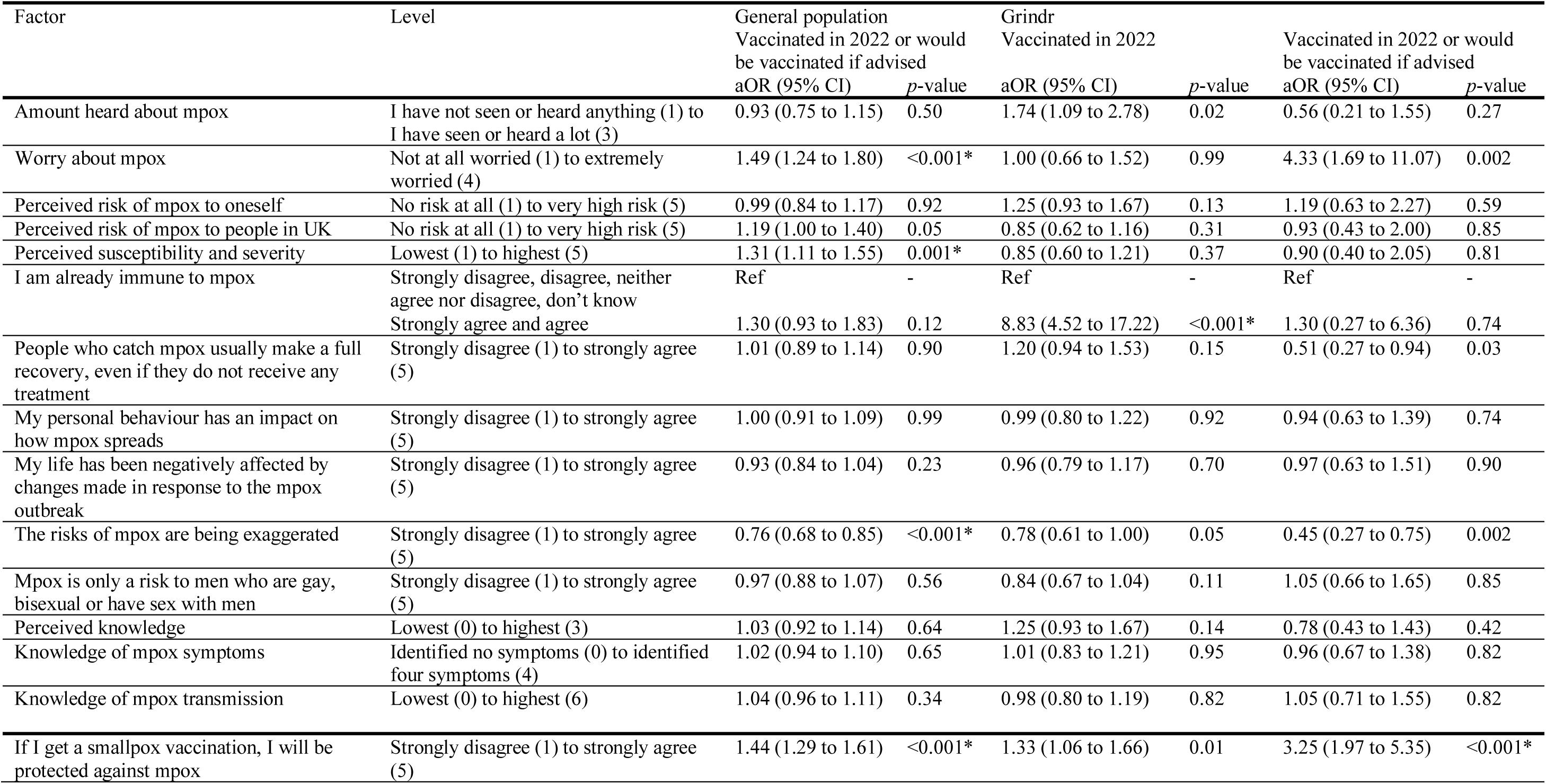
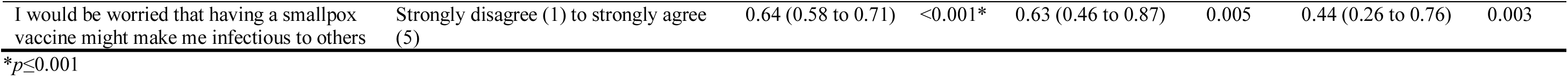
Associations between vaccine uptake (general population sample: actual and intended, GBMSM: actual, actual and intended) and psychological and contextual factors, by sample. Variables were entered into the logistic regression model in blocks (block 1: socio-demographic variables and motivational message, block 2: psychological factors, block 3: vaccination specific factors). Results for block 3, using pooled estimates are reported.

